# From metric to action: The decision value of infectious disease forecasts

**DOI:** 10.1101/2025.07.20.25331802

**Authors:** Cathal Mills, Nicholas J. Irons, Joseph L.-H. Tsui, Sarah Sparrow, Luiz M. Carvalho, Adam J. Kucharski, Oliver Ratmann, Ben Lambert, Christl A. Donnelly, Moritz U. G. Kraemer

## Abstract

Decisions for infectious disease outbreaks are difficult and consequential, required to be made in the face of considerable time and societal pressure and great uncertainty. Public health decisions can be supported by probabilistic forecasts – predictions of the future value of an epidemiological quantity, together with uncertainty. Forecasting is challenged by noisy, incomplete, and delayed data alongside non-linear and changing dynamics. Evaluation metrics for infectious disease forecasts often focus on the forecaster’s perspective; improving calibration and sharpness of forecasts. Currently, there are no systematic evaluation protocols to explicitly measure a forecast’s “value” – its ability to provide actionable insights for decision-makers. We here develop a systematic forecast evaluation framework for informing epidemic decision-making, focusing on three aspects: i) translating forecasts and popular evaluation metrics into public-health-relevant quantities; ii) defining evaluation metrics for decision-makers; iii) linking predictability of an epidemic to the value of forecasts for decision-making. By melding concepts from weather forecasting, information theory, and decision theory, our framework bridges conceptual gaps between forecasters and decision-makers. We illustrate the framework with an application to forecasts of weekly incident COVID-19 cases and find that ensemble models often provided the most value for decision-makers with varying levels of risk appetite. Focusing forecast evaluations on the decision-maker provides a new perspective for infectious disease modellers with the hope for improved public health decision-making in future outbreaks and epidemics.

**Significance statement:** Infectious disease forecasts are often used to support public health decisions, yet forecasts are typically evaluated using statistical measures that do not necessarily reflect their practical value; to inform decisions. We develop a new framework that connects forecast evaluation to decision-making by incorporating risk preferences and policy-relevant outcomes in evaluations. Drawing on concepts from decision theory, information theory, and weather forecasting, we introduce metrics that prioritise the decision-maker in evaluating forecasting models. Applying our framework to COVID-19 forecasts, we demonstrate that models performing best on average may not provide the greatest value for specific decisions or risk preferences. This work provides a foundation for aligning infectious disease forecasting with real-world decision-making and improving the use of forecasting in public health.

## 1 Introduction: The forecast–decision gap

> “Forecasts possess no intrinsic value. They acquire value through their ability to influence decisions made by users of the forecasts [1].”
>
> *Allan H. Murphy*

Infectious disease forecasting involves predicting future values of epidemiological indicators, such as disease-induced cases, hospitalisations or healthcare capacity. Forecasts differ from *scenario projections* by being unconditional predictions about the future; forecasts are the best estimate of what will happen. Short-term forecasts can quantitatively inform outbreak responses, while seasonal forecasts can inform public health capacity planning. Depending on the targeted quantity and possible actions (e.g., increase bed capacity or delivery of vaccines) of decision-makers, the predicted future can be i) modifiable (e.g., COVID-19 cases following vaccination strategies with direct impacts), or ii) largely invariant to interventions (e.g., seasonal dengue cases following expansion of healthcare capacity). A unifying goal in all forecasting tasks is to support the forecast *user* (i.e., a decision-maker) by providing the necessary scientific understanding and knowledge for making informed decisions.

There has been rapid progress in infectious disease forecasting since the COVID-19 pandemic, particularly in collaborative modelling hubs, performance evaluation metrics, and forecasting ensembles. Scoring rules (which measure the whole forecast distribution rather than comparing point estimates) and detailed performance assessments are now commonplace for forecast evaluation [2–6].

Advances have mainly focused on forecast *quality* (alignment of probabilistic forecasts with observations), yet fewer examples translate infectious disease forecasts to *value* for decision-makers (i.e., benefit to the forecast user to make decisions). However, an important recent development [7] considers decision-maker resources and advocates for new evaluation metrics linked to policy performance.

What remains unclear is how to systematically evaluate infectious disease forecasts for individual decision-makers under considerable uncertainty and societal, healthcare, and economic pressures. Although there are differences with infectious disease forecasting [8], in weather forecasting, evaluation procedures measure how well a model’s forecast informs the user’s (decision-makers) ability to take action for extreme weather events (e.g., an example is the decision to implement mitigation efforts for reducing the impact or to advise on travel avoidance) [9, 10].

Relevant questions for infectious disease forecasts remain understudied. Is a model “good” at producing forecasts that inform important epidemic decisions? Can a decision-maker take appropriate action to mitigate epidemic risks using statistical metrics and forecasting models? How do model rankings and implied decisions vary with different risk preferences, resources, and shifting epidemic dynamics?

Here we develop an evaluation framework for infectious disease forecasters and decision-makers to i) understand what drives the forecast performance of models (Section 2), ii) quantify the forecast value for specific epidemic events and decision-makers (Section 4), and iii) link epidemic predictability to forecast performance and robust forecast-informed decisions (Section 5). The key contribution of our framework is shifting the perspective from forecasters to decision-makers and to enumerate the respective forecast *value*.

## 2 Interpreting forecast quality across space and time

We introduce here a new framework for evaluating forecasts (Figure 1), where systematic procedures identify a “best”/“optimal” forecasting model conditional on the priorities of the local decision-maker, affected population, and available data (Figure 2 A, Figure 1 and Section 6).

**Figure 1:**
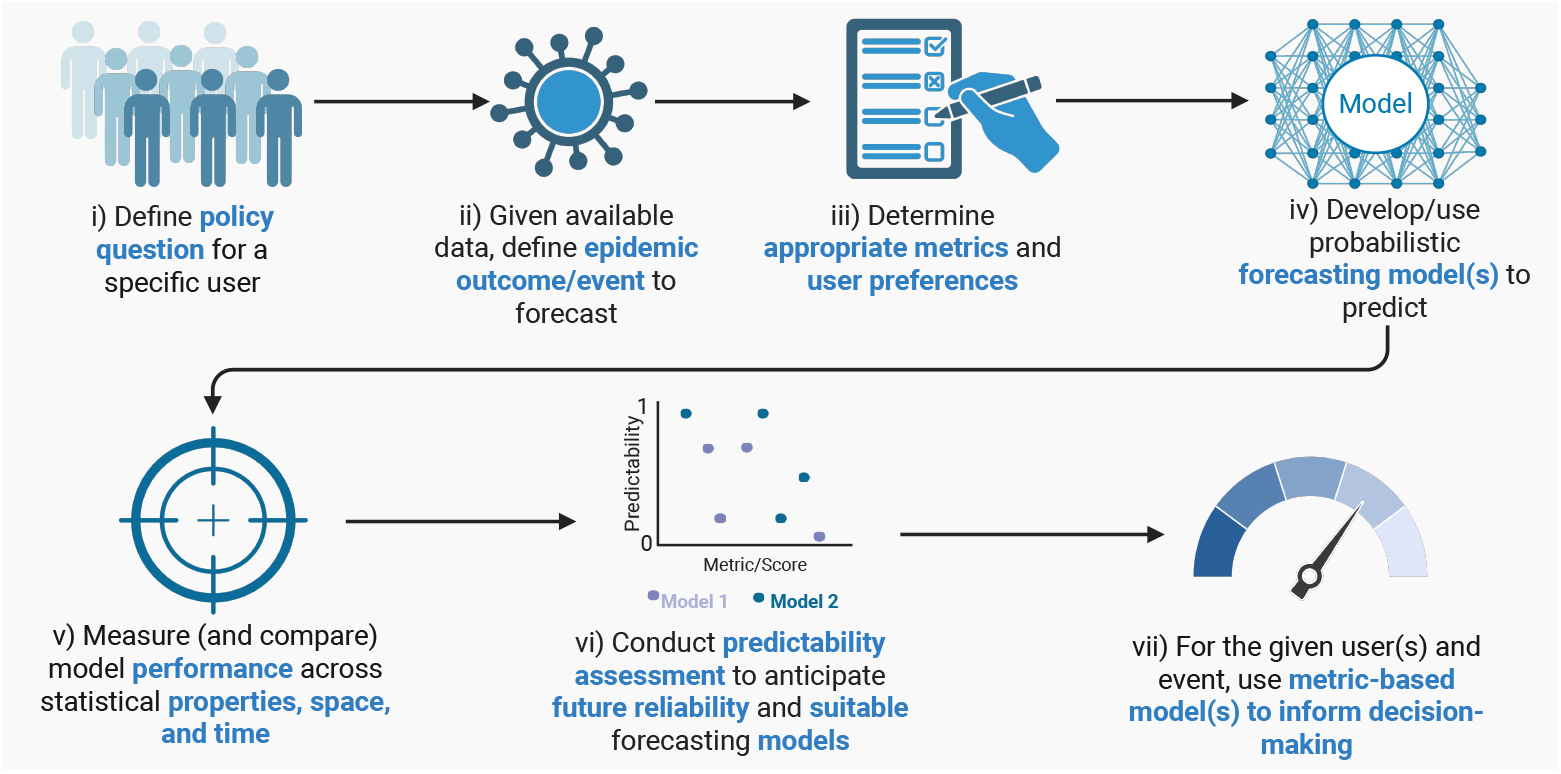
A new evaluation framework and workflow for forecasters and forecast evaluators: A proposed workflow that enables forecasters and evaluators to justify why a decision-maker would use the forecasts of a specific model (or set of models) to inform a public health policy decision. The workflow starts with, and emphasises throughout, a specific policy question of a given user/decision-maker. The workflow uses historical performance evaluations (v), yet also includes procedures to guard against historically optimal models no longer being optimal into the future. The workflow is to be applied iteratively at each time point (e.g., for multiple events and metrics), and the metric-based optimal model(s) for each decision will be based on one or more chosen metrics. For determining policy questions and an appropriate metric for a given user and policy question, see Figure 2 and Table 1. We provide details of metric-based and predictability assessments throughout Sections 2–5. See Section 6 for applications of the framework to COVID-19 case forecast evaluation.

**Figure 2:**
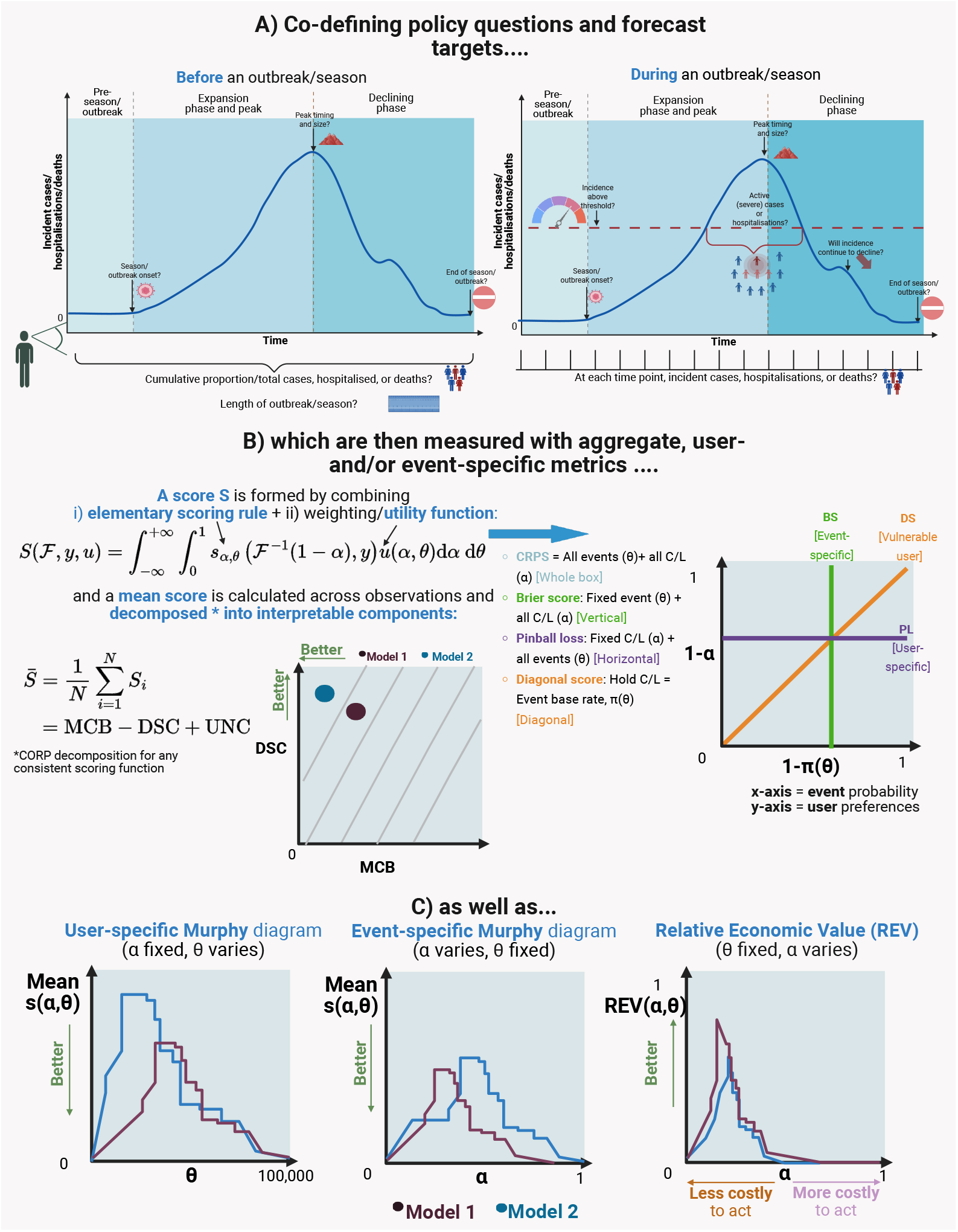
Policy questions, forecast targets, and evaluation metrics for different users and events: **A)** Generic epidemic curves illustrate example policy questions, either before or during an outbreak. Forecast evaluation in our framework emphasise these defined policy questions. **B)** Popular summary scores (e.g., CRPS) are formed by combining an elementary score and utility function, and each score makes implicit assumptions about i) the epidemic event which is captured by event threshold *θ*, and ii) the decision-maker whose preferences for the event are expressed by the C/L ratio *α*. These scores can be further decomposed into the statistical drivers of forecast performance for enhanced interpretation (see main text). **C)** User- and event-specific evaluation tools – Murphy diagrams and Relative Economic Value curves where lower and higher curves respectively indicate better forecast value.

### 2.1 Scoring functions and rules

Trust in forecast models is established by evaluating performance of forecasting models across space and time and understanding what drives forecasting performance. Scoring functions and rules measure calibration and sharpness of forecasts. Typically, lower values in these scores imply better performance (see Table SI 1). A scoring rule is (strictly) proper if the score is minimised, in expectation, by (only) the unknown, true probability distribution of the target quantity. Examples include the continuous rank probability score (CRPS) [11], Weighted Interval Score (WIS) [12], and Brier score (BS) [13]. Relative scores (e.g., relative WIS, rWIS [14]), while not strictly a proper scoring rule, compare forecast performance against a baseline model. We use WIS and CRPS as our framework’s proper scoring rules measuring overall predictive performance (Figure 2 B) for forecasting the continuous value of a target epidemiological quantity *y*.

**Table 1:**
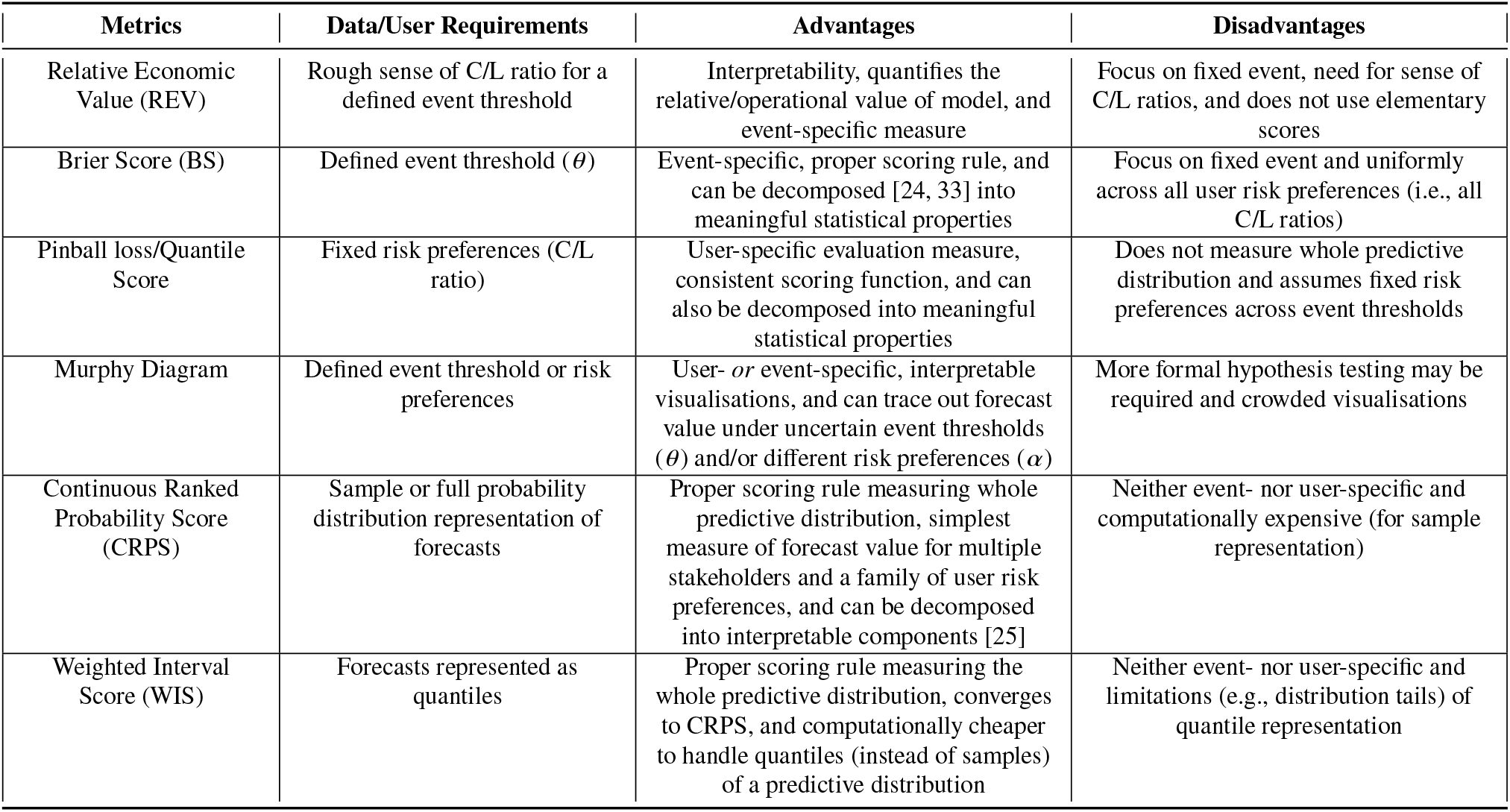
Metrics of our framework: Details of different metrics for the quality and value of probabilistic forecasts, including the information needed to compute metrics and relative advantages and disadvantages of each metric.

| Metrics | Data/User Requirements | Advantages | Disadvantages |
| --- | --- | --- | --- |
| Relative Economic Value (REV) | Rough sense of C/L ratio for a defined event threshold | Interpretability, quantifies the relative/operational value of model, and event-specific measure | Focus on fixed event, need for sense of C/L ratios, and does not use elementary scores |
| Brier Score (BS) | Defined event threshold ($\theta$) | Event-specific, proper scoring rule, and can be decomposed [24, 33] into meaningful statistical properties | Focus on fixed event and uniformly across all user risk preferences (i.e., all C/L ratios) |
| Pinball loss/Quantile Score | Fixed risk preferences (C/L ratio) | User-specific evaluation measure, consistent scoring function, and can also be decomposed into meaningful statistical properties | Does not measure whole predictive distribution and assumes fixed risk preferences across event thresholds |
| Murphy Diagram | Defined event threshold or risk preferences | User- *or* event-specific, interpretable visualisations, and can trace out forecast value under uncertain event thresholds ($\theta$) and/or different risk preferences ($\alpha$) | More formal hypothesis testing may be required and crowded visualisations |
| Continuous Ranked Probability Score (CRPS) | Sample or full probability distribution representation of forecasts | Proper scoring rule measuring whole predictive distribution, simplest measure of forecast value for multiple stakeholders and a family of user risk preferences, and can be decomposed into interpretable components [25] | Neither event- nor user-specific and computationally expensive (for sample representation) |
| Weighted Interval Score (WIS) | Forecasts represented as quantiles | Proper scoring rule measuring the whole predictive distribution, converges to CRPS, and computationally cheaper to handle quantiles (instead of samples) of a predictive distribution | Neither event- nor user-specific and limitations (e.g., distribution tails) of quantile representation |

The BS is an *event-specific* (Figure 2 B) classification measure that assumes uniform risk preferences of a decision-maker [13]. Meanwhile, pinball loss (or quantile score) is a scoring function to assess forecasts for a fixed, *user-specific* (Figure 2 B) quantile level *τ*, with higher *τ* values indicating more vulnerability (or less risk tolerance).

### 2.2 Stratifications and interpretable scoring metrics

For any scoring function or rule, the mean score 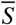 across *N* test set observations is often used to rank forecasting skill, yet can obscure i) variations in forecast performance under different epidemic conditions (e.g., temporal [15], feature [16], or spatial [4] dependencies) and ii) variation in statistical components of forecast performance (e.g., calibration versus discrimination abilities).

As epidemics often exhibit strong spatial and temporal signatures, similar to other studies [4, 16–18], we propose that all evaluation metrics are applied to different test data set stratifications, including trailing windows and different spatial units and scales (see applications in Section 6).

Uncovering determinants of the metric-implied strengths and weaknesses is important for improving models and enhancing trust. Such statistical properties control the public health utility of the information contained in forecasts. Scoring functions and rules can be decomposed for useful interpretation, and while rarely used in infectious disease forecasting, *decompositions* explain forecast performance [19–25]. To decompose 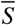, we use CORP (Consistent, Optimally binned, Reproducible, and Pool-adjacent-violators)-algorithm decompositions [24–26]:

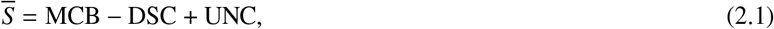

where non-negative quantities MCB, DSC, and UNC denote miscalibration, discrimination, and uncertainty. Lower values of MCB indicate superior calibration (reliability), while higher values of DSC indicate superior discrimination ability. The uncertainty component measures the irreducible variability of the data.

In our framework, we visualise CORP-based discrimination and calibration properties (Figure 2 B), where DSC (y-axis) is plotted against MCB (x-axis), with points indicating models and lines indicating constant forecast performance. Diagnosing discrimination or calibration issues across space, time, or features guides forecasters where additional data or bias correction may be required and evaluators about ensemble model weights and likely suitable models for different decision-makers. This ensures that forecasts and evaluation focus on relevant risk preferences and priorities of a decision-maker. For example, if systematically underpredicting at high quantile levels, forecast recalibration may be required to accommodate relevant risk preferences (probability levels).

## 3 Introducing the cost-loss decision framework for infectious disease forecasting

### 3.1 Scoring forecasts for decision value

The “best” average model may not be best for individual decision-makers or individual epidemic events (e.g., tail risks).

It is not often recognised in infectious disease forecasting that many popular scoring metrics (specifically, consistent scoring functions) are intricately linked to cost-loss (C/L) ratios (Figure 2 B). These scores are the output of *weighting* an *elementary score* (see below) by a utility function or mixing measure that collectively capture the forecast’s value using the user’s risk aversion (C/L ratio) for different event thresholds [27, 28].

The error scoring matrix is defined as:

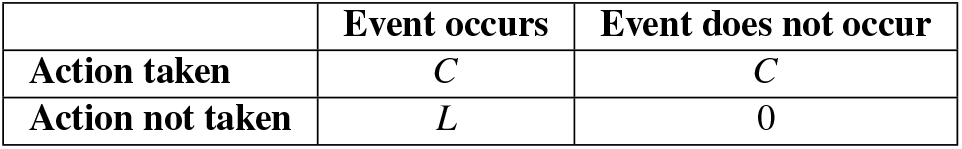

*C* will represent economic and social costs (see Section 3.2) for taking a specific preventive action (e.g., increasing intensive care unit (ICU) capacity), and the preventable loss *L* will be the *additional* impact (e.g., loss of life) if the pre-specified, binary event (e.g., severe outbreak) occurs without taking action. Loss still occurs if the event occurs and action is taken, but *L* is defined as the loss that could have been prevented by using the forecast for impact mitigation.

This C/L framework assumes that forecasts are independent of the realised outcome, and that a forecast can alter costs and losses incurred, but *not* the event probability (see Discussion).

The *elementary score* [27] can be defined for an event threshold *θ* ∈ ℝ in the outcome space (binary event occurs if *y > θ*) and C/L ratio *α* in probability space (the ratio of the cost of taking action versus the *preventable* loss if no action is taken):

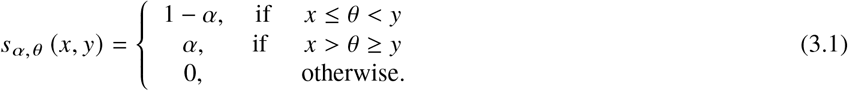

where *x* ∈ ℝ is the (1 − *α*)−quantile forecast, *x* := *F*^−1^(1 − *α*). Eq. (3.1) assigns penalties 1 − *α* for an underprediction when an event occurs and *α* for an overprediction when an event does not occur.

Under the C/L framework, the decision-maker should act when predicted probability of the event ℙ (*y* ≥ *θ*) ≥ *α*. Small *C/L* values indicate inexpensive risk mitigation relative to reducible losses for the event, the optimal action will be to act even when the forecasted event probability is low, and Eq. (3.1) will penalise an underprediction with a higher/worse elementary score.

A *summary score* weights the elementary score (eq. (3.1)) with a *utility* function *u*(*α, θ*) [27, 28].

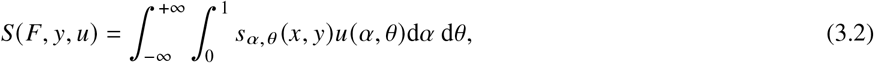

where *x* ∈ ℝ is the forecast at the (1 − *α*) −quantile level *x* := *F*^−1^(1 − *α*), *u*(*α, θ*) determines how the decision-maker weights simultaneously different combinations of C/L ratios *α* and epidemic event thresholds *θ*.

Eq. (3.2) establishes a key result for forecasters and evaluators: a summary score must make assumptions about the decision-maker and their risk preferences *α* and priorities *θ*. For example, the CRPS integrates over all probability decision thresholds *α* and all binary epidemic event thresholds *θ* (Figure 2 B), and weights thresholds by the density of the outcome distribution and weights C/L ratios quadratically by errors. CRPS produces the simplest single measure of *average* forecast value across decision-maker’s risk preferences for all epidemic events [29], yet such assumptions can make CRPS less suitable for specific decision-makers and in a fast-evolving epidemic, e.g. strongly emphasising decision-makers with moderate C/L ratios around 0.5 whose decisions are most sensitive to the forecast (and therefore contribute most to scores), rather than extremes.

### 3.2 Eliciting risk preferences for an epidemic

Plausible C/L ratios *α* for different *θ* must be defined and updated for individual decision-makers across space and time. These will *never* be known with certainty – epidemics and decisions have profound health, social, and economic impacts, forecasting models are often miscalibrated, and there will inevitably be residual uncertainty, ethical judgements, and legal implications. Many costs and losses can be unforeseen or difficult to quantify, yet through decision-maker surveys, real-time collaboration and frequent communication, and existing quantitative estimates [30–32], forecasters and evaluators can consider reasonable ranges of C/L ratios, depending on how decision-makers prioritise different types of costs depending on their resources and preferences.

For example, suppose a local decision-maker will allocate another 10 ICU beds to a district if more than 1,000 (*θ*) infections occur (the event) in a given week. ICU beds are made available at an economic cost if a chosen model (see below) forecasts infections exceeding the threshold in the next week with probability ≥ C/L (*α*). The economic costs of action (*C*) could depend not only on additional beds but also the probable length of patient stays, additional medical and staffing costs, and indirect costs if beds are relocated from elsewhere. Meanwhile, if the event occurs without preventive action, preventable losses (*L*) could entail life costs from severe disease or macroeconomic impacts.

C/L ratios will vary for different epidemic event thresholds (e.g. lower risk preference for higher numbers of infections) and across space and time by pathogen and transmission history (e.g., cumulative hospitalisations), and by decision-maker, population health, and available resources (e.g., resource-limited setting requiring greater certainty). By evaluating forecast value over ranges of uncertain C/L ratios and event thresholds, forecasters and evaluators can explore the Pareto frontier of forecasts that are performant simultaneously relative to multiple objectives. We emphasise that evaluation only requires the C/L ratio for an event, that is the relative cost of mitigation.

## 4 Measuring forecast value for decision-making

### 4.1 User-and-event-focused scoring metrics

When confronted with a new pathogen, the coming season of an endemic pathogen, or rise/decline in epidemic trajectory, decision-makers face difficult and specific decisions (Figure 2 A), for which a model’s *value* may vary. These decisions require models and evaluation to refocus on specific epidemic events, areas of the forecast distribution, and decision-makers’ risk preferences. We suggest here interpretable metrics and visualisations that provide such targeted assessments of forecast decision value.

Viewing each decision problem as a *binary* one (action or no action), the *value* of a model’s forecasts is determined by the ability to inform the decision problem(s) for different decision-makers, thus depending on both the forecasting model and the decision-maker’s risk preferences and priorities for the specific policy question. This framing does not render a forecast the sole basis for complex epidemic decision-making, but rather establishes that the value of a forecast must be its ability to inform decision-makers.

#### 4.1.1 Relative Economic Value

For a decision-maker’s chosen event threshold (*θ*) and corresponding C/L ratios (*α*), Relative Economic Value (REV) measures the operational, decision value of a model’s forecasts *relative* to the forecasts of a baseline/reference model and perfect model [29, 34, 35].

For event threshold *θ*, REV starts by converting the forecasted exceedance probability (*p*_*i*_ = ℙ (*y*_*i*_ *> θ*) under the forecast distribution) into a classification of the binary event *y*_*i*_ ∈ {0, 1} based on a given decision rule. Under the C/L decision rule, a forecasting model predicts occurrence of the binary event if the probability *p*_*i*_ exceeds C/L ratio *α*. REV then uses the expected expense (*E*) that would be incurred by using the model’s classifications for the binary event. Expected expense is calculated by combining the correct and incorrect classifications with corresponding costs and loss incurred, yet the calculation of REV relies only on knowing the ratio of costs and losses (see SI B):

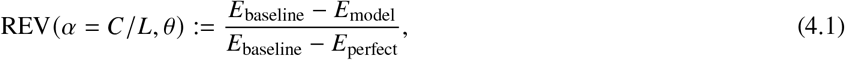

where values of 0 and 1 represent the baseline and perfect/oracle model respectively, REV is computed for different C/L ratios (*α*) while holding the event threshold (*θ*) constant. REV is then shown as a curve for different decision-maker risk preferences (*α*), with higher values indicating superior operational/relative value (Figure 2 C, Table 1).

#### 4.1.2 User-or event-specific Murphy diagrams

A Murphy diagram [27] provides user-or event-specific measures of forecast value (Figure 2 C, Table 1), using the BS as a proper scoring rule or consistent scoring function (pinball loss) without the need for a baseline model.

As a *user-specific* evaluation tool, the Murphy curve (Figure 2 C) visualises, for a fixed risk preference or C/L ratio *α*, the mean elementary score (y-axis) for a given forecasting model against different epidemic event thresholds *θ* (x-axis) across *all N* test set observations:

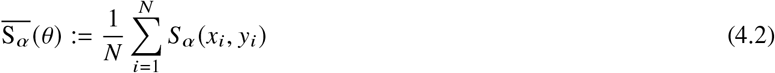

where for observation *i, x*_*i*_ ∈ ℝ is the model’s forecast at quantile level 1 − *α, y*_*i*_ *∈* ℝ is the observed outcome, and *S*_*α*_ is the corresponding elementary score (eq. (3.1)) for the fixed C/L ratio *α* and a specific event threshold *θ*. In our framework, we include Murphy diagrams, a collection of Murphy curves that trace out, for a fixed risk aversion preference (*α*) of a decision-maker, how the value of different models’ forecasts varies across different magnitudes of epidemic events (*θ*). Lower curves for a given model are preferable, and if curves for different models cross, there may be no model dominance [26, 27]. The area under each Murphy curve is the mean pinball loss for that model. This visualisation improves on the BS as it measures value for a specific decision-maker’s risk preferences and different epidemic event thresholds, which may be advantageous when epidemic events are difficult to *a priori* define.

Alternatively, a Murphy curve can be an *event-specific* evaluation tool (Figure 2 C), holding the epidemic event threshold *θ* fixed and visualising a model’s mean elementary score 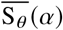 (y-axis) against different C/L ratios *α* (x-axis):

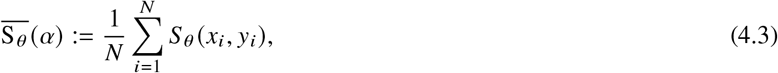

where *S*_*θ*_ is the elementary score (eq. (3.1)) for the fixed event threshold *θ* and a specific C/L ratio *α*. The area under such a Murphy curve is the mean BS, and a Murphy diagram again visualises Murphy curves for different models. This diagram traces out, for a fixed epidemic event severity (*θ*), how the forecast value of different models varies with different risk preferences (*α*) for the decision-maker.

Caution is necessary, as: a user-specific Murphy diagram does not measure the whole predictive distribution (and risk preferences will likely vary for different event thresholds), while the event-specific Murphy diagram focuses on a single event threshold; neither diagram segments forecast performance into discrimination or calibration; and formal inference and hypothesis tests may be required.

## 5 Safeguarding epidemic decision-making

A historically optimal model is not necessarily optimal for the future. Epidemic decision-making is high-stakes; decision-makers may prioritise risk-averse strategies that remain robust to extreme events and uncertainty. Above, we described uncertainty in C/L ratios which could affect forecast value, model selection, and metric-implied optimal decisions. Meanwhile, epidemiological data are imperfect, impacted by reporting errors, delayed reporting, and underrepresentation. These issues, along with fast-evolving and space-varying epidemic dynamics, can induce data distribution shifts and lead to metric-implied optimal models for decisions – from results for past test data (Sections 2–4) no longer being optimal when making the decision for the future. To protect against uncertainties, tail risks, and spatiotemporal dependencies (in data and forecast value), we recommend distributionally robust optimisation (DRO, Section I) and predictability analyses.

Focusing on predictability, epidemics are dynamical systems with no governing laws where observed data are imperfect measures of the true nonlinear epidemic dynamics. We broadly define predictability as the inherent randomness of the epidemic time series data (see below) that induces fundamental limits in the ability to predict the future values of the time series.

### 5.1 Introducing predictability for infectious disease forecasting

Forecast *quality* could be bounded above by epidemic predictability, yet forecast *value* could relate to epidemic predictability in different ways.

A “good” forecasting model could be valuable for decision-making during epidemic stages of rapid change and high unpredictability. The association between predictability and forecast value will depend on how we define each. Predictability is a magnitude-free property of the dynamical system, whereas forecast value is measured for a decision-maker and/or epidemic event, thereby depending on the magnitude of the observed data and/or the set of models. Thus, predictability will not have a one-to-one relationship with forecast value.

There are currently few procedures to know which forecast outputs can be reliably reported. Here, we describe how to assess the effects of predictability on historical forecast quality and value, and how to anticipate future predictability and the decision utility of a forecasting model.

In our framework, we hypothesise that i) assessing past predictability of the epidemic time series will help to determine whether past model forecast performance was largely caused by the model and/or the (un)predictability of the epidemic, and ii) analysing past predictability can inform the likely future predictability and forecast performance of different models.

### 5.2 Permutation entropy

For estimating predictability, following [36], we use permutation entropy (PE), a model-free measure of temporal dependence in potentially nonlinear time series data. Entropy summarises uncertainty in a system. Shannon’s entropy [37], for a random variable *X* ∈ 𝒳, where 𝒳 is the sample space, is defined as *H* (*X*) = − ∑_*x*∈𝒳_ ℙ (*X* = *x*) log ℙ (*X* = *x*).

For time series ***X*** = {*x*_*t*_ : *t* = 1,…, *N*}, PE requires defining: i) the embedding dimension *d* which determines the segment length for measuring randomness/patterns in the data and ii) the time delay *τ* which determines the number of time steps between the compared measurements (see below). These are used to construct vectors: 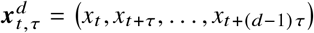. We fix *τ* = 1 to focus on successive values of the time series.

Within each vector 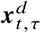, entries of this segment of length *d* are then (totally) ordered by comparing their relative magnitudes. This produces an ordinal pattern *π*. Normalised PE is then defined by using the Shannon entropy on these patterns and normalising using the maximum entropy log(*d* !):

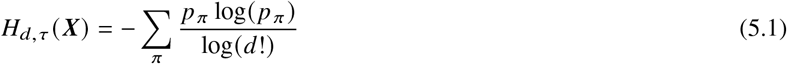

where *p*_*π*_ is the empirical probability of a specific ordinal pattern *π*, based on the frequency at which each pattern occurred.

To promote conservative estimates of PE, we again follow [36], and define predictability as:

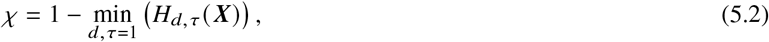

where the minimisation searches over values from *d* = 2 to a user-defined maximum dimension (see [36] for more details). Using eq. (5.2), we can compare epidemic predictability (*χ*) across space and time using trailing windows of length *N* and region-specific time series data, noting that i) the window length *N*, over which PE is calculated, can be varied, but must be greater than *d*!, and ii) PE (and therefore *χ*) is global-magnitude-invariant as it compares relative magnitudes of observations. Values of *χ* vary between 0 (low predictability) and 1 (high predictability).

### 5.3 Embedding predictability in our framework

In predictability assessments of our framework (Figure 1), values of *χ* can be compared to evaluation results in trailing windows for the different metrics of Sections 2–4. Although low predictability *could* suggest low forecastability, i) high predictability does not guarantee high forecast quality or value and ii) low predictability does not guarantee low forecast value.

Understanding which models provide the most relative improvements in forecast quality or value over baseline/reference models during different regimes of predictability is one way to link predictability to forecast quality and value. Then, an estimated recent or future low predictability will have setting-specific implications – this may provide caution *or* optimism about using, for decision-making, a given model’s forecast of the future.

Given that i) an operational forecast is often required to support decision-makers irrespective of how predictable an outbreak is, and ii) data distribution shifts are pervasive in epidemics, these predictability analyses after historical evaluations provide a useful sense-check and additional layer to evaluation. These analyses help to quantify the practical/operational value at different stages of the epidemic and protect against future erosion of public or decision-maker trust. For example, PE values could indicate regime changes, which could inform model selection or data collection. PE could also be used by forecasters as an input to a forecasting model or as an indicator of a future change in other epidemic indicators (e.g., in the time-varying reproduction number *R*(t)).

## 6 Applying metrics to epidemic forecasts

### 6.1 Application of our framework

We have introduced a conceptual framework for which the workflow (Figure 1) could be as follows. Given the available data, the first step for forecasters and evaluators is to: i) co-define a relevant policy question with local decision-makers, ii) identify a corresponding epidemic outcome to measure (e.g., incident cases or healthcare capacity), iii) identify appropriate metrics and decision-maker preferences, iv) produce/collect probabilistic forecasts, v) measure forecast performance for each model across statistical properties, over time, and across space, and vi) assess historical predictability of epidemic dynamics alongside past forecast performance to anticipate future reliability and model suitability, and vii) use metric-based results to choose the “optimal” model’s forecasts for informing future decisions where optimality is only with respect to a given user and event. As decisions will not be made on the basis of a single question, metric, or epidemic outcome, this evaluation workflow should be repeated multiple times at each time point, using different policy questions and different outcomes and metrics targeting the same policy question.

We illustrate this framework with a brief application (Table SI 2 for EPIFORGE 2020 [38] checklist items), using state-level forecasts of weekly incident cases (on a logarithmic scale [5]) from the COVID-19 Forecast Hub (see [16, 39, 40]).

We studied 1 August 2020 – 15 January 2022 inclusive and focused on six models (Table SI 3) that submitted forecasts for all weeks. We used the COVID-19 Forecast Hub baseline model as a reference model [40].

Submitted forecasts were summarised using seven quantiles, preventing the evaluation of the full forecast distribution (and all C/L ratios). We adopted a 12-week trailing window to examine epidemic predictability and time-varying model performance. We used the same weekly estimates as [16] for the instantaneous reproduction number, *R*(*t*). All code and data used in our analysis are available at https://github.com/cathalmills/forecast_evaluation, and in-depth results are included in SI Section H.

### 6.2 Evaluation workflow results

Using WIS and rWIS, the ensemble model generally outperformed other models, including across forecast horizons, locations, and time points (Figure SI 1 A), for individual locations (Figure SI 1 B), and in trailing windows of length 12 weeks (Figure SI 1 C, SI 3 – SI 6).

Zooming on specific risk preferences and specific extreme incidence events, the ensemble, baseline, and Karlen-pypm model often obtained the highest forecast value across DSC and MCB for pinball loss and Brier score (Figure 3 A). Recognising that it can be unrealistic to assume fixed risk preferences across all event thresholds, we computed Murphy diagrams for specific decision-makers across different event thresholds and for specific events across different risk preferences (Figure 3 C.1 – C.2), thereby showing how the ensemble, baseline, and Karlen-pypm models often obtained the highest forecast value for many, but not all, risk-priority combinations (Figure 3 C.1 – C.2). These diagrams capture forecast value when there is uncertainty in the defined event (e.g., evolving dynamics) and/or decision-maker’s C/L ratio (e.g., likely uncertainty about cost of intervention or epidemic event loss). REV plots capture the ensemble and Karlen-pypm models’ high relative value for anticipating different extreme events for a range of decision-maker risk preferences, especially for shorter horizons (Figure 3 C.3, Figures SI 11 –SI 14).

**Figure 3:**
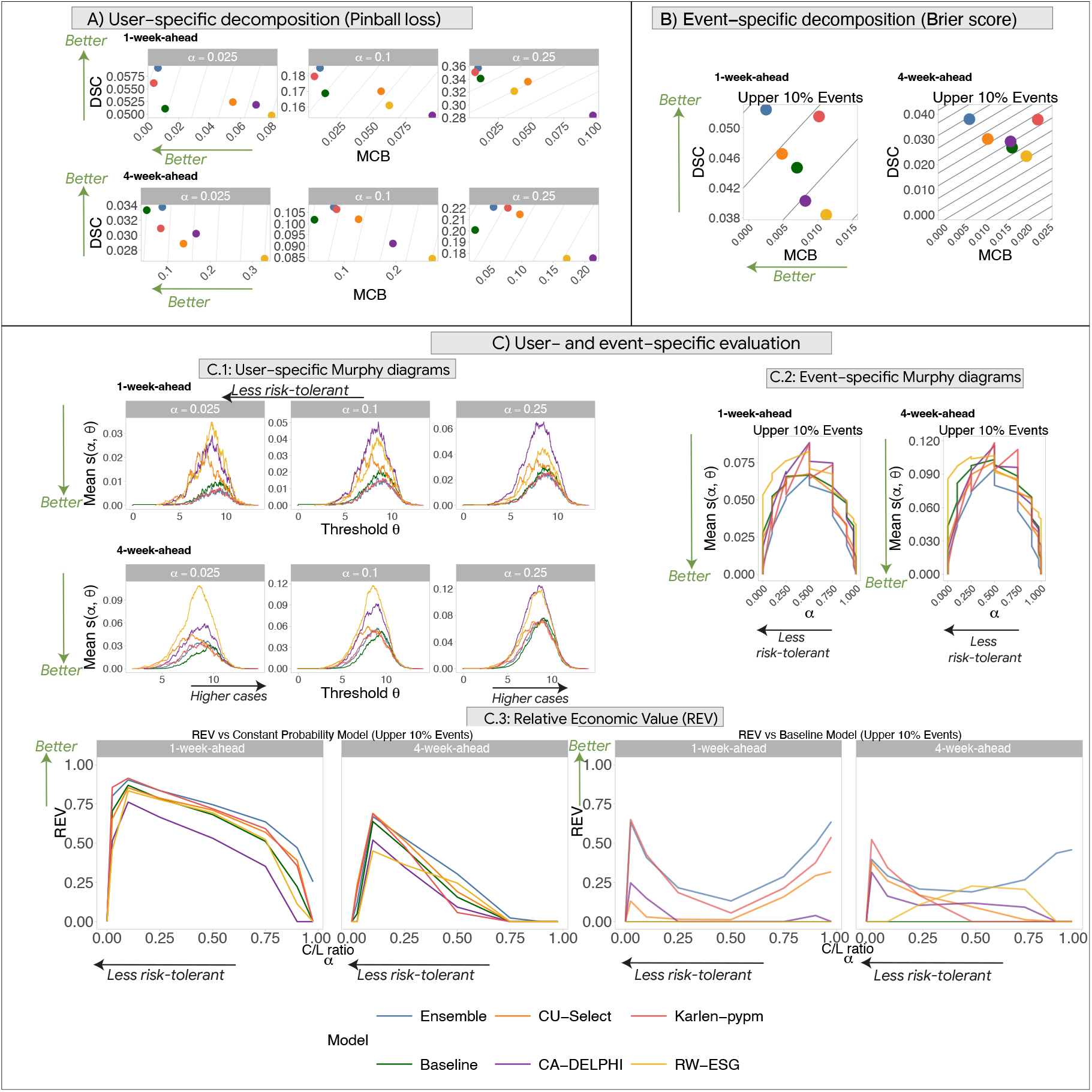
Application forecast evaluation results for our framework using state-level COVID-19 forecasts (on logarithmic scale) from 1 August 2020 – 15 January 2022. **A)** Pinball loss, a user-specific measure of forecast value, using discrimination (DSC) ability versus miscalibration (MCB). Higher DSC and lower MCB values indicate superior forecast value for a specific decision-maker’s C/L ratio *α* and grey lines indicate constant forecast value. **B)** Analogous decomposition for the Brier Score, an event-specific measure of forecast value, focusing on the upper 10% of each state’s observed cases. **C)** User- and event-specific evaluation measures of forecast value including event-specific Murphy diagrams (C.1) which capture the mean elementary score *s* (*α, θ*) for fixed C/L ratio (*α*) and varying event thresholds (*θ*), user-specific Murphy diagrams (C.2) which capture mean *s*(*α, θ*) for a fixed event threshold (*θ* is the state-specific upper 10% threshold of cases) and varying C/L ratio (*α*), and Relative Economic Value (REV) diagrams (C.3) for the upper 10% of each state’s cases. Lower values on Murphy curves and higher values on REV curves indicate superior performance of a model’s forecasts for a given user-event combination. Metric details are summarised in Table SI 1 and Figure 2.

Higher epidemic predictability was moderately yet significantly (Table SI 4) correlated with higher *R*^2^ across different horizons (Figures SI 32 and SI 27). Several models generally had *greater* relative performance when epidemic dynamics were *less* predictable (Figures SI 32, SI 29, SI 25). This does not mean that models necessarily provided improvements over a baseline model at such times, as there were generally consistent benefits, across space, time, and metrics, of using the ensemble model over the baseline model for decision-making during less *and* more predictable times, especially for longer forecast horizons (Figures SI 15 – SI 29).

We considered whether such predictability can be anticipated and what drives predictability regimes. Higher predictability within a window was strongly correlated with higher *R*(*t*) estimates within the window and in past windows, while higher predictability was correlated with higher levels of incident cases in the recent past (Figures 4, SI 30). While this does not imply any causal relationships, predictability was greatest during the epidemic growth and peak phases and we found overall low predictability that varied across space and time (Figures SI 30 – SI 31). While beyond our scope of evaluation, this may be useful for model development, ensemble weighting, model selection, and anticipating the likely future reliability of outputs. For instance, for several models, higher predictability was correlated with lower coverage of central 95% prediction intervals (PIs) (Figure 4, Table SI 5). This reflects findings from [16] where coverage dropped during growth phases which we estimated to be generally more predictable. During such phases, central 95% PI coverage, even from the ensemble’s forecasts, was often much lower than 95%, suggesting a need for updating of ensemble weights and/or models and their inputs.

**Figure 4:**
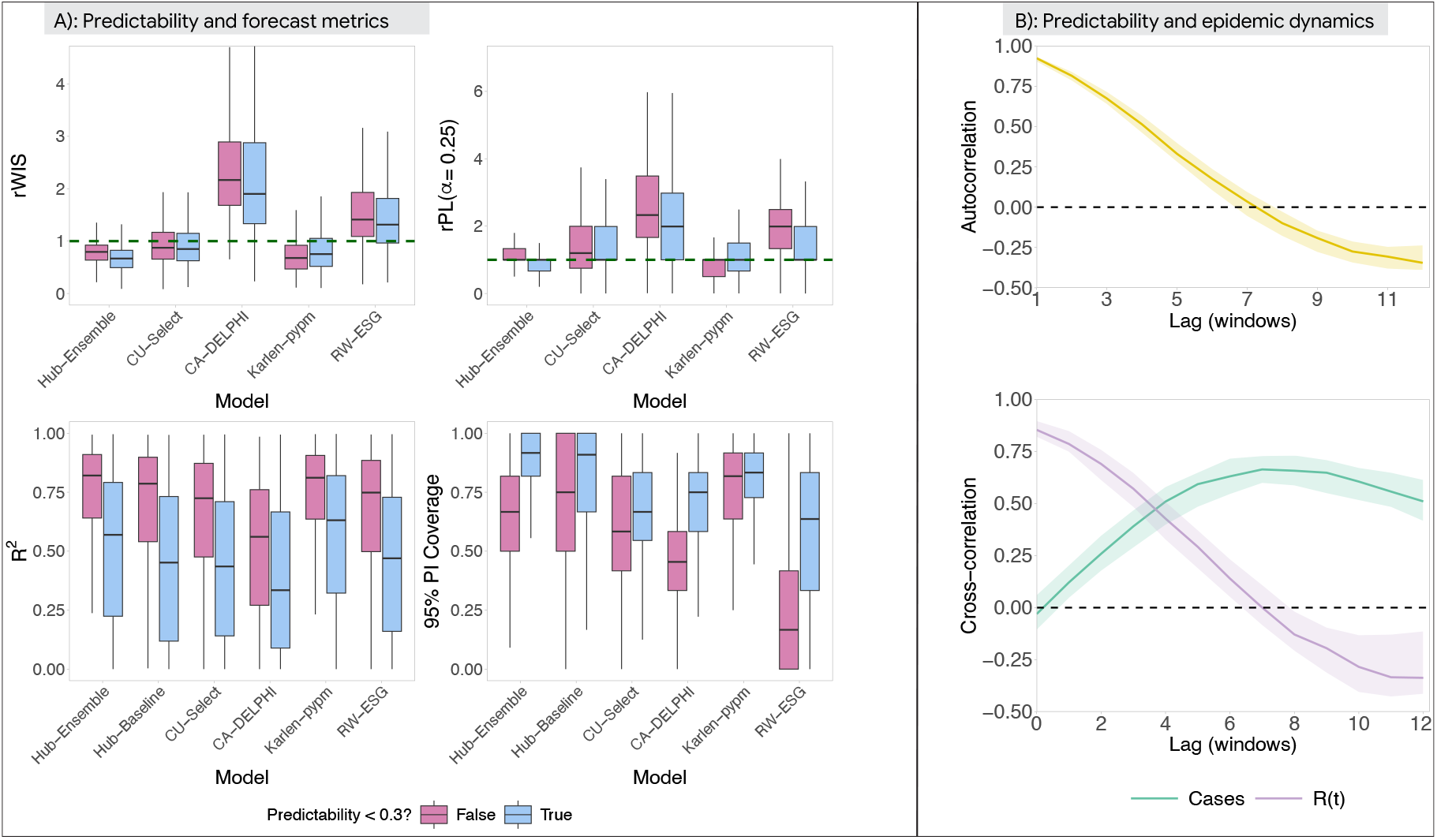
COVID-19 predictability and forecast value results. Predictability analyses include boxplots (A) of the relationship between predictability (binary thresholds here, further analyses in Tables SI 4 – SI 6, Figures SI 27 – SI 29) and forecast metrics, alongside median autocorrelation (B) of predictability and cross-correlation of predictability with cases and *R*(*t*), time-varying reproduction number estimates. Shaded intervals capture the interquartile range across locations. Metric details are summarised in Table SI 1 and Figure 2.

Finally, to focus on local decision-makers, we applied our framework to a randomly chosen state (Arizona) and estimated that different forecasting models provided highest, real-time decision value for different decision-maker risk preferences before and during a large COVID-19 wave (Section H).

## 7 Discussion

Forecasting epidemics will continue to be challenging due to demographic, climatic, and behavioural feedbacks that are both nonlinear and highly stochastic. Here, for forecasters building actionable models for the needs of decision-makers and for evaluators evaluating how well models meet these needs, we develop procedures to: i) frame forecasting models’ statistical strengths and weaknesses in policy-relevant decision value across space and time, ii) select the “optimal” model for a decision-maker’s individual policy question and risk preferences, and iii) safeguard forecast-informed decision-making. This framework lays foundations for improving the ability of forecasts to inform decisions.

The framework provides a decision-maker with an “optimal” model for their decision problem, based on past performance and their defined action set and risk preferences. Optimality conditions are specific and pre-defined, yet we show how to safeguard against data distribution shifts and incorporate uncertainties in risk preferences and priorities. Our user-focused prioritisations reflect theory that a group-level utility function which fairly weights individual preferences is mathematically problematic and arguably unattainable [41, 42]. As individuals have different risk preferences and priorities, there is no single scoring function for a group that allows all individual users to select the same forecasting model for a given decision problem. Instead, a decision-maker’s individual policy question and risk preferences determine the model(s) that provided the most value for informing decisions. This systematic decision-theoretic framework makes clear the underlying subjective choices of decision-makers and enhances the credibility and defensibility of modelling for informing uncertain policymaking. Iterative application of our evaluation framework for different policy questions connects with other research on decision problems [43, 44] as part of *integrated* assessments for policymaking. Decision-making will never be based on a single policy question or forecast, epidemic event/outcome, or branch of science, yet if forecasting is to take place, evaluation must measure how well the forecasts inform decisions.

Our framework establishes connections between best practices and other branches of science (e.g., weather forecasting and decision theory) [45, 46]. It does not replace guidelines for reporting [38], evaluation [2, 16], model development [47, 48] or diagnostics [26, 49]. For operationalisation, continuous dialogue between modellers, social scientists, and decision-makers will be essential to improve our proposed workflow (Figure 1). Models, evaluation metrics, and epidemic trajectories can be complex, and stakeholders have different priorities and backgrounds. Communicating uncertainty to both technical experts and decision-makers is essential yet non-trivial [46, 50–52]. Clear, timely, and succinct communication will involve decreasing levels of technicality as information is passed up the decision-making chain/process, while preserving key outputs and uncertainty communication effects [53, 54].

Recent guidelines for prediction tools advocate for a 3-U (useful, usable, and used) framework [55], which we embrace for evaluation with public-health-relevant quantities, user-focused evaluations, and decision value metrics. A key next step is to elicit decision-makers’ C/L ratios (*α*) for different event thresholds (*θ*), requiring careful setting-specific considerations (Section 3.2). Without plausible ranges for these C/L ratios, the framework will remain abstract. Eliciting preferences will benefit from i) regular consultation and surveys with decision-makers and experts across multiple disciplines, and ii) economic evaluation literature for different user perspectives [56]. There will always be both heterogeneity and uncertainty as decisions are multidisciplinary tasks with ethical and human components [43]. However, real-time surveys could define the distributions of users’ C/L ratios and event thresholds for different epidemic questions, which can inform custom user- and event-focused evaluation metrics.

Depending on the target quantity, the predicted future can be modifiable, modifications that a forecasting model generally does not anticipate. The “best” model may be the one that promotes the decision-maker to implement interventions that directly impact epidemic dynamics, potentially different from the model that produces the most accurate forecast of the realised future. The actions of a decision-maker can impact the very probability of the epidemic events that a model aims to forecast. A probabilistic forecast integrates over uncertainty in many different counterfactual scenarios and is a special, *unconditional* case of a scenario projection [51] – a model’s best estimate of what *will* happen.

Forecasts can be operationally advantageous for empowering decision-makers using predicted probabilities for the most likely future states of the world. This avoids conditional probability statements and evaluation in inherently counterfactual scenario projections. Model-based modifications in the intervening period after the date of forecast generation can increase limits to predictability, and this feedback from the future means that evaluation, without adjustment, measures both the adequacy of the forecasts and effectiveness of policies [57]. Issues are exacerbated (alleviated) for longer-(shorter-) term forecasts where model-based policy changes could be more (less) likely. Two ways to address these include: i) directly incorporating different policies with corresponding probabilities in the integration over all possible future states of the world, which requires modeller and decision-maker dialogue, or ii) using scenario projections for these quantities to consider “what-if” situations of future policy decisions [51]. Scenario projections, alongside our evaluation framework, may be more suitable when there is greater uncertainty about future policy changes and predictions are likely to directly alter the future that is predicted, while forecasts may be more suited to short-term horizons and targets that are largely invariant to interventions. This feedback loop is not always an issue for forecasting as targeted epidemiological quantities can be largely invariant to interventions (e.g., incident cases after adding to ICU capacity). All of this highlights the need for close collaboration with decision-makers to maintain clarity about the future policies, the future that a prediction is targeting, and the associated utility, limitations, and implications of the prediction model.

A reliable prediction does not imply effective, targeted action, which also requires understanding *why* or *how* transmission has occurred. Our framework should be used alongside inference of key epidemiological quantities, variant/serotype predictions, nowcasts of the present dynamics, analyses of intervention effectiveness, and many other tasks [52]. Beyond modelling, experts from other fields (e.g., health economics and social theory) should be embedded in multidisciplinary decision-making processes for an outbreak [43].

The applications of our framework have several limitations. We evaluated forecasts on imperfect case data without any data on policy decisions or reporting, thereby reflecting the realities of data collection, modelling, and human responses in outbreaks. Our predictability metric (PE) is robust to both observational and dynamical noise, while we focus on stable performance over 12-week windows and across space to limit sensitivity to surveillance artefacts. Epidemiological delays are also commonplace, impacting the forecasting, evaluation, and communication for decision-making [58]. This suggests using new methods for handling delays [59, 60]. Forecasters and evaluators will also benefit from i) extensions to DRO (Section I) and other predictability analyses, ii) further diagnostic checks (e.g., reliability diagrams [33]) iii) formal statistical testing [61], and iv) forecast post-processing techniques [62].

To conclude, our new framework shifts the perspective of forecast evaluation from the forecaster to the decision-maker. Recognising that there is no one-size-fits-all approach to forecasting, evaluation, or decision-making, we provide principled procedures for forecasters and evaluators to measure and improve the value of models for informing and empowering decision-makers.

## Data Availability

All code and data used in our analysis are available at https://github.com/cathalmills/forecast_evaluation.

## References

[1] Allan H. Murphy. “What Is a Good Forecast? An Essay on the Nature of Goodness in Weather Forecasting”. en. In: Weather and Forecasting 8.2 (June 1993), pp. 281–293. ISSN: 0882-8156, 1520-0434. DOI: 10.1175/1520-0434(1993)008<0281:WIAGFA>2.0.CO;2. URL: http://journals.ametsoc.org/doi/10.1175/1520-0434(1993)008%3C0281:WIAGFA%3E2.0.CO;2 (visited on 01/13/2025).

[2] Evan L. Ray et al. “Comparing trained and untrained probabilistic ensemble forecasts of COVID-19 cases and deaths in the United States”. en. In: International Journal of Forecasting 39.3 (July 2023), pp. 1366–1383. ISSN: 01692070. DOI: 10.1016/j.ijforecast.2022.06.005. URL: https://linkinghub.elsevier.com/retrieve/pii/S0169207022000966 (visited on 01/02/2024).

[3] Nikos I. Bosse et al. “Comparing human and model-based forecasts of COVID-19 in Germany and Poland”. en. In: PLOS Computational Biology 18.9 (Sept. 2022). Ed. by James M McCaw, e1010405. ISSN: 1553-7358. DOI: 10.1371/journal.pcbi.1010405. URL: https://dx.plos.org/10.1371/journal.pcbi.1010405 (visited on 10/14/2024).

[4] Sarabeth M. Mathis et al. “Evaluation of FluSight influenza forecasting in the 2021–22 and 2022–23 seasons with a new target laboratory-confirmed influenza hospitalizations”. en. In: Nature Communications 15.1 (July 2024), p. 6289. ISSN: 2041-1723. DOI: 10.1038/s41467-024-50601-9. URL: https://www.nature.com/articles/s41467-024-50601-9 (visited on 10/20/2024).

[5] Nikos I. Bosse et al. “Scoring epidemiological forecasts on transformed scales”. en. In: PLOS Computational Biology 19.8 (Aug. 2023). Ed. by James M McCaw, e1011393. ISSN: 1553-7358. DOI: 10.1371/journal.pcbi.1011393. URL: https://dx.plos.org/10.1371/journal.pcbi.1011393 (visited on 05/25/2024).

[6] Michael A. Johansson et al. “An open challenge to advance probabilistic forecasting for dengue epidemics”. en. In: Proceedings of the National Academy of Sciences 116.48 (Nov. 2019), pp. 24268–24274. ISSN: 0027-8424, 1091-6490. DOI: 10.1073/pnas.1909865116. URL: https://pnas.org/doi/full/10.1073/pnas.1909865116 (visited on 09/21/2023).

[7] Aaron Gerding et al. Evaluating infectious disease forecasts with allocation scoring rules. arXiv:2312.16201 [stat]. Mar. 2024. URL: http://arxiv.org/abs/2312.16201 (visited on 08/20/2024).

[8] Kelly R. Moran et al. “Epidemic Forecasting is Messier Than Weather Forecasting: The Role of Human Behavior and Internet Data Streams in Epidemic Forecast”. en. In: Journal of Infectious Diseases 214.suppl 4 (Dec. 2016), S404–S408. ISSN: 0022-1899, 1537-6613. DOI: 10.1093/infdis/jiw375. URL: https://academic.oup.com/jid/article-lookup/doi/10.1093/infdis/jiw375 (visited on 04/22/2025).

[9] Vanessa J. Fundel et al. “Promoting the use of probabilistic weather forecasts through a dialogue between scientists, developers and end-users”. en. In: Quarterly Journal of the Royal Meteorological Society 145.S1 (Sept. 2019), pp. 210–231. ISSN: 0035-9009, 1477-870X. DOI: 10.1002/qj.3482. URL: https://rmets.onlinelibrary.wiley.com/doi/10.1002/qj.3482 (visited on 06/09/2025).

[10] Martin Janousek Thomas Haiden. Evaluation of ECMWF forecasts, including the 2023 upgrade. en. text. 2023. URL: https://www.ecmwf.int/en/elibrary/81389-evaluation-ecmwf-forecasts-including-2023-upgrade (visited on 03/01/2025).

[11] Tilmann Gneiting and Adrian E Raftery. “Strictly Proper Scoring Rules, Prediction, and Estimation”. en. In: Journal of the American Statistical Association 102.477 (Mar. 2007), pp. 359–378. ISSN: 0162-1459, 1537-274X. DOI: 10.1198/016214506000001437. URL: http://www.tandfonline.com/doi/abs/10.1198/016214506000001437 (visited on 04/21/2025).

[12] Johannes Bracher et al. “Evaluating epidemic forecasts in an interval format”. en. In: PLOS Computational Biology 17.2 (Feb. 2021). Ed. by Virginia E. Pitzer, e1008618. ISSN: 1553-7358. DOI: 10.1371/journal.pcbi.1008618. URL: https://dx.plos.org/10.1371/journal.pcbi.1008618 (visited on 01/17/2024).

[13] Glenn W. Brier. “Verification of Forecasts Expressed in Terms of Probability”. In: Monthly Weather Review 78 (Jan. 1950). Publisher: AMS ADS Bibcode: 1950MWRv…78. 1B, p. 1. ISSN: 0027-0644. DOI: 10.1175/1520-0493(1950)078<0001:VOFEIT>2.0.CO;2. URL: https://ui.adsabs.harvard.edu/abs/1950MWRv…781B (visited on 02/15/2025).

[14] Estee Y Cramer et al. Evaluation of individual and ensemble probabilistic forecasts of COVID-19 mortality in the US. en. Feb. 2021. DOI: 10.1101/2021.02.03.21250974. URL: http://medrxiv.org/lookup/doi/10.1101/2021.02.03.21250974 (visited on 05/12/2025).

[15] Jonathon Mellor et al. “Forecasting COVID-19, influenza, and RSV hospitalizations over winter 2023–4 in England”. en. In: International Journal of Epidemiology 54.3 (Apr. 2025), dyaf066. ISSN: 0300-5771, 1464-3685. DOI: 10.1093/ije/dyaf066. URL: https://academic.oup.com/ije/article/doi/10.1093/ije/dyaf066/8156944 (visited on 06/08/2025).

[16] Velma K. Lopez et al. “Challenges of COVID-19 Case Forecasting in the US, 2020–2021”. en. In: PLOS Computational Biology 20.5 (May 2024). Ed. by Daniel B. Larremore, e1011200. ISSN: 1553-7358. DOI: 10.1371/journal.pcbi.1011200. URL: https://dx.plos.org/10.1371/journal.pcbi.1011200 (visited on 10/13/2024).

[17] Cathal Mills et al. “Multi-model approach to understand and predict past and future dengue epidemic dynamics”. en. In: Royal Society Open Science 12.11 (Nov. 2025), p. 241870. ISSN: 2054-5703. DOI: 10.1098/rsos.241870. URL: https://royalsocietypublishing.org/doi/10.1098/rsos.241870 (visited on 11/19/2025).

[18] Eduardo Correa Araujo et al. Leveraging probabilistic forecasts for dengue preparedness and control: the 2024 Dengue Forecasting Sprint in Brazil. en. May 2025. DOI: 10.1101/2025.05.12.25327419. URL: http://medrxiv.org/lookup/doi/10.1101/2025.05.12.25327419 (visited on 05/16/2025).

[19] Allan H. Murphy. “A New Vector Partition of the Probability Score”. en. In: Journal of Applied Meteorology 12.4 (June 1973), pp. 595–600. ISSN: 0021-8952. DOI: 10.1175/1520-0450(1973)012<0595:ANVPOT>2.0.CO;2. URL: http://journals.ametsoc.org/doi/10.1175/1520-0450(1973)012%3C0595:ANVPOT%3E2.0.CO;2 (visited on 04/03/2025).

[20] Hans Hersbach. “Decomposition of the Continuous Ranked Probability Score for Ensemble Prediction Systems”. en. In: Weather and Forecasting 15.5 (Oct. 2000), pp. 559–570. ISSN: 0882-8156, 1520-0434. DOI: 10.1175/1520-0434(2000)015<0559:DOTCRP>2.0.CO;2. URL: http://journals.ametsoc.org/doi/10.1175/1520-0434(2000)015%3C0559:DOTCRP%3E2.0.CO;2 (visited on 04/03/2025).

[21] G. Candille and O. Talagrand. “Evaluation of probabilistic prediction systems for a scalar variable”. en. In: Quarterly Journal of the Royal Meteorological Society 131.609 (July 2005), pp. 2131–2150. ISSN: 0035-9009, 1477-870X. DOI: 10.1256/qj.04.71. URL: https://rmets.onlinelibrary.wiley.com/doi/10.1256/qj.04.71 (visited on 04/03/2025).

[22] Meelis Kull and Peter Flach. “Novel Decompositions of Proper Scoring Rules for Classification: Score Adjustment as Precursor to Calibration”. en. In: Machine Learning and Knowledge Discovery in Databases. Ed. by Annalisa Appice et al. Vol. 9284. Series Title: Lecture Notes in Computer Science. Cham: Springer International Publishing, 2015, pp. 68–85. ISBN: 978-3-319-23527-1 978-3-319-23528-8. DOI: 10.1007/978-3-319-23528-8_5. URL: http://link.springer.com/10.1007/978-3-319-23528-8_5 (visited on 04/03/2025).

[23] Stefan Siegert. “Simplifying and generalising Murphy’s Brier score decomposition”. en. In: Quarterly Journal of the Royal Meteorological Society 143.703 (Jan. 2017), pp. 1178–1183. ISSN: 0035-9009, 1477-870X. DOI: 10.1002/qj.2985. URL: https://rmets.onlinelibrary.wiley.com/doi/10.1002/qj.2985 (visited on 04/03/2025).

[24] Timo Dimitriadis, Tilmann Gneiting, and Alexander I. Jordan. “Stable reliability diagrams for probabilistic classifiers”. en. In: Proceedings of the National Academy of Sciences 118.8 (Feb. 2021), e2016191118. ISSN: 0027-8424, 1091-6490. DOI: 10.1073/pnas.2016191118. URL: https://pnas.org/doi/full/10.1073/pnas.2016191118 (visited on 02/11/2025).

[25] Sebastian Arnold et al. “Decompositions of the mean continuous ranked probability score”. In: Electronic Journal of Statistics 18.2 (Jan. 2024). ISSN: 1935-7524. DOI: 10.1214/24-EJS2316. URL: https://projecteuclid.org/continuous-ranked-probability-score/10.1214/24-EJS2316.full (visited on 05/16/2025).

[26] Tilmann Gneiting et al. “Model Diagnostics and Forecast Evaluation for Quantiles”. en. In: Annual Review of Statistics and Its Application 10.1 (Mar. 2023), pp. 597–621. ISSN: 2326-8298, 2326-831X. DOI: 10.1146/annurev-statistics-032921-020240. URL: https://www.annualreviews.org/doi/10.1146/annurev-statistics-032921-020240 (visited on 02/02/2025).

[27] Werner Ehm et al. “Of Quantiles and Expectiles: Consistent Scoring Functions, Choquet Representations and Forecast Rankings”. en. In: Journal of the Royal Statistical Society Series B: Statistical Methodology 78.3 (June 2016), pp. 505–562. ISSN: 1369-7412, 1467-9868. DOI: 10.1111/rssb.12154. URL: https://academic.oup.com/jrsssb/article/78/3/505/7040984 (visited on 02/25/2025).

[28] Zied Ben Bouallègue, Thomas Haiden, and David S. Richardson. “The diagonal score: Definition, properties, and interpretations”. en. In: Quarterly Journal of the Royal Meteorological Society 144.714 (July 2018), pp. 1463–1473. ISSN: 0035-9009, 1477-870X. DOI: 10.1002/qj.3293. URL: https://rmets.onlinelibrary.wiley.com/doi/10.1002/qj.3293 (visited on 01/28/2025).

[29] Tim Palmer and David Richardson. “Decisions, decisions… !” In: (2014). Publisher: ECMWF. DOI: 10.21957/BYCHJ3CF. URL: https://www.ecmwf.int/node/17333 (visited on 01/25/2025).

[30] Thomas J. Kniesner and W. Kip Viscusi. “The Value of a Statistical Life”. en. In: Oxford Research Encyclopedia of Economics and Finance. July 2019. ISBN: 978-0-19-062597-9. DOI: 10.1093/acrefore/9780190625979.013.138. URL: https://oxfordre.com/economics/display/10.1093/acrefore/9780190625979.001.0001/acrefore-9780190625979-e-138 (visited on 05/22/2025).

[31] Christopher McCabe, Karl Claxton, and Anthony J. Culyer. “The NICE cost-effectiveness threshold: what it is and what that means”. eng. In: PharmacoEconomics 26.9 (2008), pp. 733–744. ISSN: 1170-7690. DOI: 10.2165/00019053-200826090-00004.

[32] Nicholas J. Irons and Adrian E. Raftery. US COVID-19 school closure was not cost-effective, but other measures were. arXiv:2411.12016 [stat]. Nov. 2024. DOI: 10.48550/arXiv.2411.12016. URL: http://arxiv.org/abs/2411.12016 (visited on 05/22/2025).

[33] Timo Dimitriadis et al. “Evaluating probabilistic classifiers: The triptych”. en. In: International Journal of Forecasting 40.3 (July 2024), pp. 1101–1122. ISSN: 01692070. DOI: 10.1016/j.ijforecast.2023.09.007. URL: http://linkinghub.elsevier.com/retrieve/pii/S0169207023000997 (visited on 01/26/2025).

[34] Ilan Price et al. “Probabilistic weather forecasting with machine learning”. en. In: Nature (Dec. 2024). ISSN: 0028-0836, 1476-4687. DOI: 10.1038/s41586-024-08252-9. URL: https://www.nature.com/articles/s41586-024-08252-9 (visited on 12/05/2024).

[35] David Richardson. “Skill and relative economic value of the ECMWF Ensemble Prediction System”. In: (1998). Publisher: ECMWF. DOI: 10.21957/2XJS554NR. URL: https://www.ecmwf.int/node/11903 (visited on 02/15/2025).

[36] Samuel V. Scarpino and Giovanni Petri. “On the predictability of infectious disease outbreaks”. en. In: Nature Communications 10.1 (Feb. 2019), p. 898. ISSN: 2041-1723. DOI: 10.1038/s41467-019-08616-0. URL: https://www.nature.com/articles/s41467-019-08616-0 (visited on 02/06/2025).

[37] C. E. Shannon. “A Mathematical Theory of Communication”. en. In: Bell System Technical Journal 27.3 (July 1948), pp. 379–423. ISSN: 00058580. DOI: 10.1002/j.1538-7305.1948.tb01338.x. URL: https://ieeexplore.ieee.org/document/6773024 (visited on 05/15/2025).

[38] Simon Pollett et al. “Recommended reporting items for epidemic forecasting and prediction research: The EPIFORGE 2020 guidelines”. en. In: PLOS Medicine 18.10 (Oct. 2021), e1003793. ISSN: 1549-1676. DOI: 10.1371/journal.pmed.1003793. URL: https://dx.plos.org/10.1371/journal.pmed.1003793 (visited on 06/13/2024).

[39] Estee Y. Cramer et al. “The United States COVID-19 Forecast Hub dataset”. en. In: Scientific Data 9.1 (Aug. 2022), p. 462. ISSN: 2052-4463. DOI: 10.1038/s41597-022-01517-w. URL: https://www.nature.com/articles/s41597-022-01517-w (visited on 03/27/2025).

[40] Estee Y. Cramer et al. “Evaluation of individual and ensemble probabilistic forecasts of COVID-19 mortality in the United States”. eng. In: Proceedings of the National Academy of Sciences of the United States of America 119.15 (Apr. 2022), e2113561119. ISSN: 1091-6490. DOI: 10.1073/pnas.2113561119.

[41] Charles Blackorby, Walter Bossert, and David J. Donaldson. Population Issues in Social Choice Theory, Welfare Economics, and Ethics. 1st ed. Cambridge University Press, Aug. 2005. ISBN: 978-0-521-53258-7 978-0-521-82551-1 978-1-139-05224-5. DOI: 10.1017/CCOL0521825512. URL: https://www.cambridge.org/core/product/identifier/9781139052245/type/book (visited on 02/24/2025).

[42] Kenneth J. Arrow. “A Difficulty in the Concept of Social Welfare”. en. In: Journal of Political Economy 58.4 (Aug. 1950), pp. 328–346. ISSN: 0022-3808, 1537-534X. DOI: 10.1086/256963. URL: https://www.journals.uchicago.edu/doi/10.1086/256963 (visited on 02/24/2025).

[43] Loïc Berger et al. “Rational policymaking during a pandemic”. en. In: Proceedings of the National Academy of Sciences 118.4 (Jan. 2021), e2012704118. ISSN: 0027-8424, 1091-6490. DOI: 10.1073/pnas.2012704118. URL: https://pnas.org/doi/full/10.1073/pnas.2012704118 (visited on 04/03/2025).

[44] Oliver Morgan. “How decision makers can use quantitative approaches to guide outbreak responses”. eng. In: Philosophical Transactions of the Royal Society of London. Series B, Biological Sciences 374.1776 (July 2019), p. 20180365. ISSN: 1471-2970. DOI: 10.1098/rstb.2018.0365.

[45] Cabinet Office. Independent review into the response to the 2009 swine flu pandemic. Tech. rep. Cabinet Office, July 2010.

[46] Graham F. Medley. “A consensus of evidence: The role of SPI-M-O in the UK COVID-19 response”. In: Advances in Biological Regulation. COVID-19 models and expectations 86 (Dec. 2022), p. 100918. ISSN: 2212-4926. DOI: 10.1016/j.jbior.2022.100918. URL: https://www.sciencedirect.com/science/article/pii/S2212492622000586 (visited on 03/03/2025).

[47] Rob J. Hyndman and George Athanasopoulos. Forecasting: principles and practice. eng. Third print edition. Melbourne, Australia: Otexts, Online Open-Access Textbooks, 2021. ISBN: 978-0-9875071-3-6.

[48] Paul-Christian Bürkner, Jonah Gabry, and Aki Vehtari. “Approximate leave-future-out cross-validation for Bayesian time series models”. en. In: Journal of Statistical Computation and Simulation 90.14 (Sept. 2020), pp. 2499–2523. ISSN: 0094-9655, 1563-5163. DOI: 10.1080/00949655.2020.1783262. URL: https://www.tandfonline.com/doi/full/10.1080/00949655.2020.1783262 (visited on 05/31/2025).

[49] Tilmann Gneiting and Johannes Resin. “Regression diagnostics meets forecast evaluation: conditional calibration, reliability diagrams, and coefficient of determination”. In: Electronic Journal of Statistics 17.2 (Jan. 2023). ISSN: 1935-7524. DOI: 10.1214/23-EJS2180. URL: https://projecteuclid.org/journals/electronic-journal-of-statistics/volume-17/issue-2/Regression-diagnostics-meets-forecast-evaluation--conditional-calibration-reliability-diagrams/10.1214/23-EJS2180.full (visited on 02/15/2025).

[50] Ruth McCabe et al. “Communicating uncertainty in epidemic models”. en. In: Epidemics 37 (Dec. 2021), p. 100520. ISSN: 17554365. DOI: 10.1016/j.epidem.2021.100520. URL: https://linkinghub.elsevier.com/retrieve/pii/S1755436521000669 (visited on 08/18/2024).

[51] Michael C. Runge et al. “Scenario design for infectious disease projections: Integrating concepts from decision analysis and experimental design”. en. In: Epidemics 47 (June 2024), p. 100775. ISSN: 17554365. DOI: 10.1016/j.epidem.2024.100775. URL: https://linkinghub.elsevier.com/retrieve/pii/S1755436524000367 (visited on 02/12/2025).

[52] Anne Cori and Adam Kucharski. “Inference of epidemic dynamics in the COVID-19 era and beyond”. en. In: Epidemics 48 (Sept. 2024), p. 100784. ISSN: 17554365. DOI: 10.1016/j.epidem.2024.100784. URL: https://linkinghub.elsevier.com/retrieve/pii/S1755436524000458 (visited on 10/09/2024).

[53] Grace Liu et al. “Binary climate data visuals amplify perceived impact of climate change”. en. In: Nature Human Behaviour (Apr. 2025). ISSN: 2397-3374. DOI: 10.1038/s41562-025-02183-9. URL: https://www.nature.com/articles/s41562-025-02183-9 (visited on 05/12/2025).

[54] Claudia R. Schneider et al. “The effects of communicating scientific uncertainty on trust and decision making in a public health context”. en. In: Judgment and Decision Making 17.4 (July 2022), pp. 849–882. ISSN: 1930-2975. DOI: 10.1017/S1930297500008962. URL: https://www.cambridge.org/core/product/identifier/S1930297500008962/type/journal_article (visited on 05/02/2025).

[55] Dung Phung et al. “Advancing adoptability and sustainability of digital prediction tools for climate-sensitive infectious disease prevention and control”. en. In: Nature Communications 16.1 (Feb. 2025), p. 1644. ISSN: 2041-1723. DOI: 10.1038/s41467-025-56826-6. URL: https://www.nature.com/articles/s41467-025-56826-6 (visited on 02/17/2025).

[56] Manit Sittimart et al. “An overview of the perspectives used in health economic evaluations”. en. In: Cost Effectiveness and Resource Allocation 22.1 (May 2024), p. 41. ISSN: 1478-7547. DOI: 10.1186/s12962-024-00552-1. URL: https://resource-allocation.biomedcentral.com/articles/10.1186/s12962-024-00552-1 (visited on 01/28/2025).

[57] Rob J. Hyndman. “Forecasting, causality and feedback”. en. In: International Journal of Forecasting 39.2 (Apr. 2023), pp. 558–560. ISSN: 01692070. DOI: 10.1016/j.ijforecast.2022.09.007. URL: https://linkinghub.elsevier.com/retrieve/pii/S0169207022001388. (visited on 01/29/2025).

[58] Christopher J. M. Whitty. “What makes an academic paper useful for health policy?” In: BMC Medicine 13.1 (Dec. 2015), p. 301. ISSN: 1741-7015. DOI: 10.1186/s12916-015-0544-8. URL: https://doi.org/10.1186/s12916-015-0544-8 (visited on 04/11/2025).

[59] Kelly Charniga et al. Best practices for estimating and reporting epidemiological delay distributions of infectious diseases using public health surveillance and healthcare data. Version Number: 1. 2024. DOI: 10.48550/ARXIV.2405.08841. URL: https://arxiv.org/abs/2405.08841 (visited on 07/01/2024).

[60] Sang Woo Park et al. Estimating epidemiological delay distributions for infectious diseases. en. Jan. 2024. DOI: 10.1101/2024.01.12.24301247. URL: http://medrxiv.org/lookup/doi/10.1101/2024.01.12.24301247 (visited on 05/15/2024).

[61] Raffaella Giacomini and Ivana Komunjer. “Evaluation and Combination of Conditional Quantile Forecasts”. In: Journal of Business & Economic Statistics 23.4 (Oct. 2005). Publisher: ASA Website _eprint: https://doi.org/10.1198/073500105000000018, xpp. 416–431. ISSN: 0735-0015. DOI: 10.1198/073500105000000018. URL: https://doi.org/10.1198/073500105000000018 (visited on 06/03/2025).

[62] Anastasios N. Angelopoulos, Emmanuel J. Candes, and Ryan J. Tibshirani. Conformal PID Control for Time Series Prediction. arXiv:2307.16895 [cs, eess, stat]. July 2023. URL: http://arxiv.org/abs/2307.16895 (visited on 08/23/2024).

[63] Tilmann Gneiting and Peter Vogel. “Receiver operating characteristic (ROC) curves: equivalences, beta model, and minimum distance estimation”. en. In: Machine Learning 111.6 (June 2022), pp. 2147–2159. ISSN: 0885-6125, 1573-0565. DOI: 10.1007/s10994-021-06115-2. URL: https://link.springer.com/10.1007/s10994-021-06115-2 (visited on 03/28/2025).

[64] Nikos I. Bosse et al. Evaluating Forecasts with scoringutils in R. arXiv:2205.07090 [stat]. Nov. 2024. DOI: 10.48550/arXiv.2205.07090. URL: http://arxiv.org/abs/2205.07090 (visited on 04/14/2025).

[65] Alexander Jordan, Fabian Krüger, and Sebastian Lerch. “Evaluating Probabilistic Forecasts with scoringRules”. en. In: Journal of Statistical Software 90.12 (2019). ISSN: 1548-7660. DOI: 10.18637/jss.v090.i12. URL: http://www.jstatsoft.org/v90/i12/ (visited on 01/04/2024).

[66] Timo Dimitriadis and Alexander I. Jordan. triptych: Diagnostic Graphics to Evaluate Forecast Performance. en. Institution: Comprehensive R Archive Network Pages: 0.1.3. Oct. 2023. DOI: 10.32614/CRAN.package.triptych. URL: https://CRAN.R-project.org/package=triptych (visited on 06/07/2025).

[67] R Core Team. R: A Language and Environment for Statistical Computing. Vienna, Austria: R Foundation for Statistical Computing, 2025. URL: https://www.r-project.org/.

[68] Richard W. Katz and Jeffrey K. Lazo. “Economic Value of Weather and Climate Forecasts”. In: The Oxford Handbook of Economic Forecasting. Ed. by Michael P. Clements and David F. Hendry. Oxford University Press, July 2011, p. 0. ISBN: 978-0-19-539864-9. DOI: 10.1093/oxfordhb/9780195398649.013.0021. URL: https://doi.org/10.1093/oxfordhb/9780195398649.013.0021 (visited on 05/21/2025).

[69] R Tyrrell Rockafellar, Stanislav Uryasev, et al. “Optimization of conditional value-at-risk”. In: Journal of risk 2 (2000). Publisher: Citeseer, pp. 21–42.

[70] Hamed Rahimian and Sanjay Mehrotra. “Frameworks and results in distributionally robust optimization”. In: Open Journal of Mathematical Optimization 3 (2022), pp. 1–85.

[71] Tilmann Gneiting et al. “Calibrated Probabilistic Forecasting Using Ensemble Model Output Statistics and Minimum CRPS Estimation”. en. In: Monthly Weather Review 133.5 (May 2005), pp. 1098–1118. ISSN: 1520-0493, 0027-0644. DOI: 10.1175/MWR2904.1. URL: http://journals.ametsoc.org/doi/10.1175/MWR2904.1 (visited on 09/03/2024).

[72] Tobias Fissler and Johanna F. Ziegel. “Higher order elicitability and Osband’s principle”. In: The Annals of Statistics 44.4 (2016). Publisher: Institute of Mathematical Statistics, pp. 1680–1707. DOI: 10.1214/16-AOS1439. URL: https://doi.org/10.1214/16-AOS1439.

