## Supplementary material for "From metric to action: The decision value of infectious disease forecasts"

### A Table of framework concepts

Table SI 1: **Key concepts for our framework of probabilistic infectious disease forecasting:** We define important concepts discussed throughout the text and used in our framework for forecasting infectious diseases. Given the wealth of metrics and tools above, the choice of metric/tool may not be immediately obvious, and may be both user- and event-specific, which motivates our workflow (Figure 1 A) starting with the well-defined policy question(s) for a user/decision-maker and corresponding epidemic outcome to measure.

| Concept | Meaning |
|---|---|
| Brier Score (BS) | A strictly proper scoring rule defined as the mean squared error for a probabilistic forecast of a binary outcome [13]. |
| Classifiers | A probabilistic classifier produces a probability forecast for a binary outcome. |
| Consistent | A scoring function is consistent for a given functional $T(P)$ (e.g., quantile level) if the score is minimised when the forecast aligns with $T(P)$ consistent. |
| Continuous Ranked Probability Score (CRPS) | A proper scoring rule that is strict when the first moment of the forecast distribution is finite, and generalises mean absolute error for probabilistic forecasts of continuous outcomes [11]. |
| Cost-loss (C/L) ratio | In this decision model for binary outcome events, C/L ratio $\alpha$ is the cost (C) of preparation for an adverse event, divided by the avoidable loss (L) if the event occurs without any preparation [1, 35]. The optimal decision is to act/prepare if the forecasted probability exceeds this *decision threshold* C/L. Knowing plausible values of the ratio for a given binary event is sufficient for studying forecast model results, and thus for metric-based decision-making. |
| Discrimination (DSC) | A non-negative component of the decomposition for the mean value of a score, and measures the ability of a model's forecasts to distinguish events versus non-events [24]. Higher values indicate superior discrimination ability. |
| Event threshold | A cut-off value $\theta$ that is used for defining whether a binary outcome occurs or not. |
| Forecast/predictive distribution | A probabilistic forecast's distribution for the future value of a given quantity, often represented by a parametric distribution, samples from the distribution, and/or quantiles of the distribution. |
| Forecast horizon | The lead time at which forecasts are made. |
| Forecast quality | Measure of the alignment of a model's forecasted values with observed values. |
| Forecast value | Measure of the ability of a model's forecasted values to inform decision-making. |
| Horizon | The lead time at which a forecast for a future value is produced. |
| Miscalibration (MSC) | A non-negative component of the decomposition for the mean value of a score, and measures the calibration of a model's forecasts [24]. Lower values indicate superior calibration. |
| Murphy curve/diagram | For a given model, a curve of mean elementary scores (eq. (3.1)) across different C/L ratios ($\alpha$) or event thresholds ($\theta$). Lower curves indicate better performance, and a Murphy diagram compares Murphy curves for different models. |
| Pinball loss | Measure of forecast performance for a specific user-defined probability/quantile level. Pinball loss is also known as Quantile Score (QS) or the asymmetric piecewise linear scoring function. |
| Permutation entropy (PE) | Measure of predictability which works by studying temporal dependencies in both linear and nonlinear time series. PE is a model-free measure that uses Shannon's Entropy to study the frequency of distinct patterns in time series data. |
| Predictability | The inherent randomness of a system. |
| Proper | A scoring rule is proper if the expected score is minimised by using the true probability distribution. |
| Scoring function | A function that assigns a score to a given functional (e.g., quantile level). This is not necessarily proper, and can asymmetrically penalise probabilistic forecasts (e.g., pinball loss). Lower values for a scoring function imply better predictive performance. |
| Scoring rule | A scoring rule (e.g., CRPS) is a specific type of scoring function (see above) that assigns a score to a probabilistic forecast, and lower values for a scoring rule imply better predictive performance. |

### B Relative Economic Value (REV)

The REV curve for a model is based on the C/L model presented in Section 3. We present the C/L matrix again to aid reader interpretation:

| | **Event occurs** | **Event does not occur** |
|---|---|---|
| **Action taken** | $C$ | $C$ |
| **Action not taken** | $L$ | 0 |

Suppose $\theta \in \mathbb{R}$ is a fixed event threshold (e.g., number of cases). Binary events ($y = 1$ if $> \theta$) are predicted to occur if the model forecasts the event with probability greater than or equal to the user-defined C/L ratio $\alpha$. This is equivalent to a forecast at quantile level 1-$\alpha$ being greater than or equal to $\theta$.

Then, the expected *expense* of the model ($E_{\text{model}}$) is derived from the confusion matrix for the model's classifications, incurring costs $C$ for observations where the model correctly predicted an event that occurs (a true positive rate, TP) or where the model incorrectly predicted an event that does not occur (a false positive rate, FP). On the other hand, events where the model failed to predict the event (false negative rate, FN) will incur the preventable loss $L$, while a true negative will incur no cost or loss. Mathematically, we write this as:

$$E_{\text{model}} = (\text{TP} + \text{FP}) \cdot C + \text{FN} \cdot L, \tag{B.1}$$

while a perfect (or oracle) probability model incurs expense only when the model predicts the event (which always coincides when the event occurs

$$E_{\text{perfect}} = \text{TP} \cdot C, \tag{B.2}$$

while a baseline (or reference) model is derived analogously to the studied model – using the baseline model's confusion matrix

$$E_{\text{baseline}} = (\text{TP}_{\text{baseline}} + \text{FP}_{\text{baseline}}) \cdot C + \text{FN}_{\text{baseline}} \cdot L. \tag{B.3}$$

Then, REV can be defined as:

$$\text{REV}(\alpha = C/L, \theta) := \frac{E_{\text{baseline}} - E_{\text{model}}}{E_{\text{baseline}} - E_{\text{perfect}}}. \tag{B.4}$$

For calculation, we see that REV is:

$$\text{REV}(\alpha = C/L, \theta) := \frac{(\text{TP}_{\text{baseline}} + \text{FP}_{\text{baseline}}) \cdot C + \text{FN}_{\text{baseline}} \cdot L - (\text{TP} + \text{FP}) \cdot C + \text{FN} \cdot L}{(\text{TP}_{\text{baseline}} + \text{FP}_{\text{baseline}}) \cdot C + \text{FN}_{\text{baseline}} \cdot L - \text{TP} \cdot C}, \tag{B.5}$$

where dividing through by loss $L$, allows us to compute REV when we only know the C/L ratio $\alpha$.

### C Other measures of forecast value

We briefly highlight other metrics that focus forecast evaluation on specific decision-makers and their priorities. Originating from weather forecasting [28], the diagonal score (Figure 2 B) assumes a fixed relationship between C/L $\alpha$ and the empirical probability of epidemic events $\pi(\theta)$ (i.e., how often different events occur). This makes it suitable for risk-intolerant decision-makers, as if the cost of action ($C$) remains fixed across event thresholds, rare extreme large event thresholds are associated with large losses ($L$).

Traditional classification tools include Receiver Operating Characteristic (ROC) curves and the Area Under the ROC Curve (AUC). As ROC curves can fail to satisfy concavity requirements and AUC ignores miscalibration [33, 63], we recommend the use of DSC-MCB properties in the CORP-based decompositions (e.g., of BS) and/or concave ROC curves that use the recalibrated forecasts of the CORP decomposition [33].

### D Extending predictability analyses

A key limitation of using eq. (5.2) is its univariate nature. An epidemic may be more predictable conditional on knowing other information, often provided by heterogeneous data streams. To quantify historical drivers of (un)predictability, we propose analyses across space and time. For example, $\chi$ could be estimated alongside $R(t)$ to understand the association between predictability and stages of epidemic growth. By further associating results with those for the forecast metrics of the previous sections, these retrospective analyses can enhance our understanding of the reasons behind historical forecast performance, and this understanding can then be used for anticipating future predictability and future performance.

### E EPIFORGE 2020 guidelines

Table SI 2: **EPIFORGE 2020 guidelines:** We present the pages which correspond to the checklist items of the EPIFORGE 2020 guidelines for forecast reporting [38].

| Section of Manuscript | # | Checklist Item | Reported on Page |
|---|---|---|---|
| Title/Abstract | 1 | Describe the study as forecast or prediction research in at least the title or abstract. | 1 |
| Introduction | 2 | Define the purpose of study and forecasting targets. | 1 and 10 |
| Methods | 3 | Fully document the methods. | 2-10 and SI |
| Methods | 4 | Identify whether the forecast was performed prospectively, in real time, and/or retrospectively. | 12 |
| Methods | 5 | Explicitly describe the origin of input source data, with references. | 10 |
| Methods | 6 | Provide source data with publication, or document reasons as to why this was not possible. | 10 |
| Methods | 7 | Describe input data processing procedures in detail. | 10 and SI |
| Methods | 8 | State and describe the model type, and document model assumptions, including references. | Table SI 2 |
| Methods | 9 | Make the model code available or document the reasons why this is not possible. | 10 |
| Methods | 10 | Describe the model validation and justify the approach. | 2-10 |
| Methods | 11 | Describe the forecast accuracy evaluation method used, with justification. | 2-10 |
| Methods | 12 | Where possible, compare results to a benchmark or other comparator model, with justification of comparator choice. | 10-11 |
| Methods | 13 | Describe the forecast horizon, with justification of its length. | 10-11 |
| Results | 14 | Present and explain uncertainty of forecasting results. | 10-11 |
| Results | 15 | Briefly summarize the results in nontechnical terms, including a non-technical interpretation of forecast uncertainty. | 10-12 |
| Results | 16 | If results are published as a data object, encourage a time-stamped version number. | 10-11 |
| Discussion | 17 | Describe the weaknesses of the forecast, including weaknesses specific to data quality and methods. | 10 and 12 |
| Discussion | 18 | If the forecast research is applicable to a specific epidemic, comment on its potential implications and impact for public health action and decision-making. | 11-12, 22 |
| Discussion | 19 | If the forecast research is applicable to a specific epidemic, comment on how generalizable it may be across populations. | 11-12 |

### F COVID-19 Forecast Hub Application

The COVID-19 Forecast Hub was the host for a collaborative forecasting effort in the United States [39]. Teams were invited to submit probabilistic forecasts for weekly incident COVID-19 cases and deaths at forecast horizons of one to four weeks. We focused on the forecasting of COVID-19 cases in this work, where teams submitted forecasts using seven quantile levels (0.025, 0.1, 0.25, 0.5, 0.75, 0.9, 0.975). We used the forecast data provided by [16].

For forecast evaluation in this work, we used the scoringutils, scoringrules, and triptych packages [33, 64–66] in R version 4.4.3 [67].

Our objective was not to repeat previous analyses, but rather to highlight i) how different metrics capture forecast value for informing simple policy questions, and ii) the implications of the statistical theory for different decision-makers and epidemic stages. There will be no one-size-fits-all approach for forecasting, evaluation, or decision-making, and any application should make clear the subjective choices made. Setting-specific considerations for the decision-maker include relevant policy questions, intervention action sets, costs and resources, and likely responses from the public. For the forecaster and evaluator, considerations for model development and selection include evaluation window sizes and spatial scales, both of which depend on practical data availability and requirements of public health authorities.

Table SI 3: **Submitted model names and descriptions:** The following table includes our abbreviation, the original model name from each modelling team's submission, and the corresponding model description provided in [16].

| Model name | Original name | Description |
|---|---|---|
| Ensemble | COVIDhub-4_week_ensemble | Unweighted average or median of submitted forecasts to the COVID-19 Forecast Hub |
| Baseline | COVIDhub-baseline | Median prediction at all future horizons is equal to the most recent observed incidence |
| CU-Select | CU-Select | A metapopulation county-level SEIR model for projecting future COVID-19 incidence cases and deaths |
| Karlen-pypm | Karlen-pypm | Finite time difference equations implemented as a general-purpose population modelling framework |
| CA-DELPHI | COVIDAnalytics-DELPHI | SEIR model augmented with underdetection and interventions |
| RW-ESG | RobertWalraven-ESG | Multiple skewed Gaussian mathematical fit |

### G Additional figures

#### G.1 Weighted Interval Score (WIS) across space and time

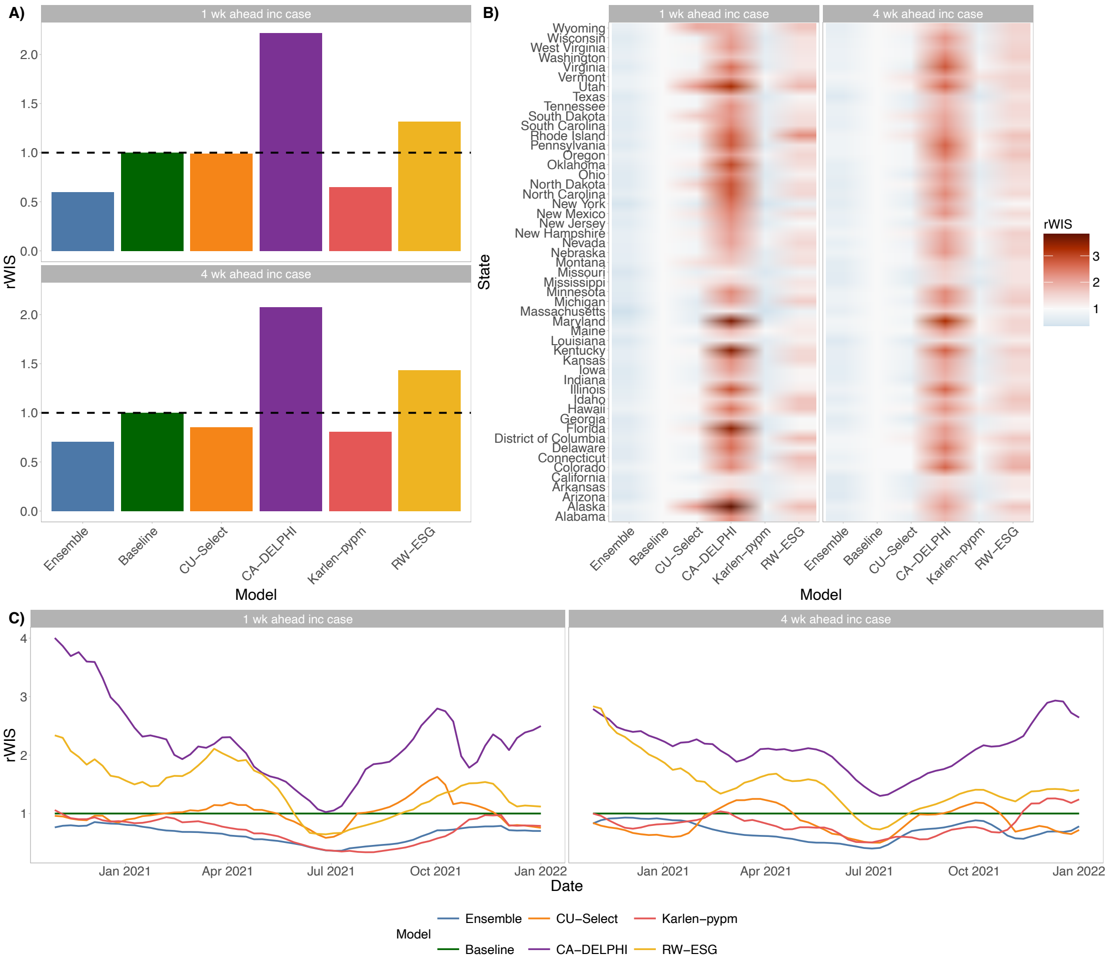


Figure SI 1: **Overall forecast quality across time and space:** Results for application of the relative Weighted Interval Score (rWIS) to forecasts of incident weekly COVID-19 cases on a logarithmic scale at one- and four-week horizons. rWIS is a scaled measure of relative performance (relative to the baseline model) that accounts for potentially different numbers of forecasts for different locations and time points. Each plot panel visualises mean rWIS **A)** across all locations, **B)** for specific locations, and **C)** in trailing windows across all locations.

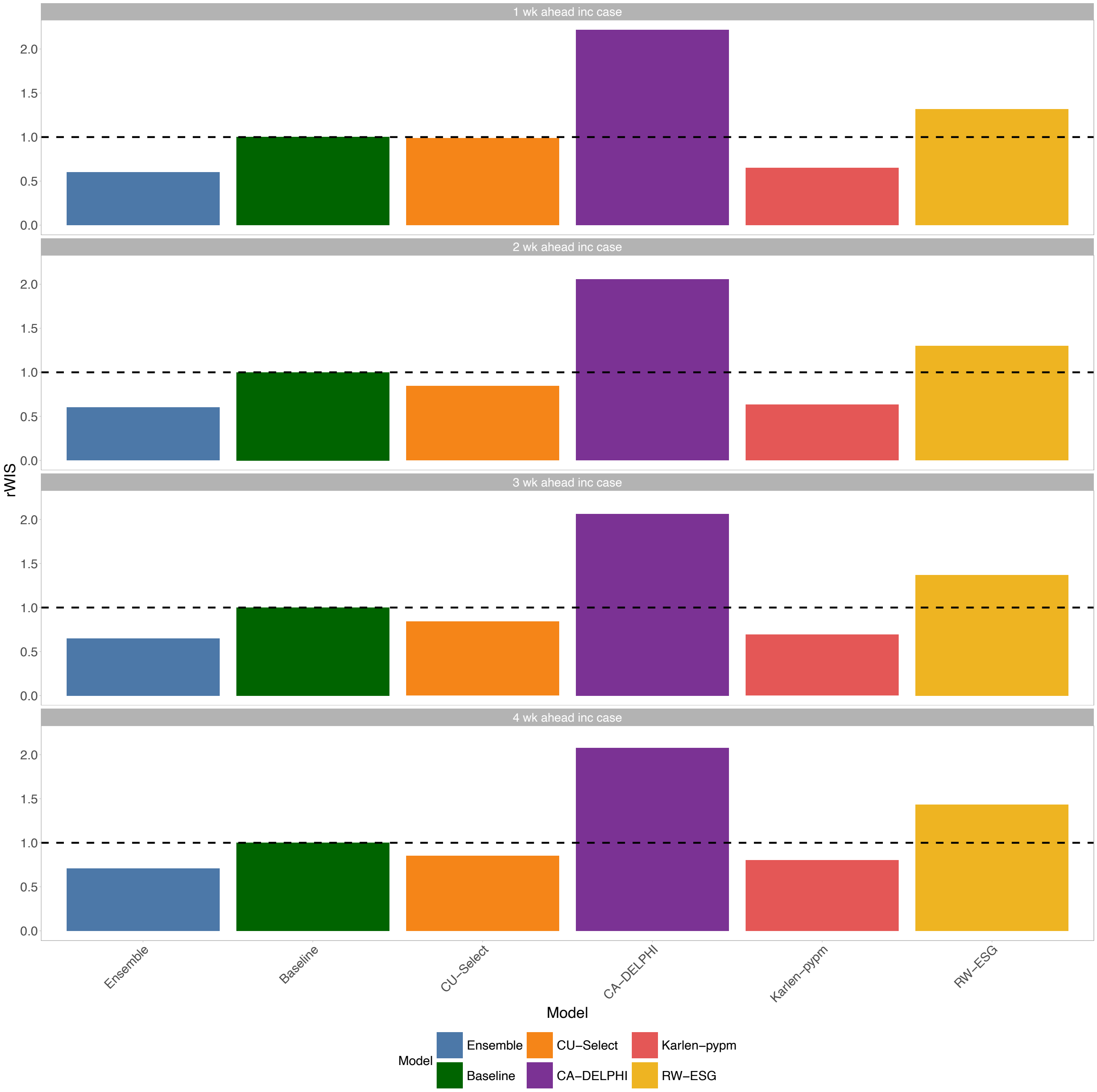


Figure SI 2: **Relative Weighted Interval Score (WIS) by different forecast horizons across all locations:** Results for application of the Weighted Interval Score (WIS) to forecasts of incident weekly COVID-19 cases (on a logarithmic scale) at one- to four-week horizons.

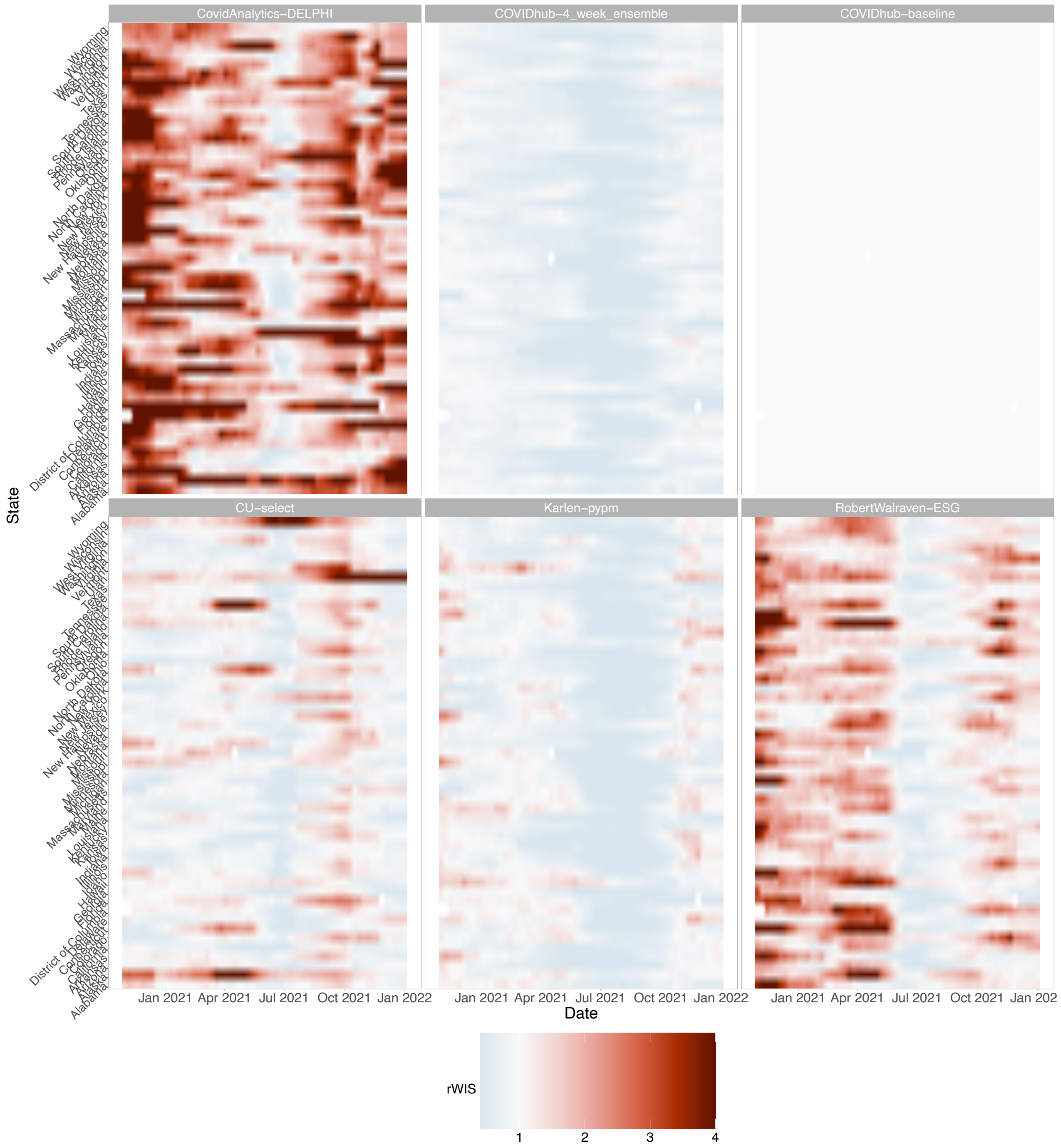


Figure SI 3: **Relative Weighted Interval Score (rWIS) for one-week-ahead forecasts by model and location in 12-week trailing windows:** Scaled relative Weighted Interval Score [12] for different models is visualised for each state (y-axis) and each trailing window where the x-axis denotes the maximum date of the 12-week trailing window. We calculated rWIS relative to the baseline model, and rWIS > 1 indicates greater forecast value for the uniform family of decision-makers in the given location and trailing window.

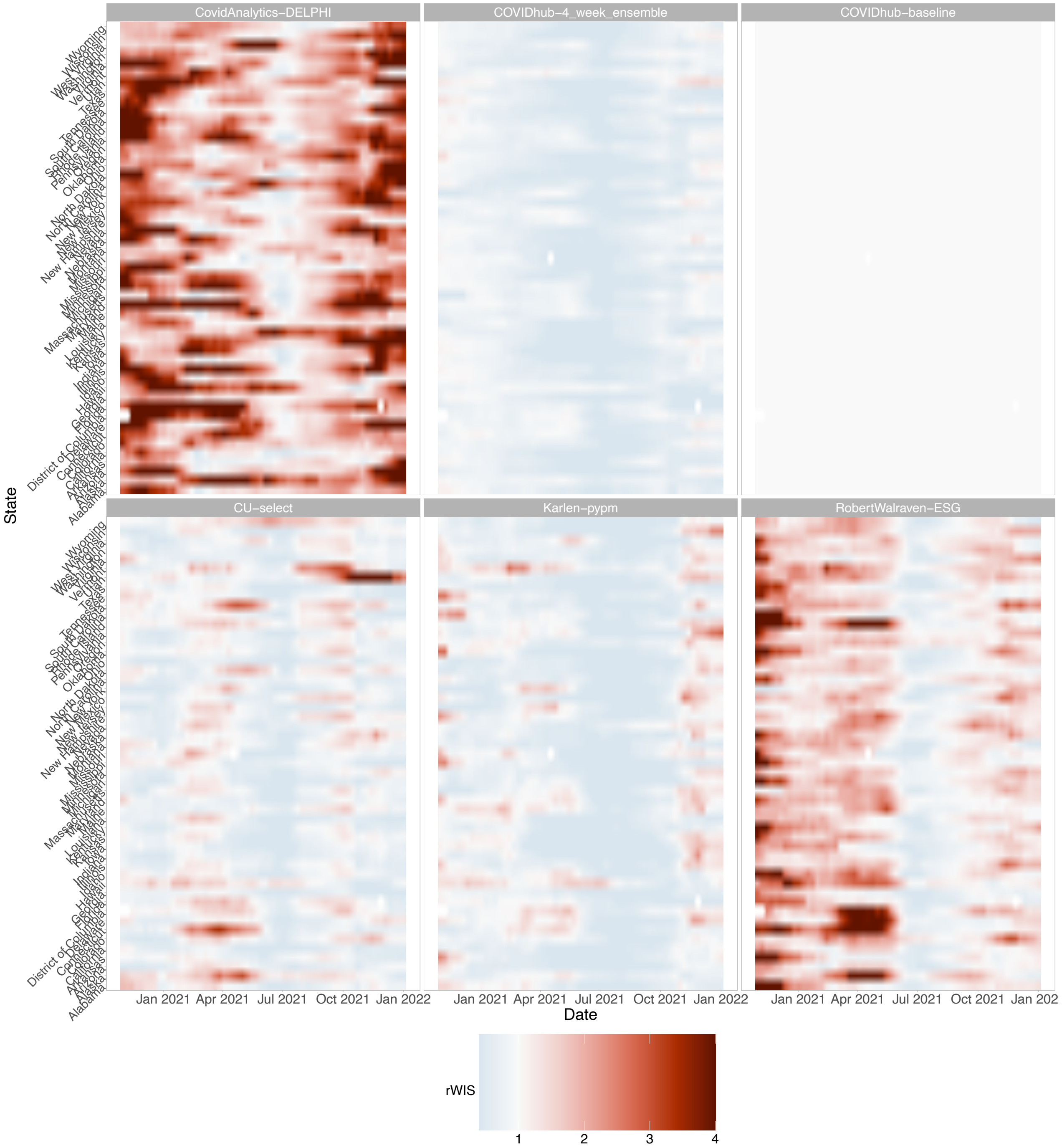


Figure SI 4: **Relative Weighted Interval Score (rWIS) for two-weeks-ahead forecasts by model and location in 12-week trailing windows:** Scaled relative Weighted Interval Score [12] for different models is visualised for each state (y-axis) and each trailing window where the x-axis denotes the maximum date of the 12-week trailing window. We calculated rWIS relative to the baseline model, and rWIS > 1 indicates greater forecast value for the uniform family of decision-makers in the given location and trailing window.

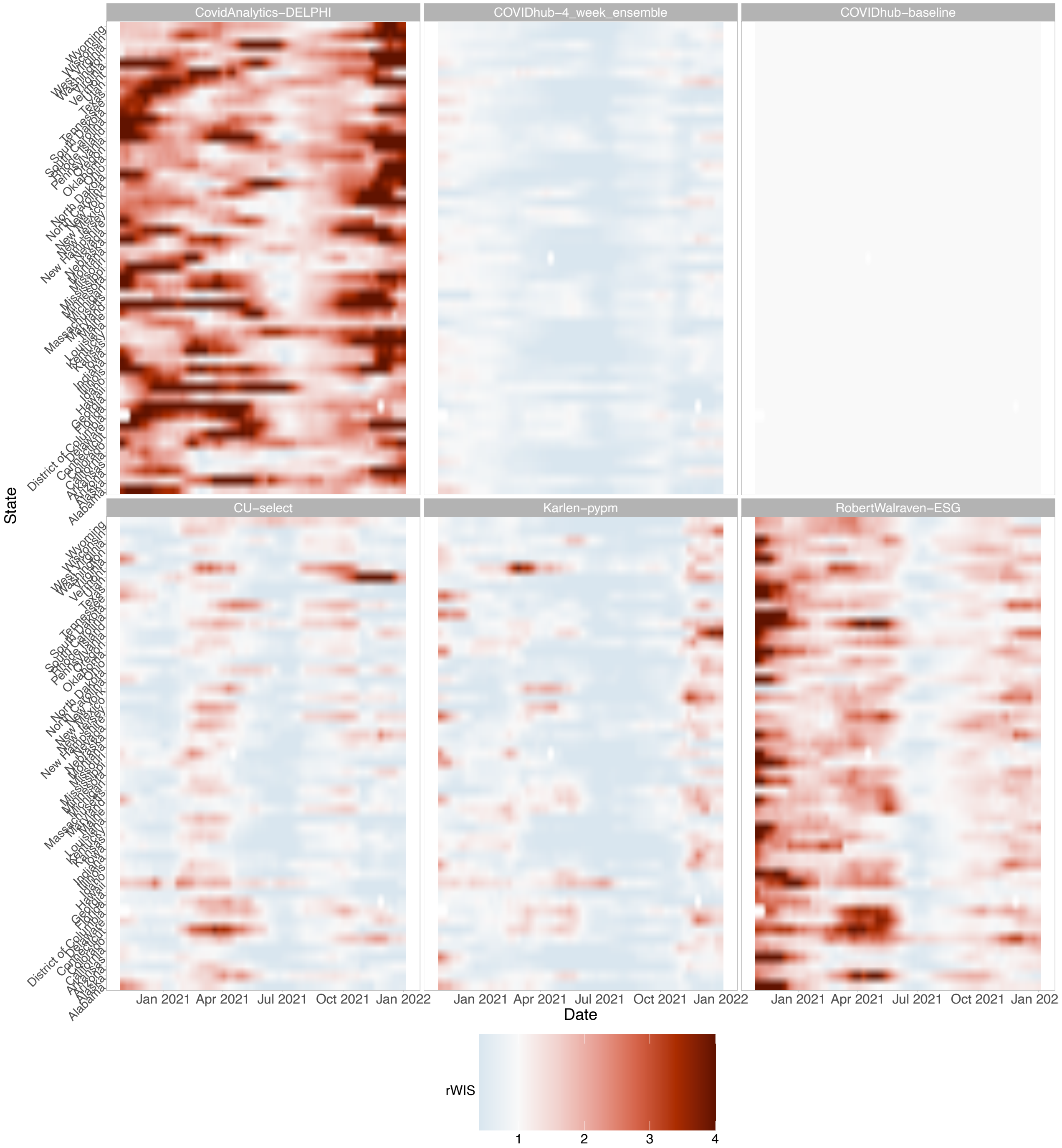


Figure SI 5: **Relative Weighted Interval Score (rWIS) for three-weeks-ahead forecasts by model and location in 12-week trailing windows:** Scaled relative Weighted Interval Score [12] for different models is visualised for each state (y-axis) and each trailing window where the x-axis denotes the maximum date of the 12-week trailing window. We calculated rWIS relative to the baseline model, and rWIS > 1 indicates greater forecast value for the uniform family of decision-makers in the given location and trailing window.

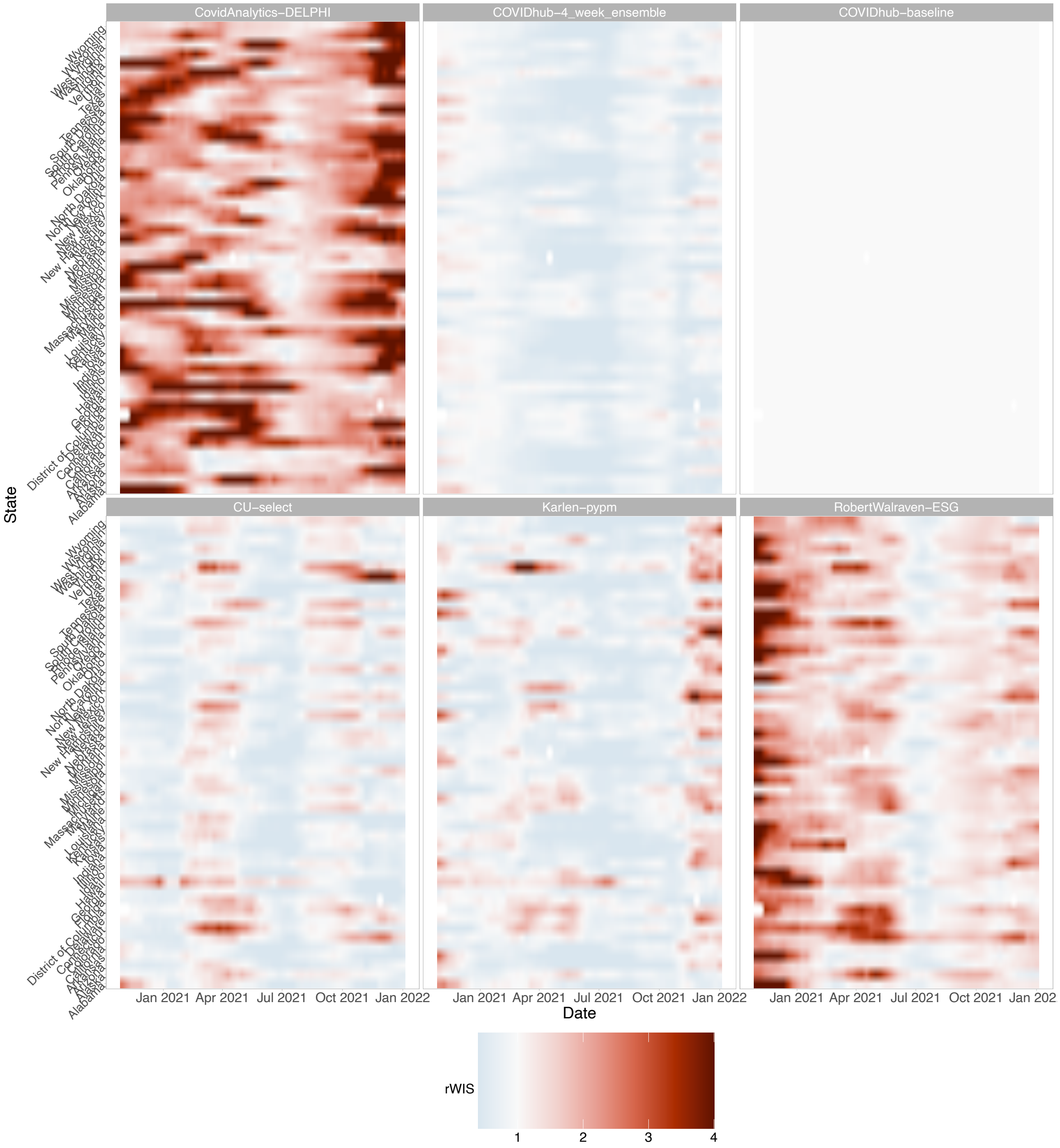


Figure SI 6: **Relative Weighted Interval Score (rWIS) for four-weeks-ahead forecasts by model and location in 12-week trailing windows:** Scaled relative Weighted Interval Score [12] for different models is visualised for each state (y-axis) and each trailing window where the x-axis denotes the maximum date of the 12-week trailing window. We calculated rWIS relative to the baseline model, and rWIS $> 1$ indicates greater forecast value for the uniform family of decision-makers in the given location and trailing window.

#### G.2 Relative Economic Value (REV) – Differing events and reference models

##### G.2.1 Upper 10% of all events

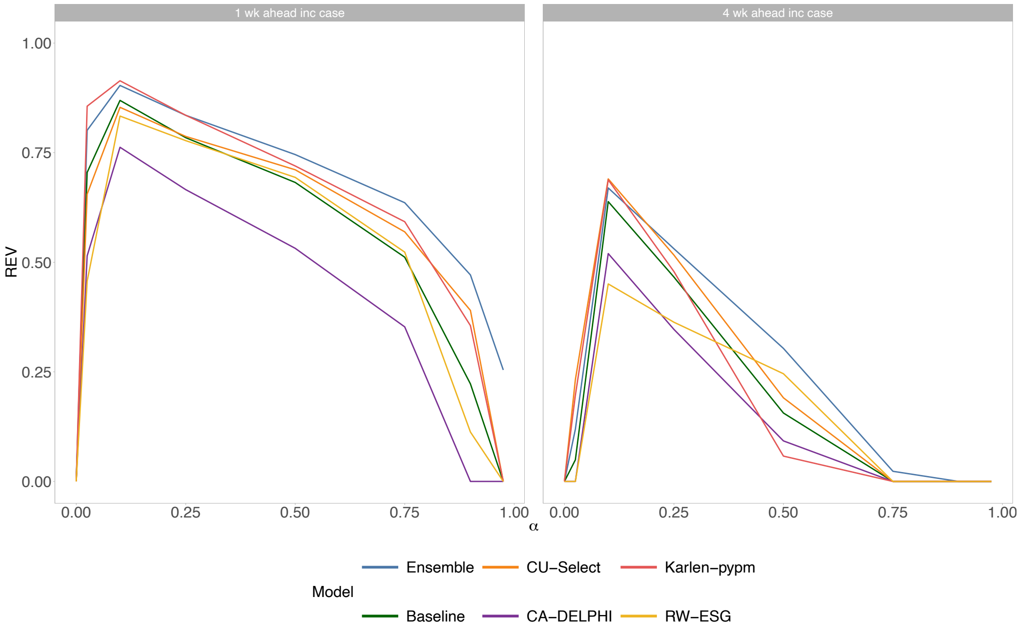


Figure SI 7: **Relative Economic Value (REV) for upper 10% of *all* observed cases:** REV for the different forecasting models, compared to a forecasting model which forecasts the binary event with probability of 10% (i.e., analogous to the climatology model used in weather forecasting).

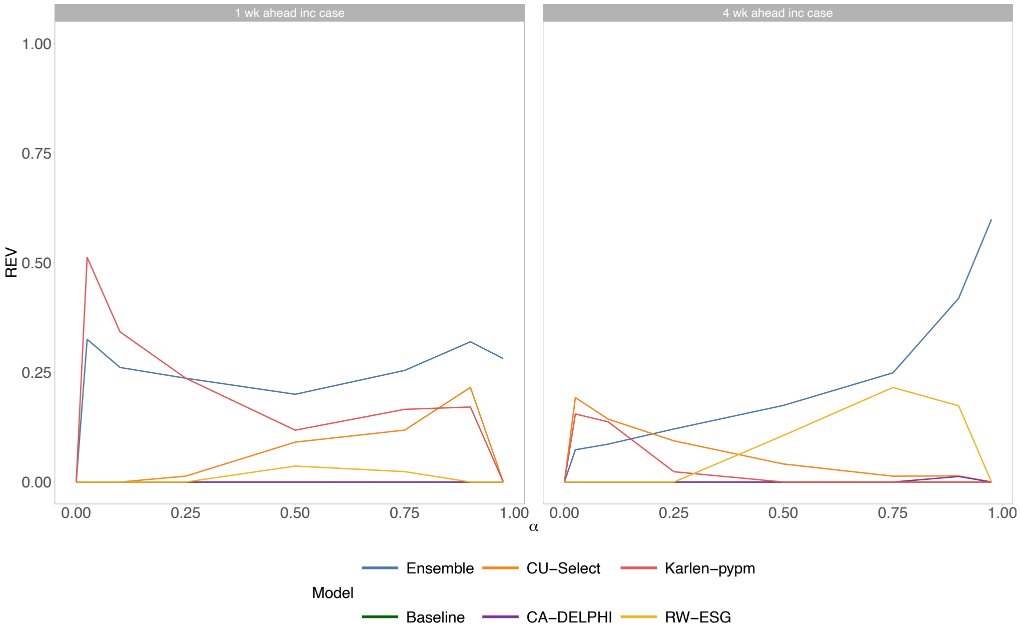


Figure SI 8: **Relative Economic Value (REV) for upper 10% of *all* observed incident cases:** REV for the different forecasting models, compared to the COVID-19 Forecast Hub baseline forecasting model which forecasts the most recent observed incident cases as its best (i.e., median) estimate of the future.

##### G.2.2 Upper 10% of each state's events

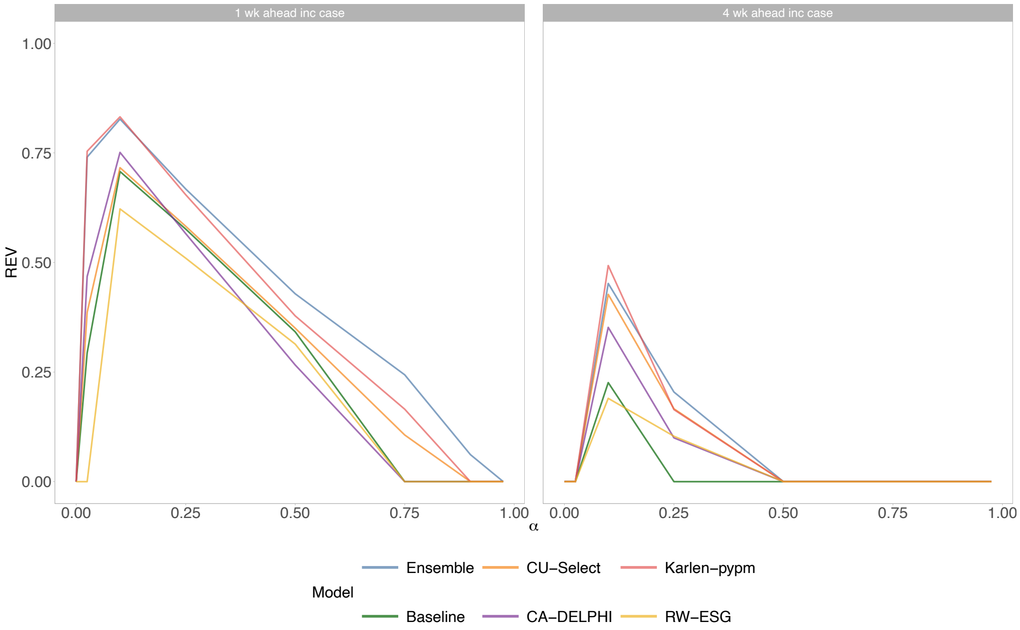


Figure SI 9: **Relative Economic Value (REV) for upper 10% of each state's observed incident cases:** REV for the different forecasting models, compared to a forecasting model which forecasts the binary event with probability of 10% (i.e., analogous to the climatology model used in weather forecasting). This differs to Figures SI 7 – SI 8 by focusing on value for the upper 10% of events (i.e., cases) observed in each state.

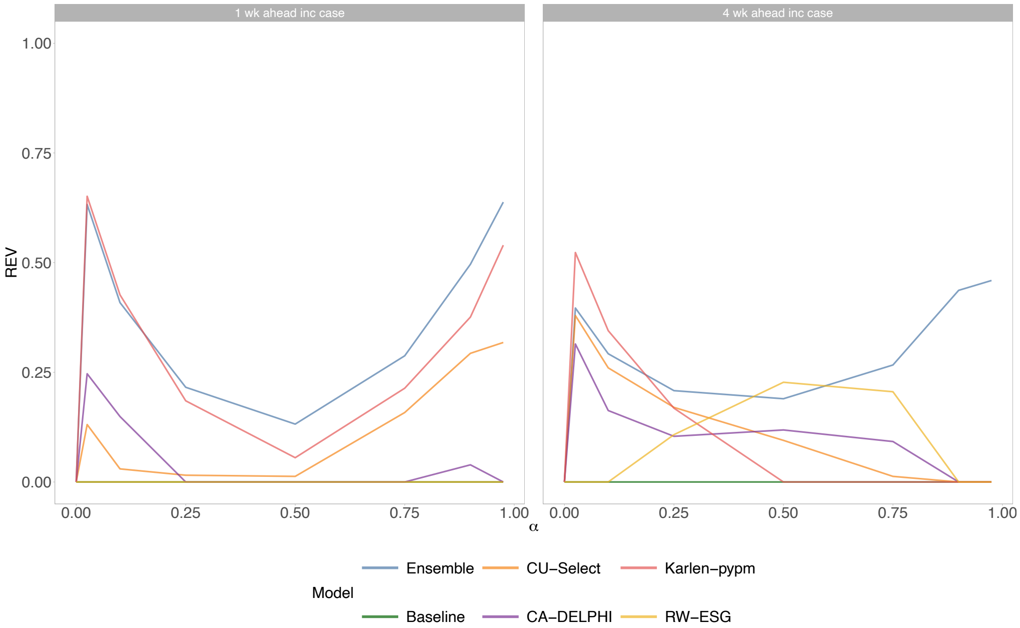


Figure SI 10: **Relative Economic Value (REV) for upper 10% of each location's observed incident cases:** REV for the different forecasting models, compared to the COVID-19 Forecast Hub baseline forecasting model which forecasts the most recent observed incident cases as its best (i.e., median) estimate of the future.

##### G.2.3 Upper 15% of all events

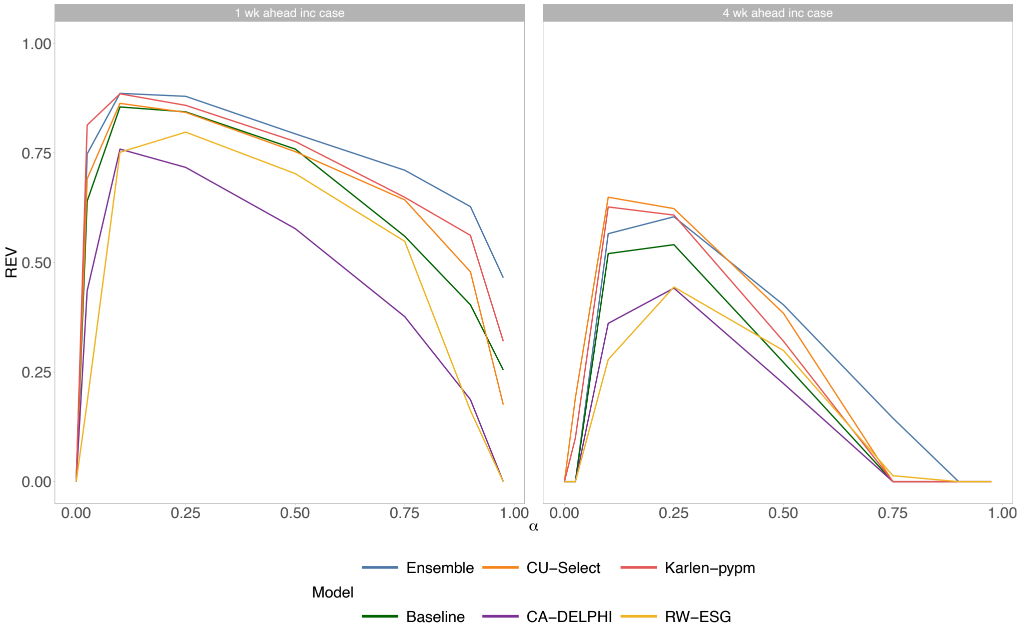


Figure SI 11: **Relative Economic Value (REV) for upper 15% of all observed incident cases:** REV for the different forecasting models, compared to a forecasting model which forecasts the binary event with probability of 15% (i.e., analogous to the climatology model used in weather forecasting). This plot has featured in Figure 3 C.3.

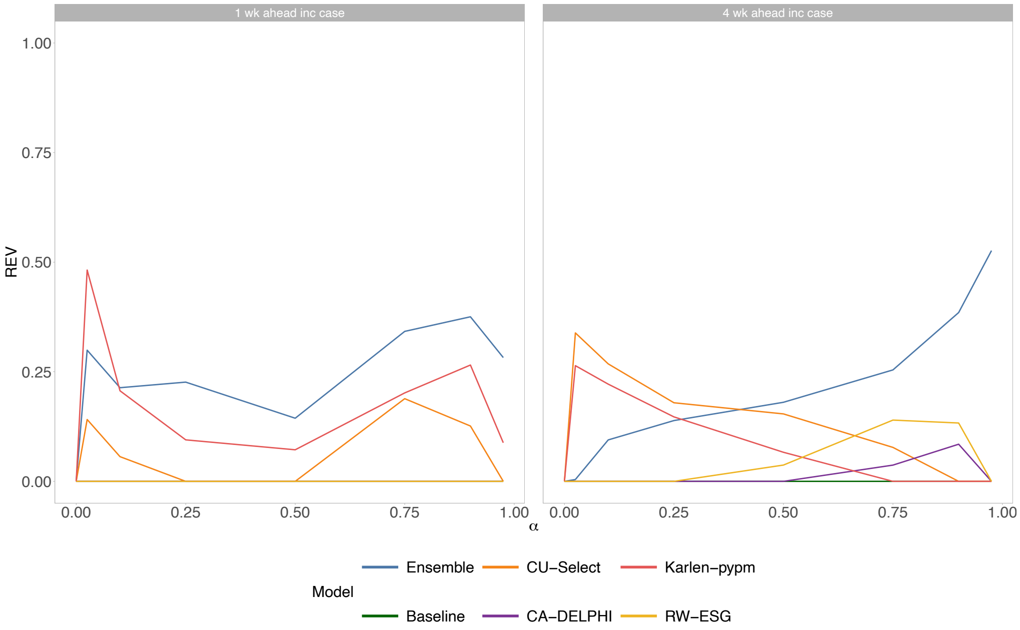


Figure SI 12: **Relative Economic Value (REV) for upper 15% of all observed incident cases:** REV for the different forecasting models, compared to the COVID-19 Forecast Hub baseline forecasting model which forecasts the most recent observed incident cases as its best (i.e., median) estimate of the future.

##### G.2.4 Upper 15% of each state's events

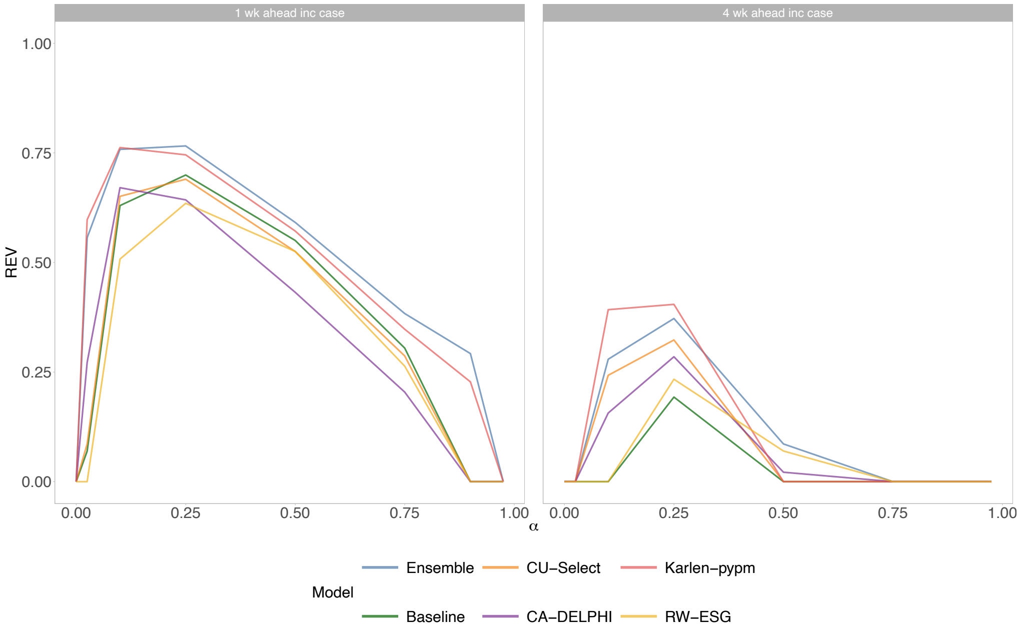


Figure SI 13: **Relative Economic Value (REV) for upper 15% of each location's observed incident cases:** REV for the different forecasting models, compared to a forecasting model which forecasts the binary event with probability of 15% (i.e., analogous to the climatology model used in weather forecasting). This differs to Figures SI 11 – SI 12 by focusing on value for the upper 15% of events (i.e., cases) observed within each state.

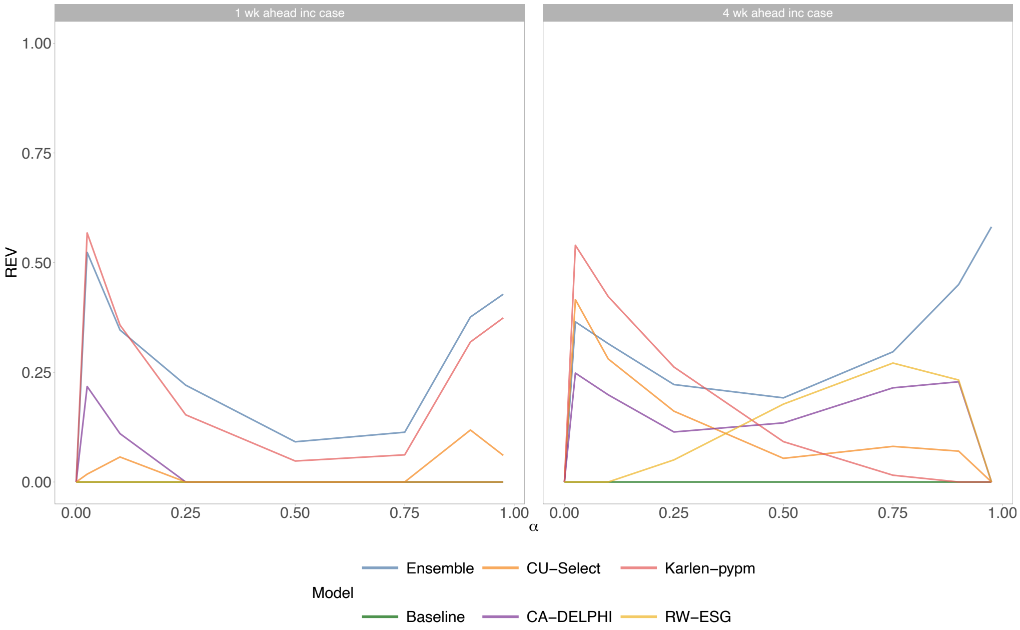


Figure SI 14: **Relative Economic Value (REV) for upper 15% of each location's observed incident cases:** REV for the different forecasting models, compared to the COVID-19 Forecast Hub baseline forecasting model which forecasts the most recent observed incident cases as its best (i.e., median) estimate of the future.

#### G.3 Relative pinball loss (rPL) across space and time

##### G.3.1 Median rPL for models across locations over time for risk-intolerant and risk-tolerant decision-makers

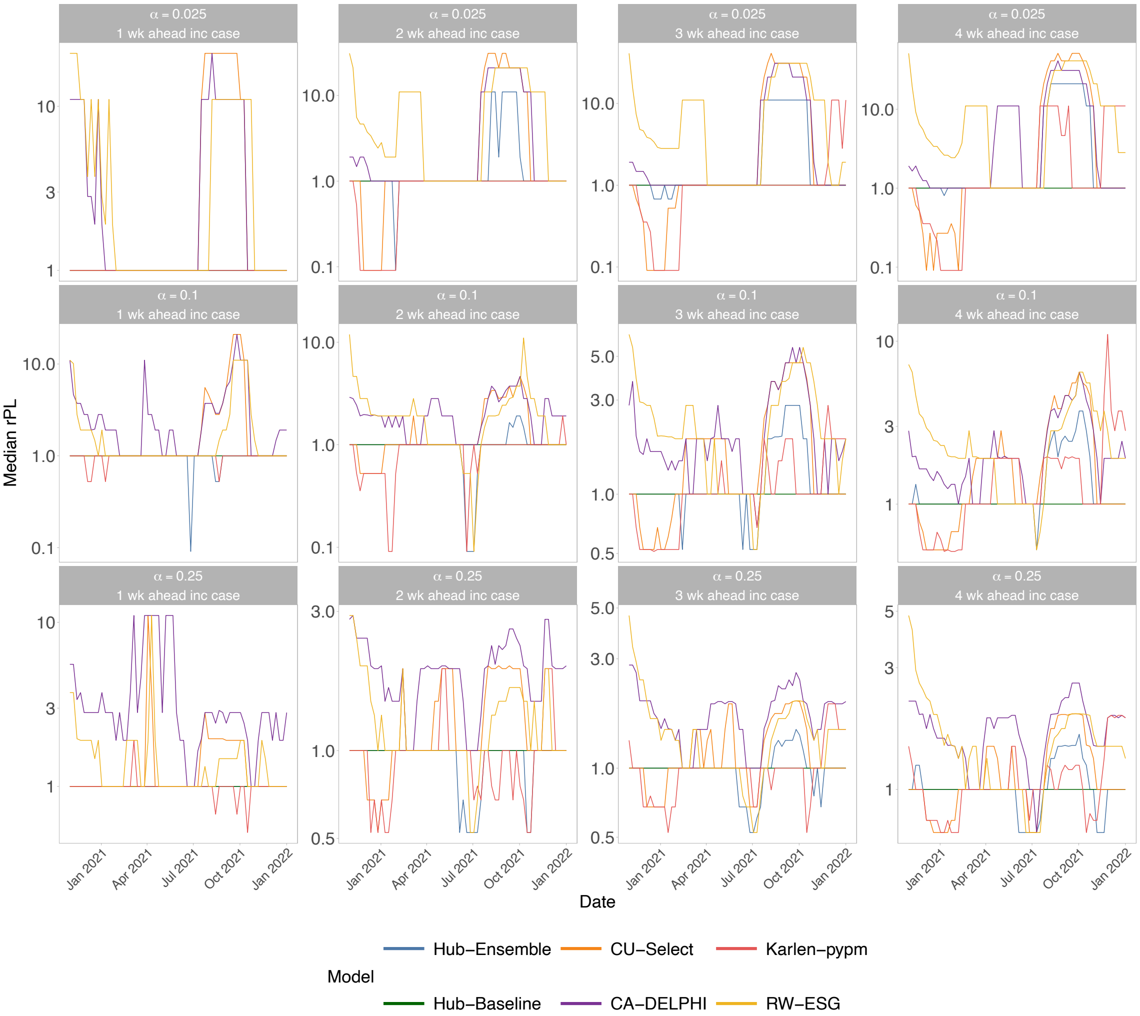


Figure SI 15: **Median relative pinball loss (rPL) for each model's forecasts across individual locations from 12-week trailing windows:** Focusing on decision-makers with low C/L ratios (i.e., cost of action is inexpensive relative to harm), median rPL across all states (y-axis) is visualised for each model and each trailing window where the x-axis denotes the maximum date of the 12-week trailing window. We calculated rPL relative to the baseline model, and rPL $< 1$ indicates greater forecast value for that decision-maker in the trailing window.

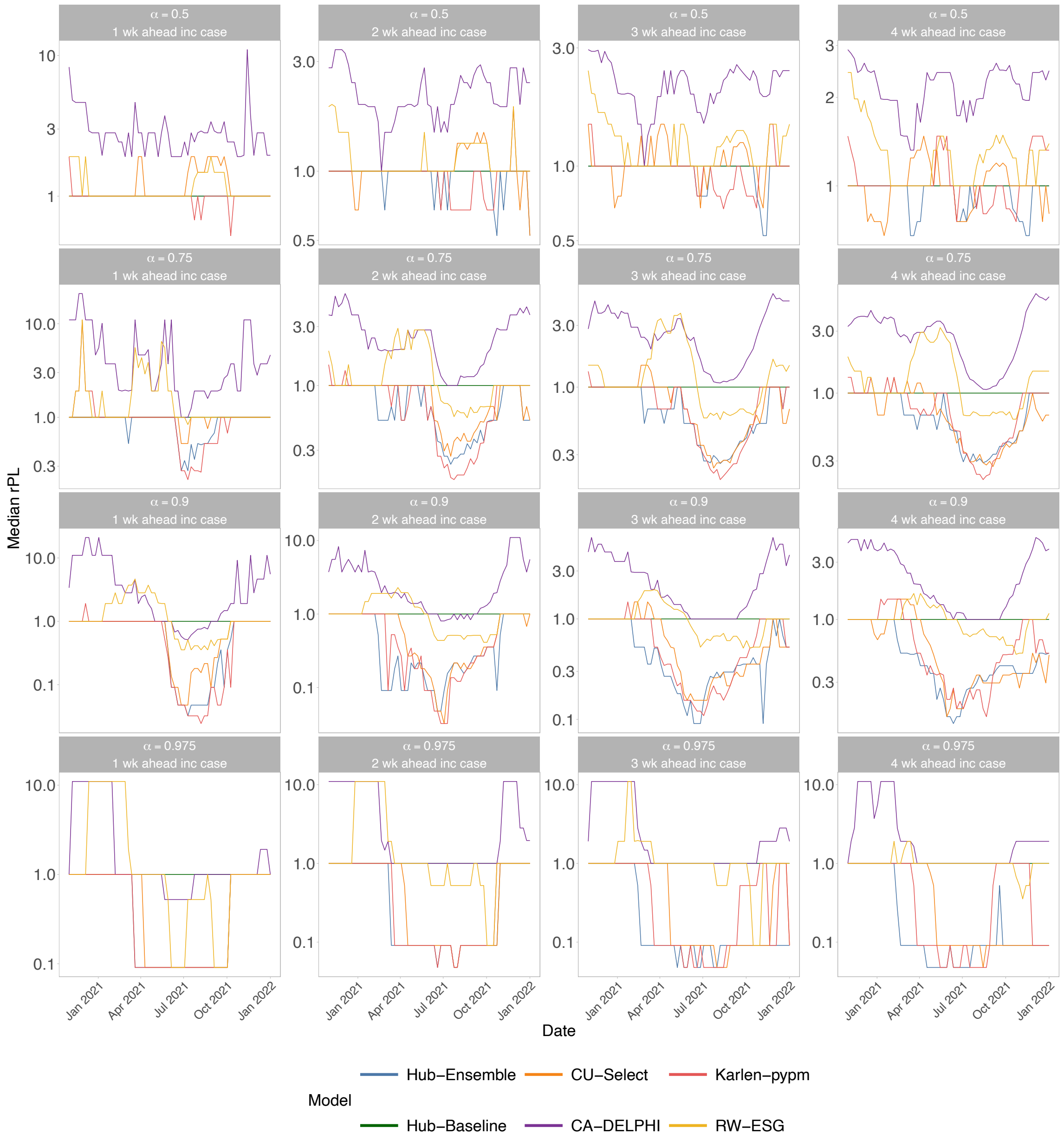


Figure SI 16: **Median relative pinball loss (rPL) for each model's forecasts across individual locations from 12-week trailing windows:** Focusing on decision-makers with high C/L ratios (i.e., cost of action is expensive relative to harm), median rPL across all states (y-axis) is visualised for each model and each trailing window where the x-axis denotes the maximum date of the 12-week trailing window. We calculated rPL relative to the baseline model, and rPL $< 1$ indicates greater forecast value for that decision-maker in the trailing window.

##### G.3.2 rPL for the ensemble model's one-week-ahead forecasts at low and high C/L ratios

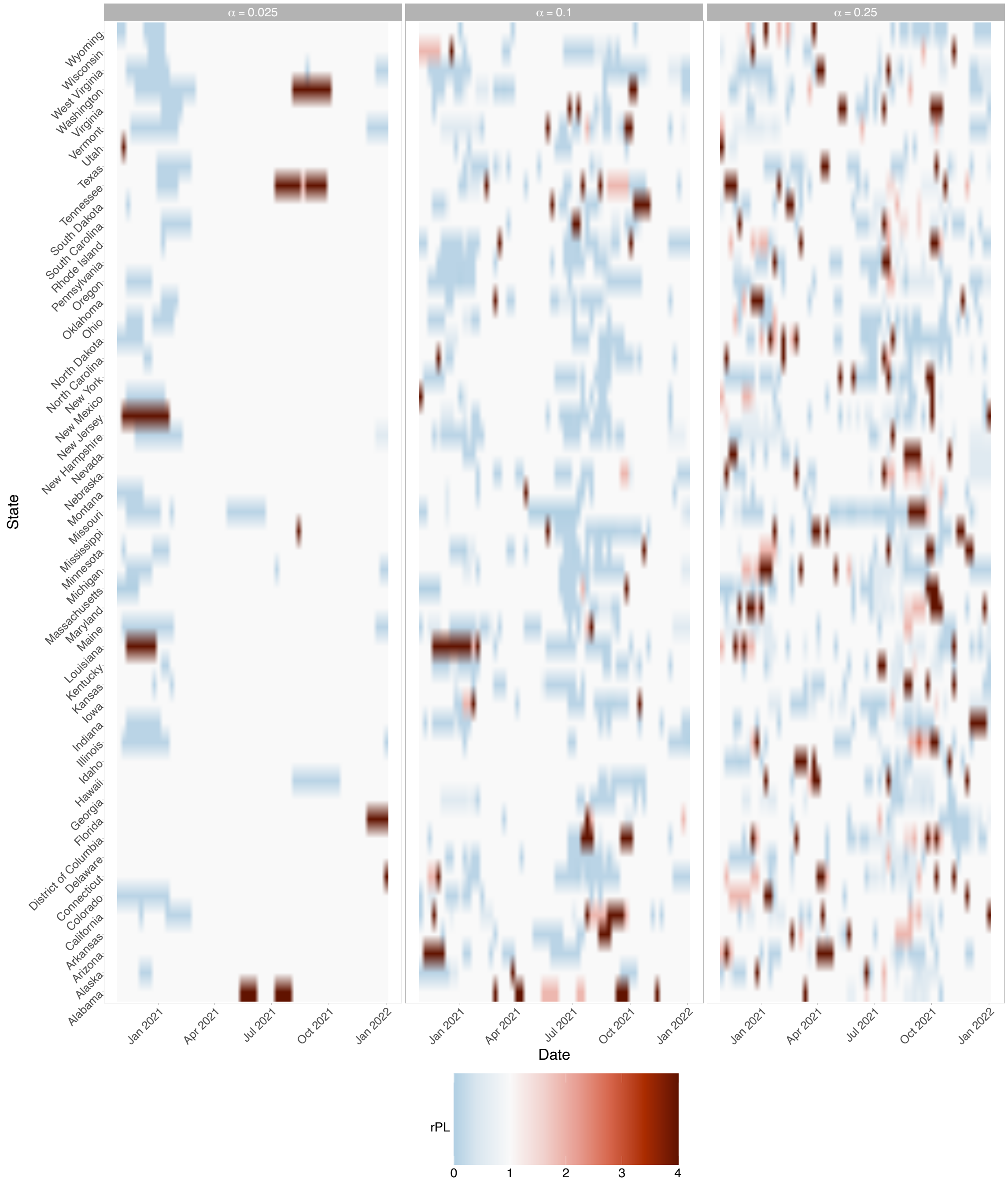


Figure SI 17: **The relative pinball loss (rPL) for the ensemble's one-week-ahead forecasts for individual locations from 12-week trailing windows:** Focusing on risk-averse decision-makers with low C/L ratios (i.e., cost of action is inexpensive relative to harm), rPL is visualised for each state (y-axis) and each trailing window where the x-axis denotes the maximum date of the 12-week trailing window. We calculated rPL relative to the baseline model, and rPL < 1 indicates greater forecast value for that decision-maker in the given location and trailing window.

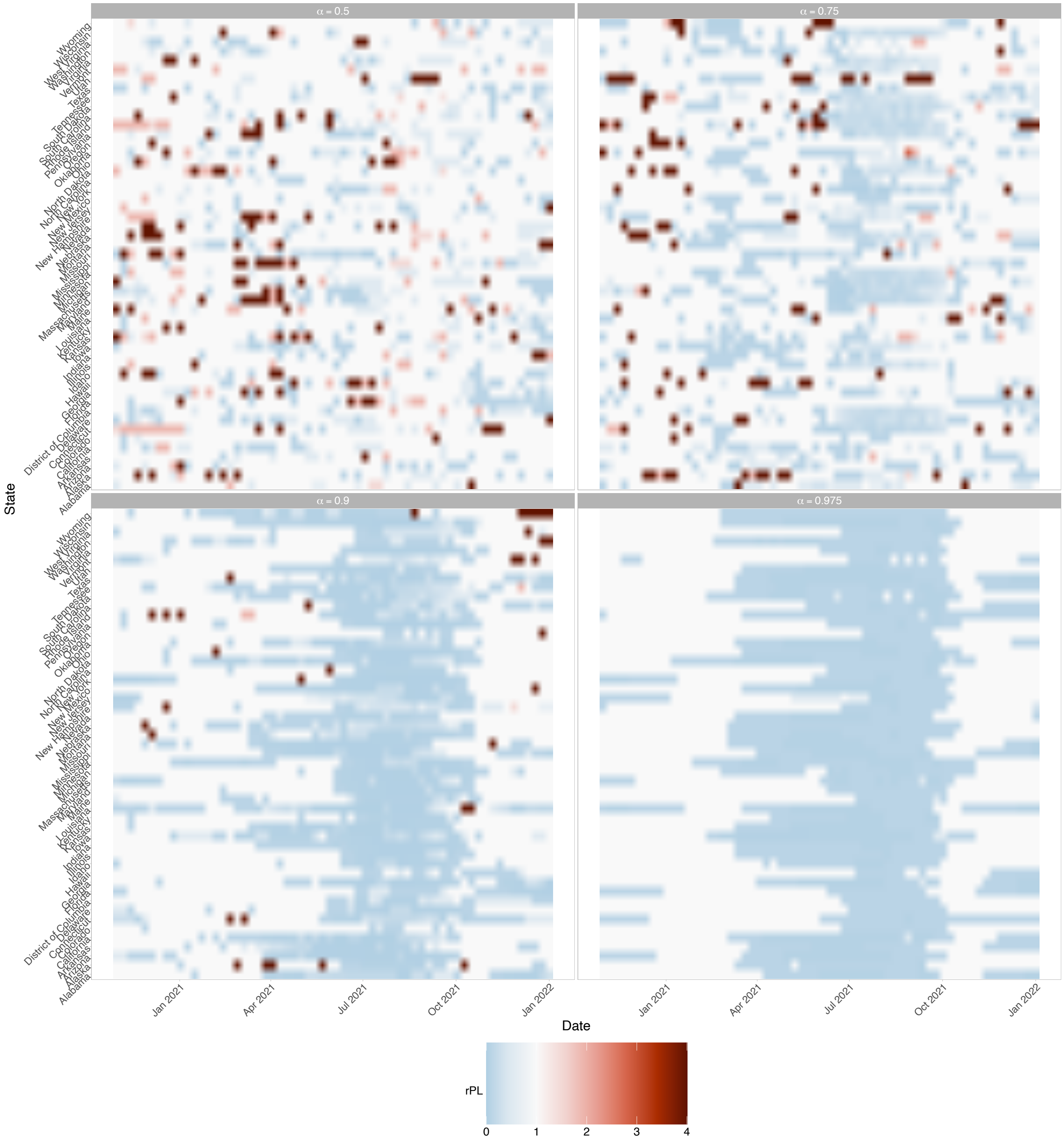


Figure SI 18: **The relative pinball loss (rPL) for the ensemble's one-week-ahead forecasts for individual locations from 12-week trailing windows:** Focusing on risk-tolerant decision-makers with hi/gh C/L ratios (i.e., cost of action is high relative to harm), rPL is visualised for each state (y-axis) and each trailing window where the x-axis denotes the maximum date of the 12-week trailing window. We calculated rPL relative to the baseline model, and rPL $< 1$ indicates greater forecast value for that decision-maker in the given location and trailing window.

##### G.3.3 rPL for the ensemble model's two-weeks-ahead forecasts at low and high C/L ratios

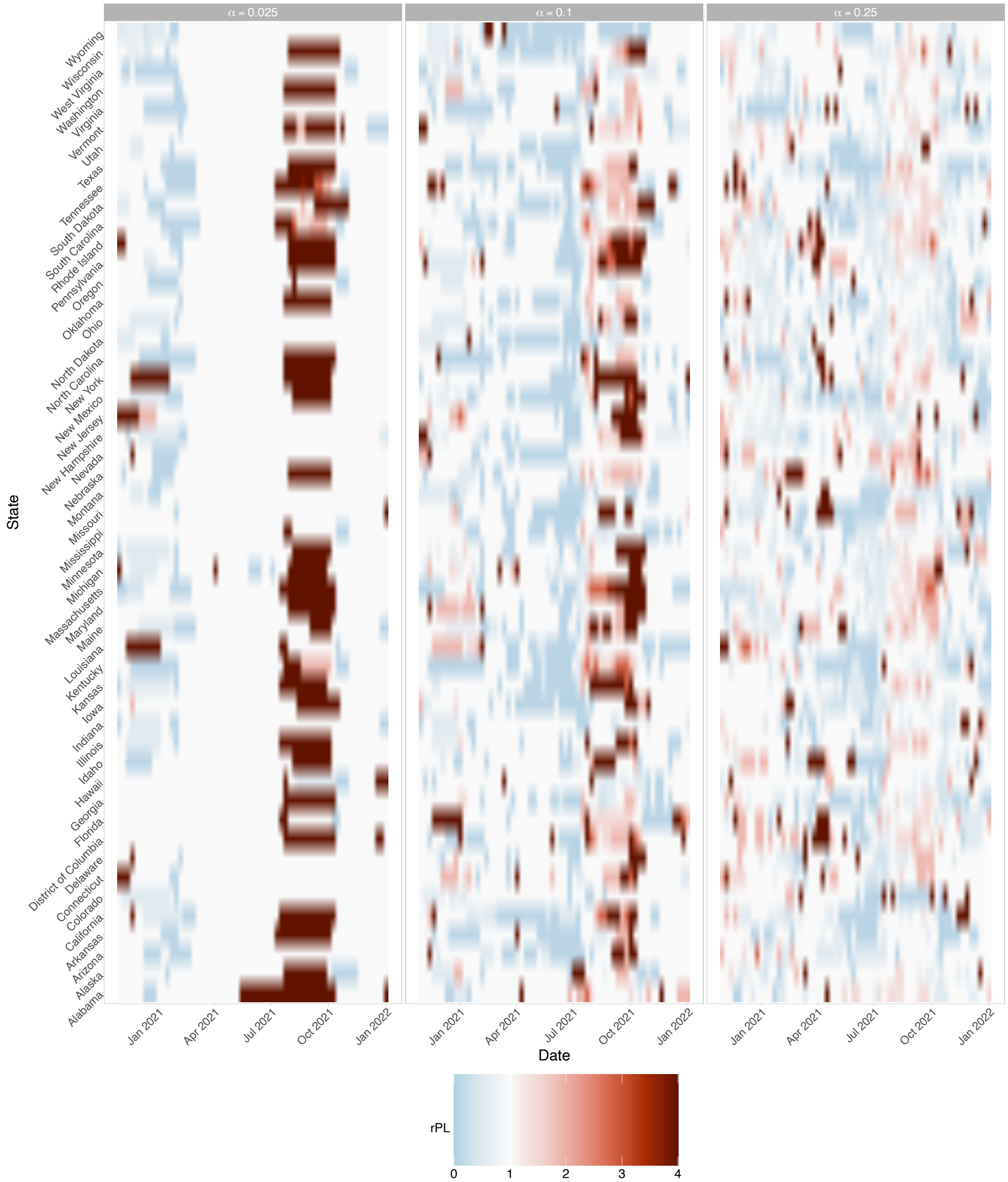


Figure SI 19: **The relative pinball loss (rPL) for the ensemble's two-weeks-ahead forecasts for individual locations from 12-week trailing windows:** Focusing on risk-averse decision-makers with low C/L ratios (i.e., cost of action is inexpensive relative to harm), rPL is visualised for each state (y-axis) and each trailing window where the x-axis denotes the maximum date of the 12-week trailing window. We calculated rPL relative to the baseline model, and rPL < 1 indicates greater forecast value for that decision-maker in the given location and trailing window.

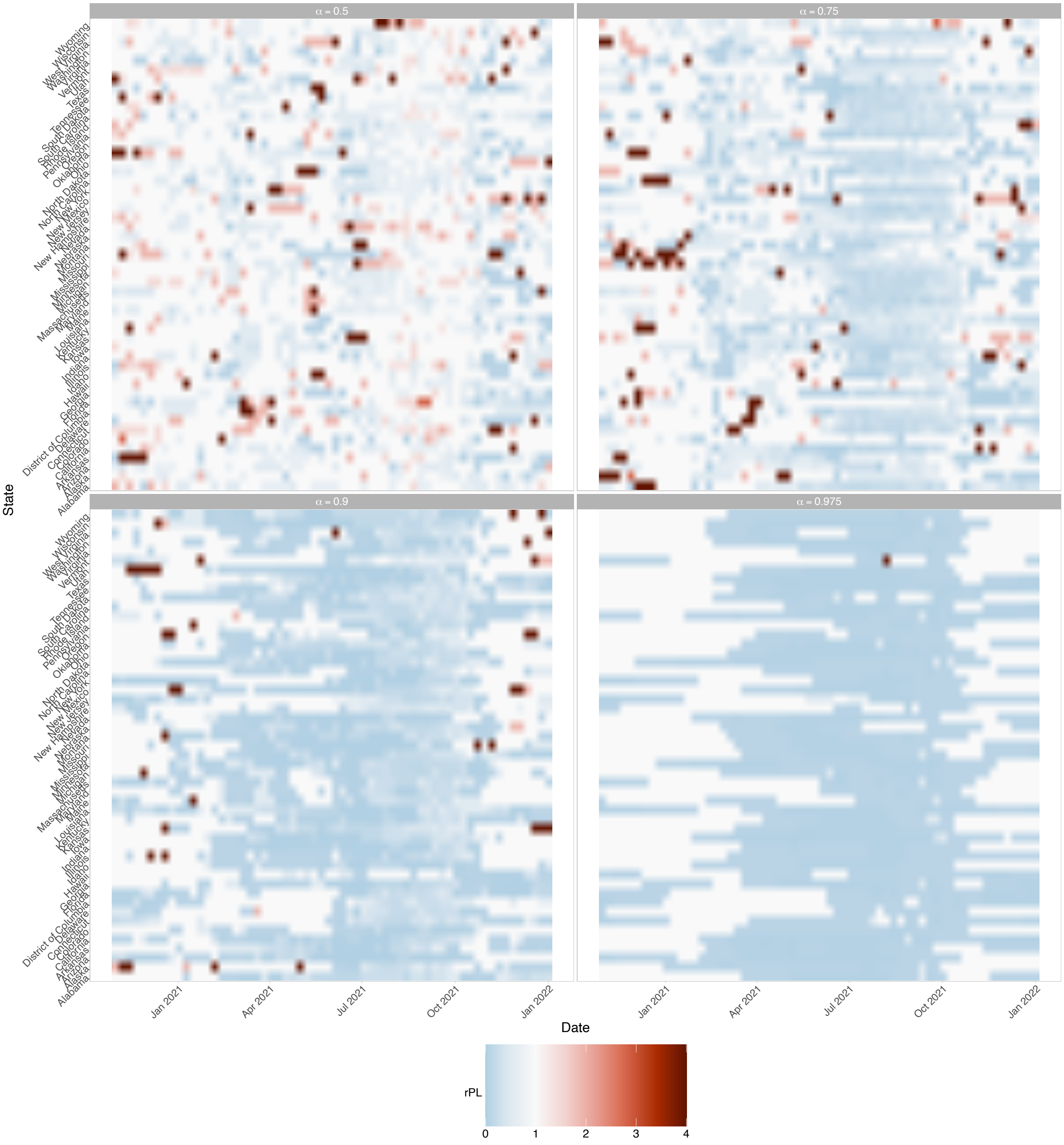


Figure SI 20: **The relative pinball loss (rPL) for the ensemble's two-weeks-ahead forecasts for individual locations from 12-week trailing windows:** Focusing on risk-tolerant decision-makers with high C/L ratios (i.e., cost of action is high relative to harm), rPL is visualised for each state (y-axis) and each trailing window where the x-axis denotes the maximum date of the 12-week trailing window. We calculated rPL relative to the baseline model, and rPL $< 1$ indicates greater forecast value for that decision-maker in the given location and trailing window.

##### G.3.4 rPL for the ensemble model's three-weeks-ahead forecasts at low and high C/L ratios

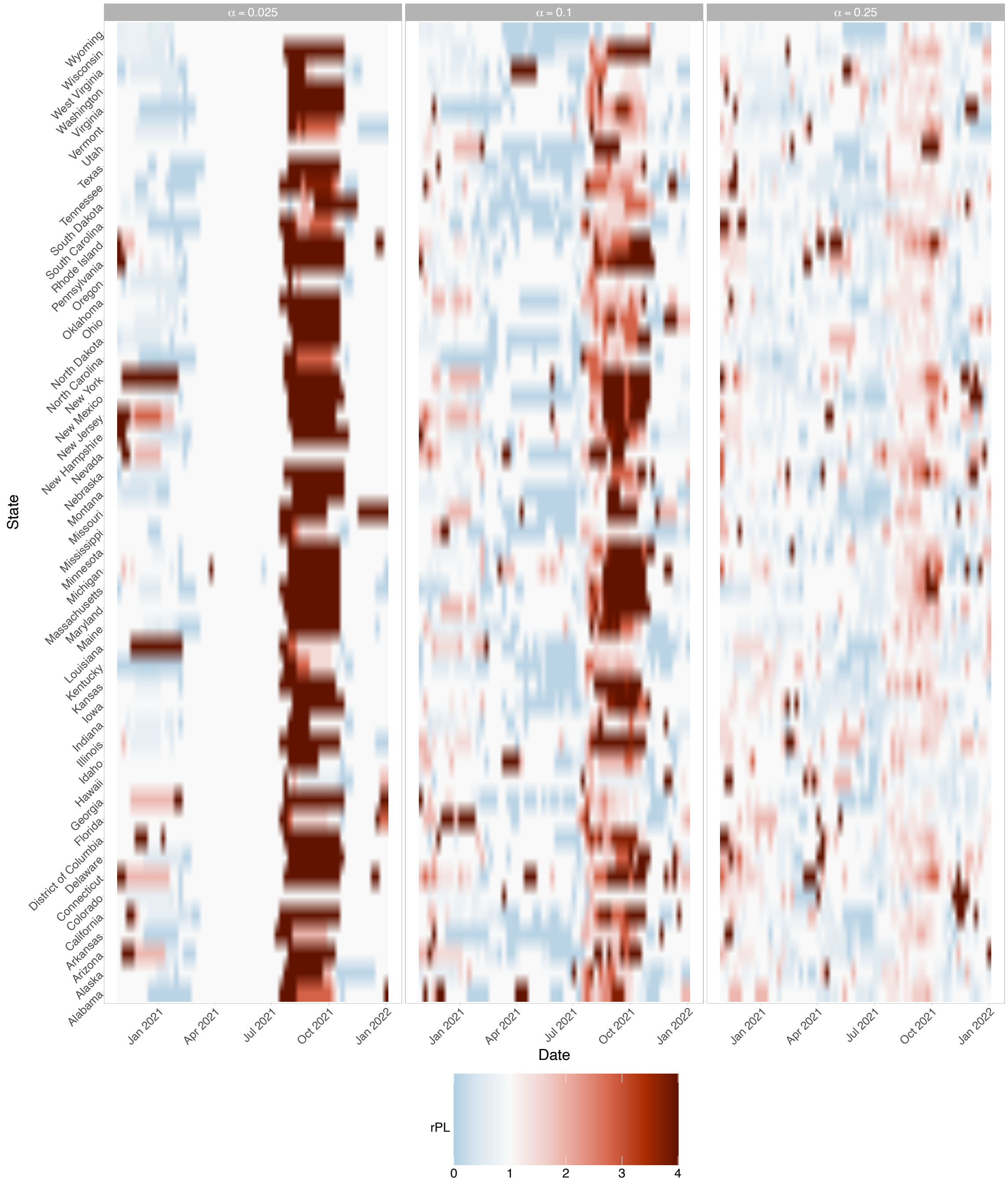


Figure SI 21: **The relative pinball loss (rPL) for the ensemble's two-weeks-ahead forecasts for individual locations from 12-week trailing windows:** Focusing on risk-averse decision-makers with low C/L ratios (i.e., cost of action is inexpensive relative to harm), rPL is visualised for each state (y-axis) and each trailing window where the x-axis denotes the maximum date of the 12-week trailing window. We calculated rPL relative to the baseline model, and rPL $< 1$ indicates greater forecast value for that decision-maker in the given location and trailing window.

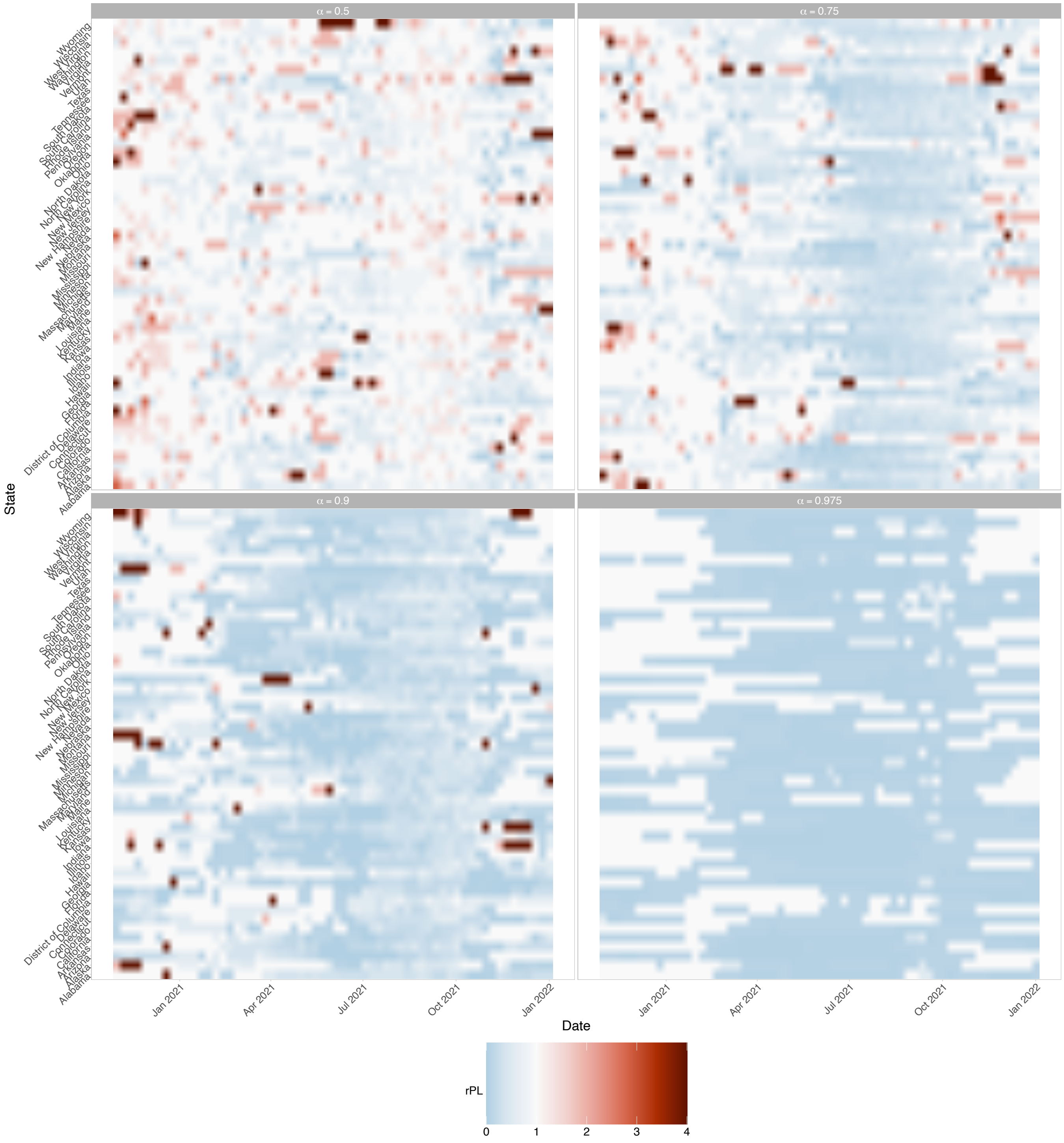


Figure SI 22: **The relative pinball loss (rPL) for the ensemble's three-weeks-ahead forecasts for individual locations from 12-week trailing windows:** Focusing on risk-tolerant decision-makers with high C/L ratios (i.e., cost of action is high relative to harm), rPL is visualised for each state (y-axis) and each trailing window where the x-axis denotes the maximum date of the 12-week trailing window. We calculated rPL relative to the baseline model, and rPL < 1 indicates greater forecast value for that decision-maker in the given location and trailing window.

##### G.3.5 rPL for the ensemble model's four-weeks-ahead forecasts at low and high C/L ratios

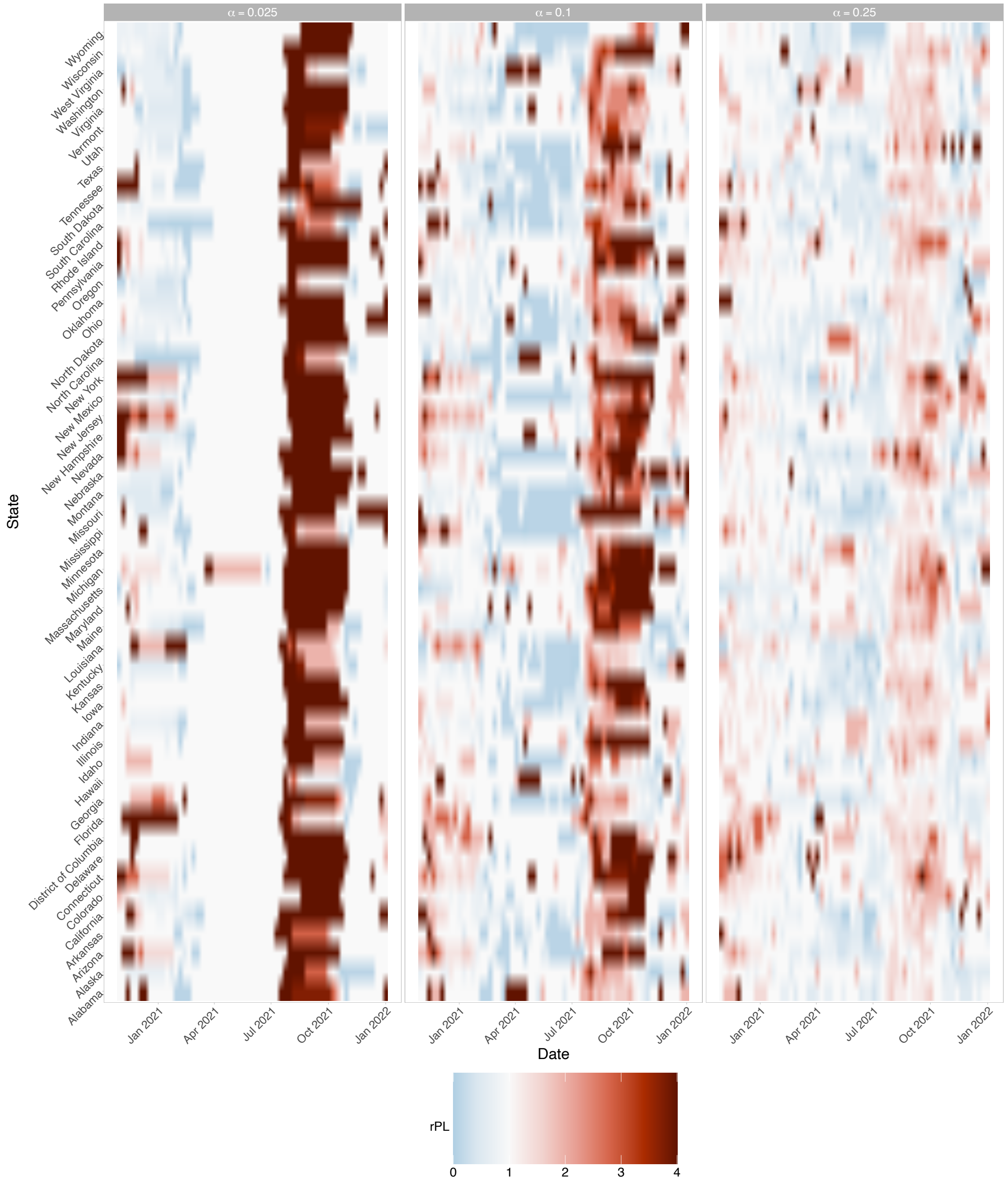


Figure SI 23: **The relative pinball loss (rPL) for the ensemble's four-weeks-ahead forecasts for individual locations from 12-week trailing windows:** Focusing on risk-averse decision-makers with low C/L ratios (i.e., cost of action is inexpensive relative to harm), rPL is visualised for each state (y-axis) and each trailing window where the x-axis denotes the maximum date of the 12-week trailing window. We calculated rPL relative to the baseline model, and rPL $< 1$ indicates greater forecast value for that decision-maker in the given location and trailing window.

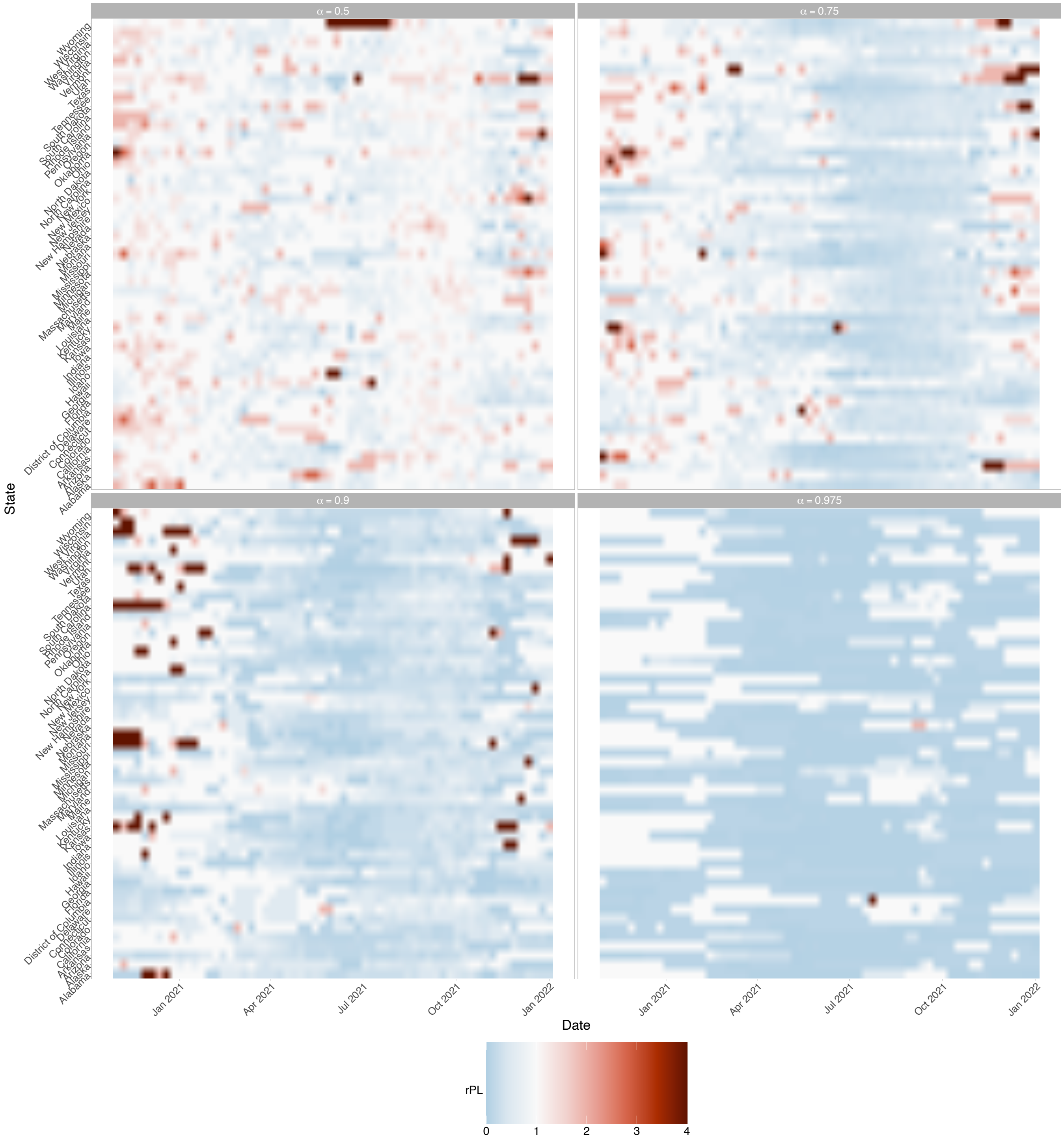


Figure SI 24: **The relative pinball loss (rPL) for the ensemble's four-weeks-ahead forecasts for individual locations from 12-week trailing windows:** Focusing on decision-makers with high C/L ratios (i.e., cost of action is high relative to harm), rPL is visualised for each state (y-axis) and each trailing window where the x-axis denotes the maximum date of the 12-week trailing window. We calculated rPL relative to the baseline model, and rPL $< 1$ indicates greater forecast value for that decision-maker in the given location and trailing window.

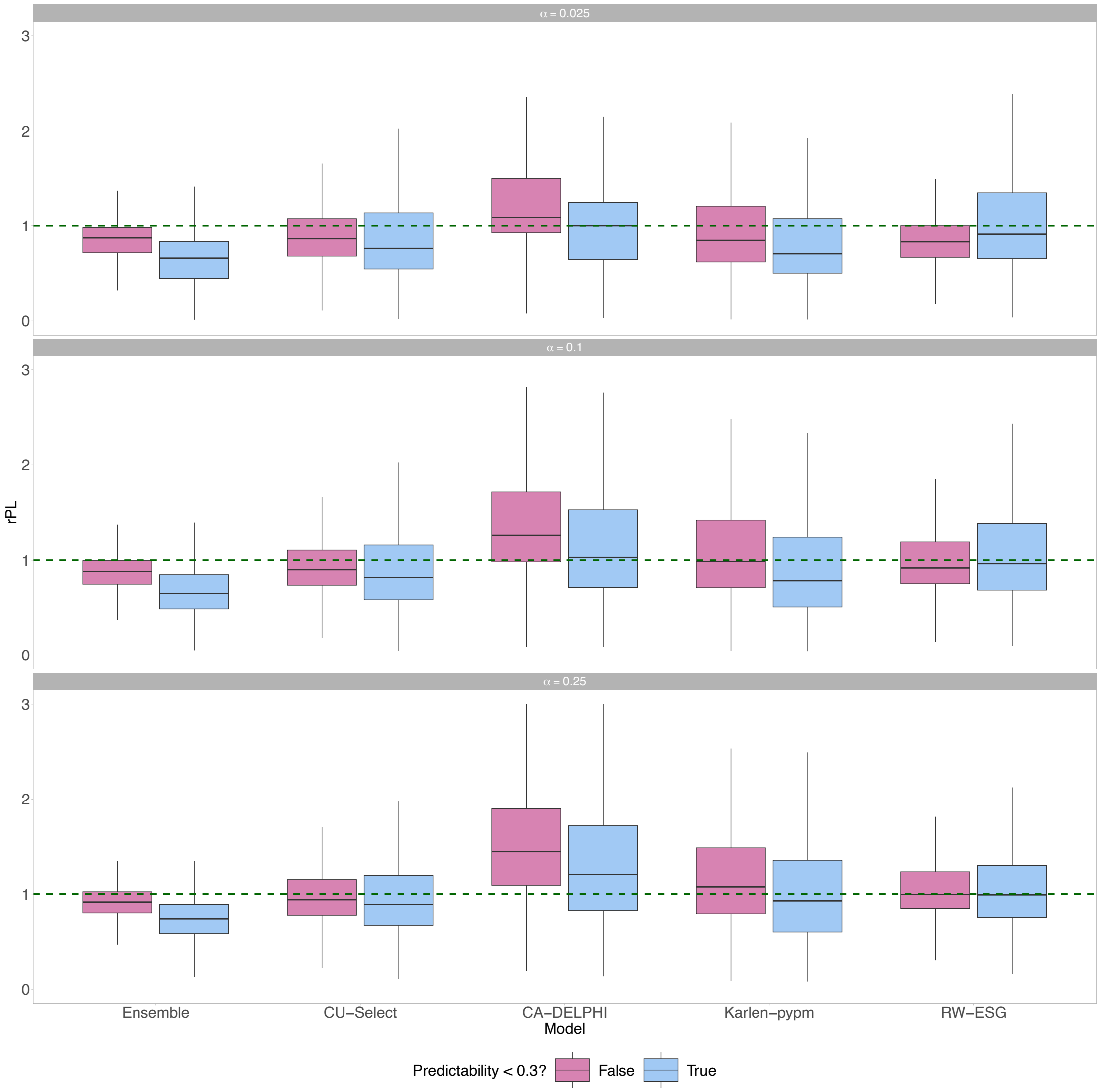


Figure SI 25: **Relative pinball loss summary by low/high predictability regimes from trailing windows:** For visualisation, we use a binary classification here of predictability $< 0.3$ and focus on risk-intolerant decision-makers. Values of relative pinball loss (rPL) below 1 indicate greater forecast value at that fixed C/L ratio across all observations compared to the baseline model.

#### G.4 User-specific Murphy diagrams for higher C/L ratios

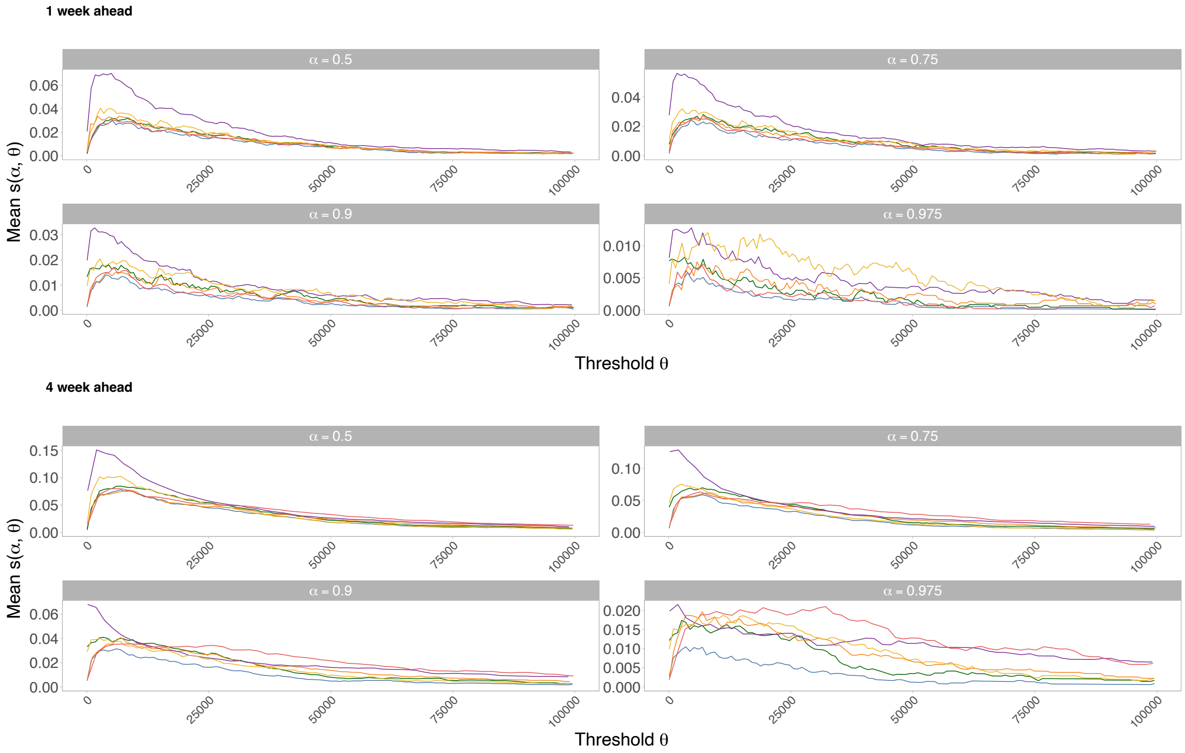


Figure SI 26: **User-specific Murphy diagrams for higher C/L ratios:** We visualise Murphy diagrams for risk-tolerant decision-makers for whom the defined action is expensive relative to the loss/harm (i.e., high C/L ratios). Each Murphy curve captures mean elementary score $s(\alpha, \theta)$ for a fixed C/L ratio ($\alpha$) and varying event thresholds ($\theta$). Lower values on Murphy curves indicate superior performance of a model's forecasts for the given user-event combination. Figure 3 C.1 is the analogous visualisation for risk-intolerant decision-makers for whom the defined action is inexpensive relative to the loss/harm (i.e., low C/L ratios).

#### G.5 Predictability versus forecast metrics

##### G.5.1 Predictability versus $R^2$

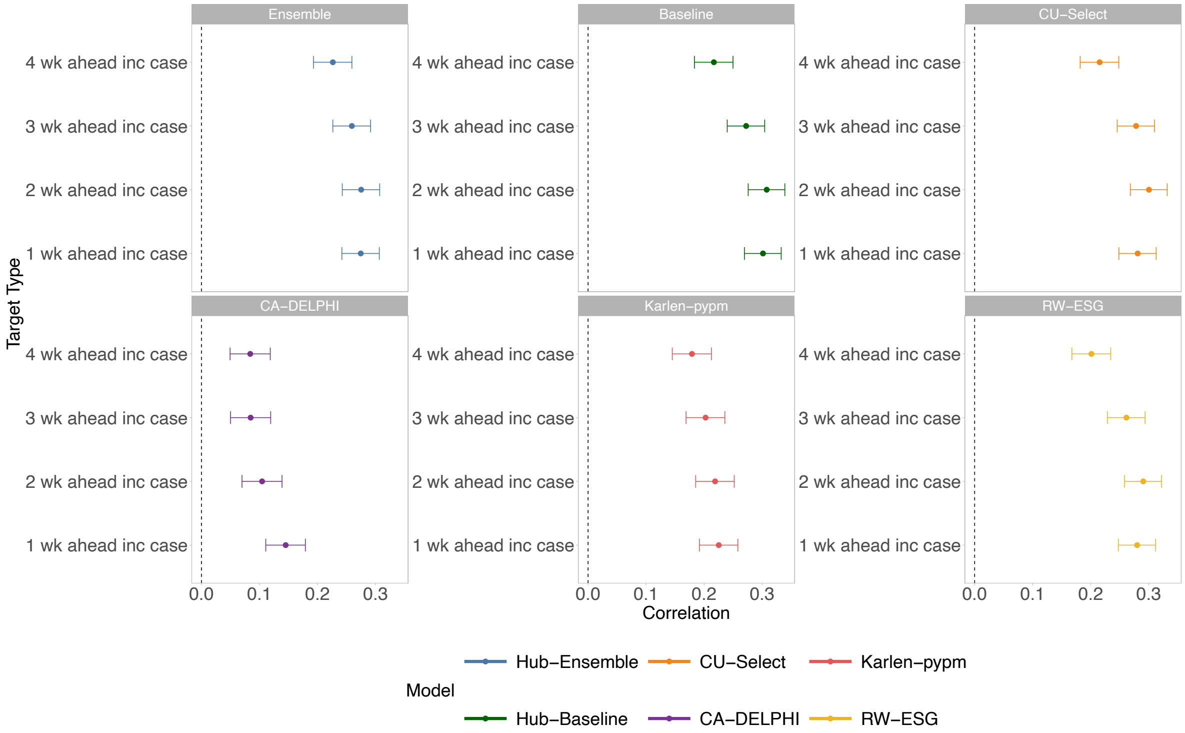


Figure SI 27: **Correlations between predictability and $R^2$:** We visualise the Pearson's correlations (Table SI 4) between predictability and $R^2$ for each model and forecast target type, alongside the 95% Confidence Intervals (CIs) calculated using Fisher z-transformations.

Table SI 4: **Correlations between predictability and $R^2$:** For each model and forecasted quantity, we report the Pearson's correlation $r$ between predictability and $R^2$, alongside the 95% Confidence Intervals (CIs) calculated using Fisher z-transformations and the Bonferroni-adjusted p-value for the two-sided hypothesis test with null hypothesis that $r = 0$.

| Model | Target Type | r | 95% CI | Bonferroni-adjusted $p$ |
|---|---|---|---|---|
| Ensemble | 1 wk ahead inc case | 0.27 | [0.24, 0.31] | P < 0.001 |
| Ensemble | 2 wk ahead inc case | 0.28 | [0.24, 0.31] | P < 0.001 |
| Ensemble | 3 wk ahead inc case | 0.26 | [0.23, 0.29] | P < 0.001 |
| Ensemble | 4 wk ahead inc case | 0.23 | [0.19, 0.26] | P < 0.001 |
| COVIDhub-baseline | 1 wk ahead inc case | 0.30 | [0.27, 0.33] | P < 0.001 |
| COVIDhub-baseline | 2 wk ahead inc case | 0.31 | [0.28, 0.34] | P < 0.001 |
| COVIDhub-baseline | 3 wk ahead inc case | 0.27 | [0.24, 0.30] | P < 0.001 |
| COVIDhub-baseline | 4 wk ahead inc case | 0.22 | [0.18, 0.25] | P < 0.001 |
| CU-select | 1 wk ahead inc case | 0.28 | [0.25, 0.31] | P < 0.001 |
| CU-select | 2 wk ahead inc case | 0.30 | [0.27, 0.33] | P < 0.001 |
| CU-select | 3 wk ahead inc case | 0.28 | [0.25, 0.31] | P < 0.001 |
| CU-select | 4 wk ahead inc case | 0.22 | [0.18, 0.25] | P < 0.001 |
| CovidAnalytics-DELPHI | 1 wk ahead inc case | 0.15 | [0.11, 0.18] | P < 0.001 |
| CovidAnalytics-DELPHI | 2 wk ahead inc case | 0.10 | [0.07, 0.14] | P < 0.001 |
| CovidAnalytics-DELPHI | 3 wk ahead inc case | 0.08 | [0.05, 0.12] | P < 0.001 |
| CovidAnalytics-DELPHI | 4 wk ahead inc case | 0.08 | [0.05, 0.12] | P < 0.001 |
| Karlen-pypm | 1 wk ahead inc case | 0.23 | [0.19, 0.26] | P < 0.001 |
| Karlen-pypm | 2 wk ahead inc case | 0.22 | [0.19, 0.25] | P < 0.001 |
| Karlen-pypm | 3 wk ahead inc case | 0.20 | [0.17, 0.24] | P < 0.001 |
| Karlen-pypm | 4 wk ahead inc case | 0.18 | [0.15, 0.21] | P < 0.001 |
| RobertWalraven-ESG | 1 wk ahead inc case | 0.28 | [0.25, 0.31] | P < 0.001 |
| RobertWalraven-ESG | 2 wk ahead inc case | 0.29 | [0.26, 0.32] | P < 0.001 |
| RobertWalraven-ESG | 3 wk ahead inc case | 0.26 | [0.23, 0.29] | P < 0.001 |
| RobertWalraven-ESG | 4 wk ahead inc case | 0.20 | [0.17, 0.23] | P < 0.001 |

##### G.5.2 Predictability versus 95% Prediction Interval (PI) Coverage

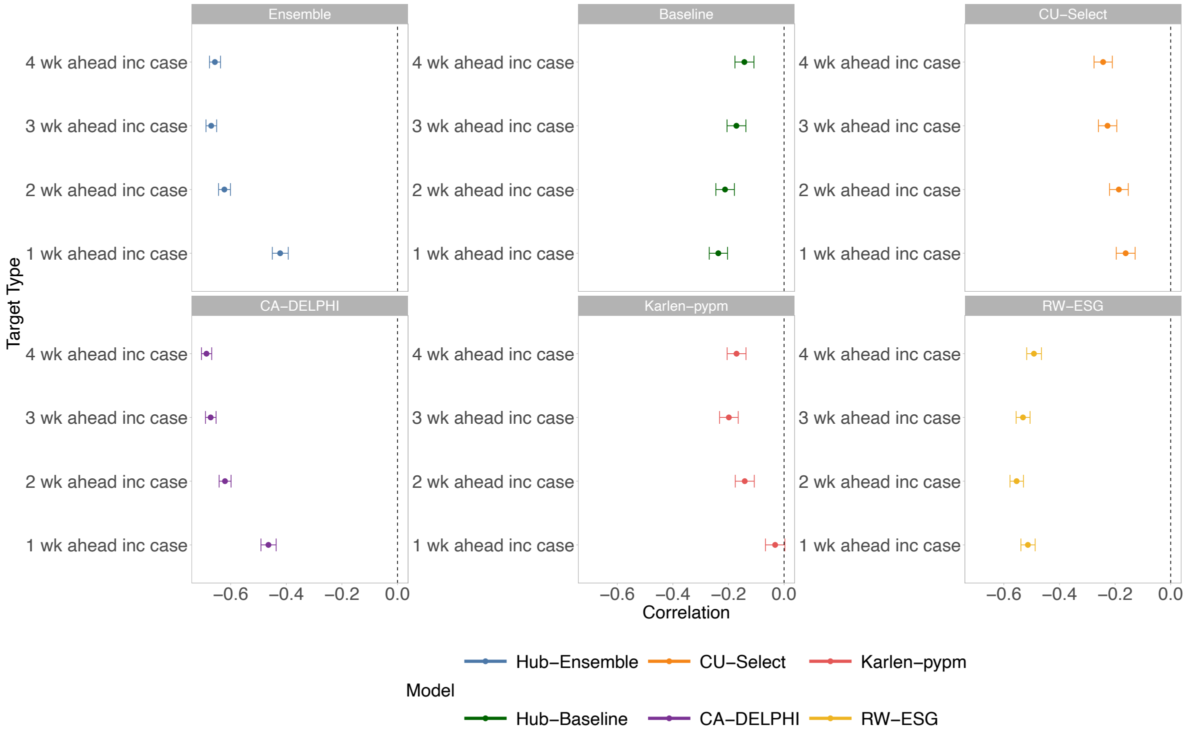


Figure SI 28: **Correlations between predictability and central 95% prediction interval (PI) coverage:** We visualise the Pearson's correlations (Table SI 5) between predictability and 95% PI coverage for each model and forecast target type, alongside the 95% Confidence Intervals (CIs) calculated using Fisher z-transformations.

Table SI 5: **Correlations between predictability and central 95% Prediction Interval (PI) coverage:** For each model and forecasted quantity, we report the Pearson's correlation $r$ between predictability and 95% PI, alongside the 95% Confidence Intervals (CIs) calculated using Fisher z-transformations and the Bonferroni-adjusted p-value for the two-sided hypothesis test with null hypothesis that $r = 0$.

| Model | Target Type | r | 95% CI | Bonferroni-adjusted $p$ |
|---|---|---|---|---|
| Ensemble | 1 wk ahead inc case | -0.42 | [-0.45, -0.39] | P < 0.001 |
| Ensemble | 2 wk ahead inc case | -0.62 | [-0.64, -0.60] | P < 0.001 |
| Ensemble | 3 wk ahead inc case | -0.67 | [-0.69, -0.65] | P < 0.001 |
| Ensemble | 4 wk ahead inc case | -0.66 | [-0.68, -0.64] | P < 0.001 |
| COVIDhub-baseline | 1 wk ahead inc case | -0.24 | [-0.27, -0.20] | P < 0.001 |
| COVIDhub-baseline | 2 wk ahead inc case | -0.21 | [-0.25, -0.18] | P < 0.001 |
| COVIDhub-baseline | 3 wk ahead inc case | -0.17 | [-0.21, -0.14] | P < 0.001 |
| COVIDhub-baseline | 4 wk ahead inc case | -0.14 | [-0.18, -0.11] | P < 0.001 |
| CU-select | 1 wk ahead inc case | -0.16 | [-0.20, -0.13] | P < 0.001 |
| CU-select | 2 wk ahead inc case | -0.19 | [-0.22, -0.15] | P < 0.001 |
| CU-select | 3 wk ahead inc case | -0.23 | [-0.26, -0.19] | P < 0.001 |
| CU-select | 4 wk ahead inc case | -0.24 | [-0.28, -0.21] | P < 0.001 |
| CovidAnalytics-DELPHI | 1 wk ahead inc case | -0.46 | [-0.49, -0.44] | P < 0.001 |
| CovidAnalytics-DELPHI | 2 wk ahead inc case | -0.62 | [-0.64, -0.60] | P < 0.001 |
| CovidAnalytics-DELPHI | 3 wk ahead inc case | -0.67 | [-0.69, -0.65] | P < 0.001 |
| CovidAnalytics-DELPHI | 4 wk ahead inc case | -0.69 | [-0.71, -0.67] | P < 0.001 |
| Karlen-pypm | 1 wk ahead inc case | -0.03 | [-0.07, 0.00] | P = 1 |
| Karlen-pypm | 2 wk ahead inc case | -0.14 | [-0.18, -0.11] | P < 0.001 |
| Karlen-pypm | 3 wk ahead inc case | -0.20 | [-0.23, -0.16] | P < 0.001 |
| Karlen-pypm | 4 wk ahead inc case | -0.17 | [-0.20, -0.14] | P < 0.001 |
| RobertWalraven-ESG | 1 wk ahead inc case | -0.51 | [-0.54, -0.49] | P < 0.001 |
| RobertWalraven-ESG | 2 wk ahead inc case | -0.55 | [-0.58, -0.53] | P < 0.001 |
| RobertWalraven-ESG | 3 wk ahead inc case | -0.53 | [-0.56, -0.51] | P < 0.001 |
| RobertWalraven-ESG | 4 wk ahead inc case | -0.49 | [-0.52, -0.46] | P < 0.001 |

##### G.5.3 Predictability versus relative Weighted Interval Score (rWIS)

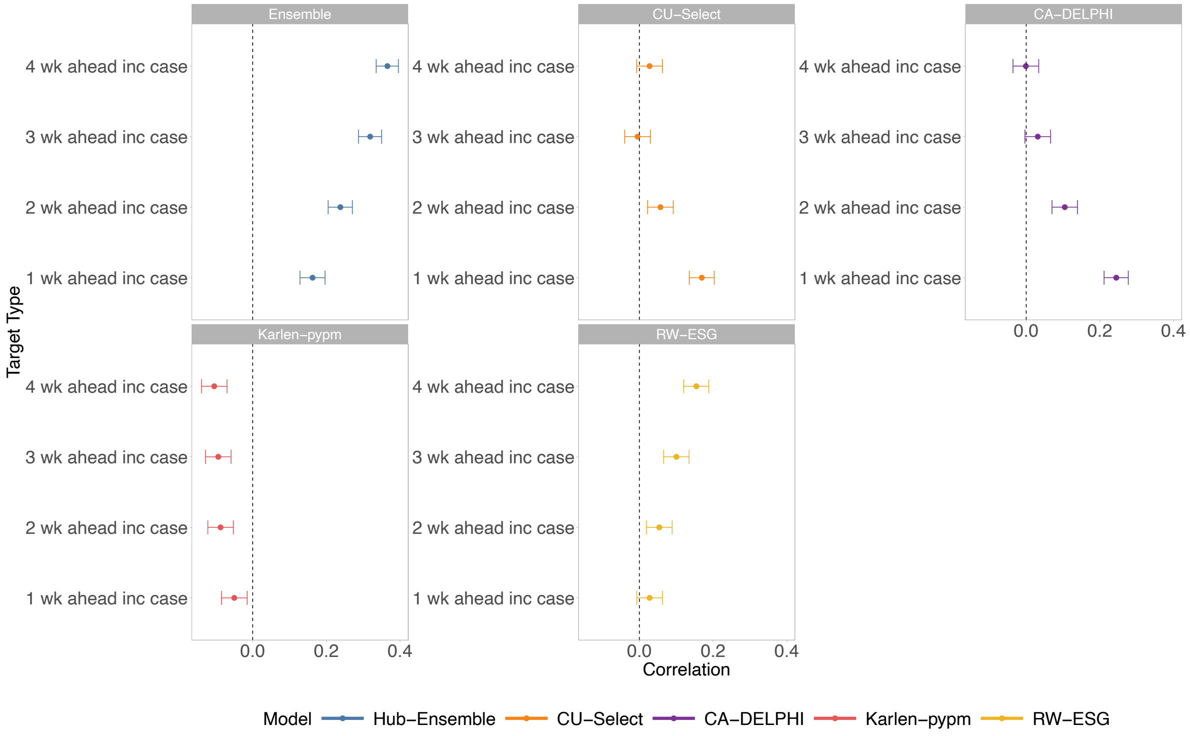


Figure SI 29: **Correlations between predictability and relative Weighted Interval Score (rWIS)** We visualise the Pearson's correlations (Table SI 6) between predictability and rWIS for each model and forecast target type, alongside the 95% Confidence Intervals (CIs) calculated using Fisher z-transformations.

Table SI 6: **Correlations between predictability and relative Weighted Interval Score (rWIS)** For each model and forecasted quantity, we report the Pearson's correlation $r$ between predictability and rWIS, alongside the 95% Confidence Intervals (CIs) calculated using Fisher z-transformations and the Bonferroni-adjusted p-value for the two-sided hypothesis test with null hypothesis that $r = 0$. The baseline values are intentionally marked as NA because it is the comparison model for rWIS.

| Model | Target Type | r | 95% CI | Bonferroni-adjusted $p$ |
|---|---|---|---|---|
| Ensemble | 1 wk ahead inc case | 0.16 | [0.13, 0.20] | P < 0.001 |
| Ensemble | 2 wk ahead inc case | 0.24 | [0.20, 0.27] | P < 0.001 |
| Ensemble | 3 wk ahead inc case | 0.32 | [0.29, 0.35] | P < 0.001 |
| Ensemble | 4 wk ahead inc case | 0.37 | [0.34, 0.40] | P < 0.001 |
| COVIDhub-baseline | 1 wk ahead inc case | NA | NA | NA |
| COVIDhub-baseline | 2 wk ahead inc case | NA | NA | NA |
| COVIDhub-baseline | 3 wk ahead inc case | NA | NA | NA |
| COVIDhub-baseline | 4 wk ahead inc case | NA | NA | NA |
| CU-select | 1 wk ahead inc case | 0.17 | [0.14, 0.20] | P < 0.001 |
| CU-select | 2 wk ahead inc case | 0.06 | [0.02, 0.09] | P = 0.0273 |
| CU-select | 3 wk ahead inc case | -0.01 | [-0.04, 0.03] | P = 1 |
| CU-select | 4 wk ahead inc case | 0.03 | [-0.01, 0.06] | P = 1 |
| CovidAnalytics-DELPHI | 1 wk ahead inc case | 0.24 | [0.21, 0.28] | P < 0.001 |
| CovidAnalytics-DELPHI | 2 wk ahead inc case | 0.10 | [0.07, 0.14] | P < 0.001 |
| CovidAnalytics-DELPHI | 3 wk ahead inc case | 0.03 | [-0.00, 0.07] | P = 1 |
| CovidAnalytics-DELPHI | 4 wk ahead inc case | -0.00 | [-0.04, 0.03] | P = 1 |
| Karlen-pypm | 1 wk ahead inc case | -0.05 | [-0.08, -0.02] | P = 0.0992 |
| Karlen-pypm | 2 wk ahead inc case | -0.09 | [-0.12, -0.05] | P < 0.001 |
| Karlen-pypm | 3 wk ahead inc case | -0.09 | [-0.13, -0.06] | P < 0.001 |
| Karlen-pypm | 4 wk ahead inc case | -0.10 | [-0.14, -0.07] | P < 0.001 |
| RobertWalraven-ESG | 1 wk ahead inc case | 0.03 | [-0.01, 0.06] | P = 1 |
| RobertWalraven-ESG | 2 wk ahead inc case | 0.05 | [0.02, 0.09] | P = 0.0483 |
| RobertWalraven-ESG | 3 wk ahead inc case | 0.10 | [0.07, 0.13] | P < 0.001 |
| RobertWalraven-ESG | 4 wk ahead inc case | 0.15 | [0.12, 0.19] | P < 0.001 |

#### G.6 Predictability and $R(t)$

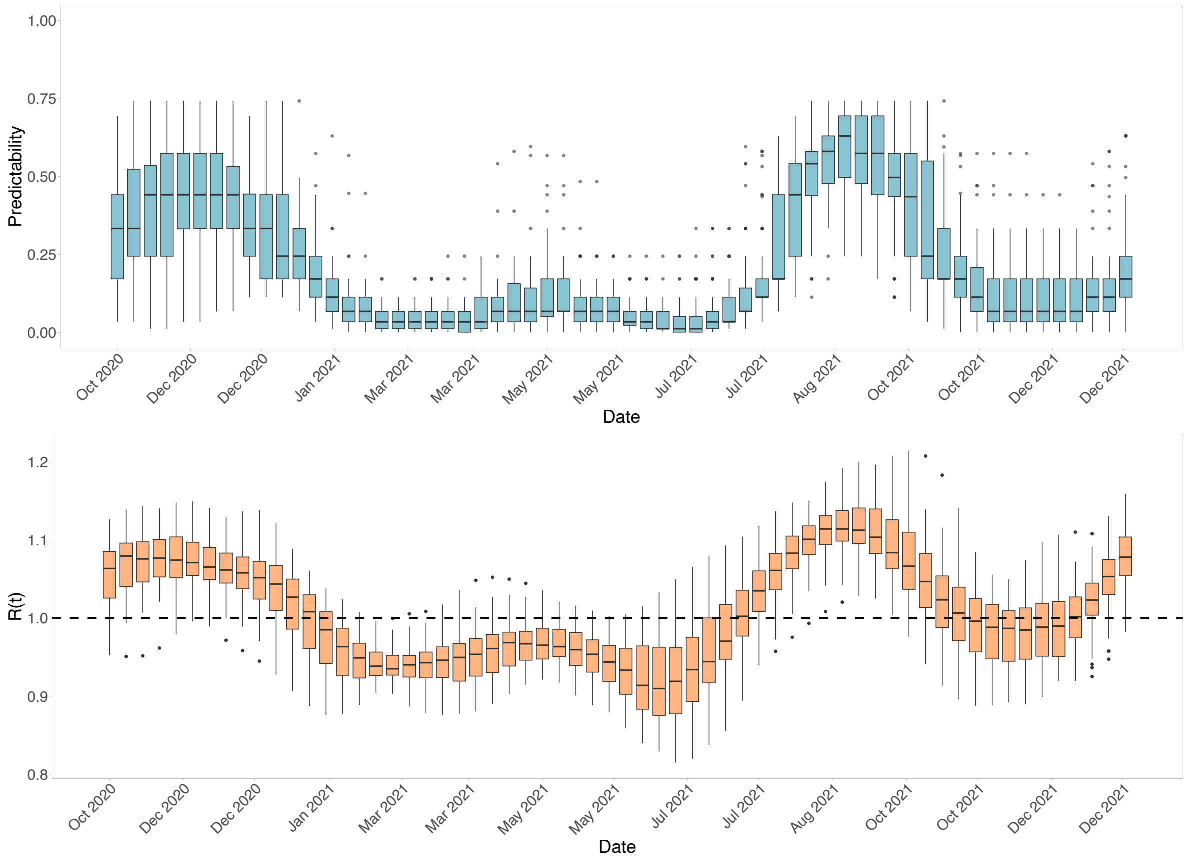


Figure SI 30: **Predictability and $R(t)$:** We visualise the boxplots of window-specific predictability for the observed state-level case data (top row) and mean time-varying reproduction number $R(t)$ (bottom row), which summarise the values (y-axis) obtained in each state in each 12-week trailing window, where the maximum date of each window is shown (x-axis). The box and whiskers cover the interquartile range, and points represent outliers.

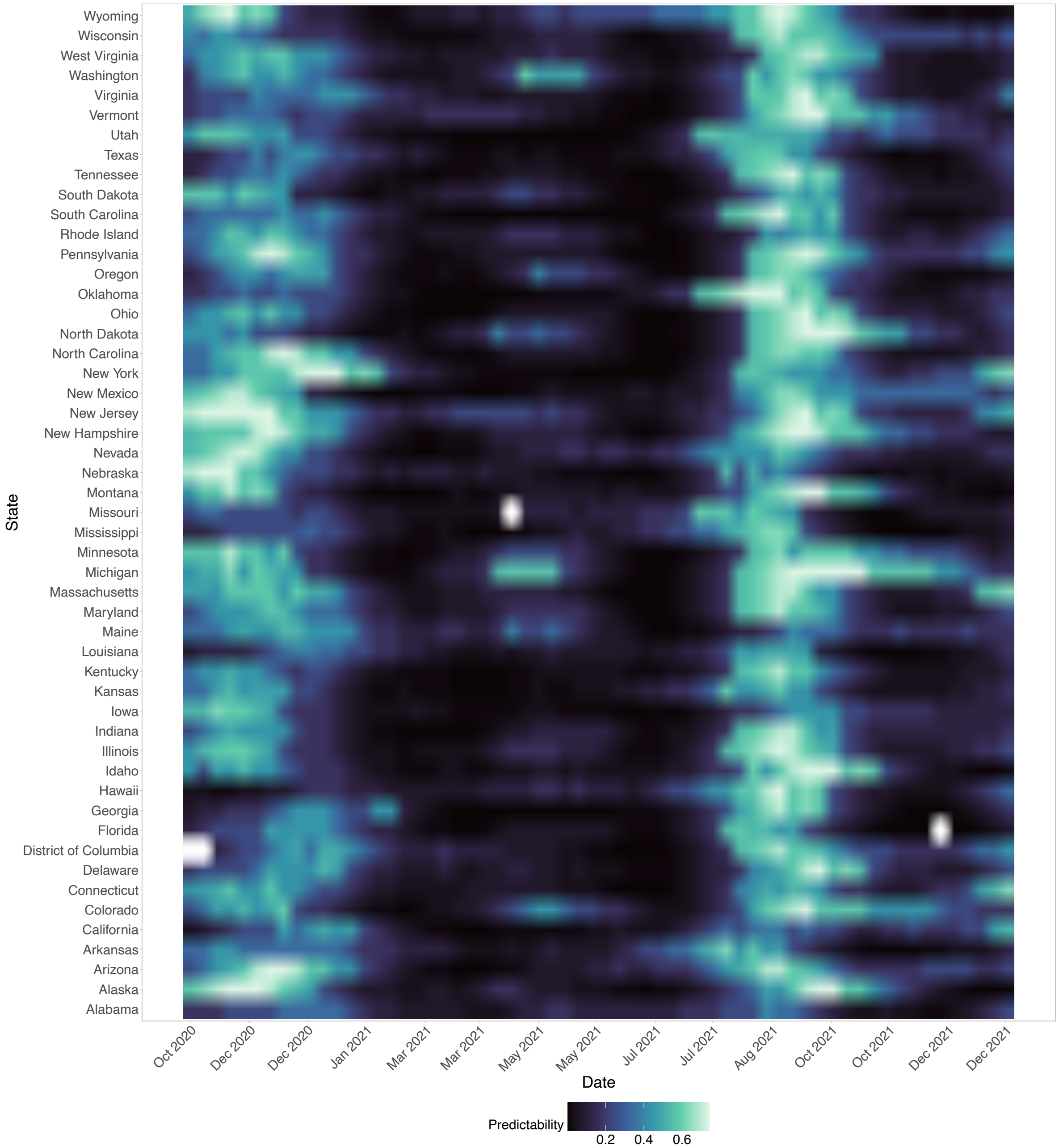


Figure SI 31: **Predictability over time and across space:** We visualise the window-specific predictability of each state (y-axis) in each 12-week trailing window, where the maximum date of each window is shown (x-axis).

### H COVID-19 application further analyses

#### H.1 Further details on overall evaluation results

We first evaluated the forecasts across the studied time period. Following our workflow (Figure 1), suppose a first policy question is the common task of predicting the future continuous value of weekly incident COVID-19 cases for each of the 51 studied states/territories. Using WIS and rWIS, for both forecast horizons of 1–4 weeks, the ensemble model generally outperformed other models, including across all time points (Figure SI 1 A), for individual locations (Figure SI 1 B), and in trailing windows of length 12 weeks (Figure SI 1 C, SI 3 – SI 6). Outperformance over time and across space was less clear for longer forecast horizons (Figure SI 1 C, Figures SI 5 – SI 6). As WIS approximates CRPS for quantile-represented forecasts, the ensemble model generally produced forecasts with the most value for a decision-maker with uniform risk preferences, when value was measured across all epidemic event thresholds (see Section 3).

These forecasts have already been extensively analysed through the lens of WIS [2, 16]. In epidemic settings, we are often interested in the many decisions for epidemic control faced by a specific decision-maker. A forecaster or evaluator can consider a risk-intolerant decision-maker with low C/L ratio ($\alpha$) – action is inexpensive, relative to epidemic losses. In weather forecasting, a typical range for extreme events considered is $\alpha$ between 0.01 and 0.2 [34, 68]. For both forecast horizons and a range of C/L ratios, according to the pinball loss and its CORP decomposition, the ensemble model produced simultaneously more discriminative and better calibrated forecasts (Figure 3 A). This indicates higher forecast value for the ensemble model for risk-intolerant decision-makers across the entire set of observed epidemic event thresholds. Such user-specific insights may be useful if the decision-maker could not, or did not wish to, define a specific epidemic event *a priori* (e.g., evolving dynamics), but had a fixed risk preference across event thresholds and over time. We further examined pinball loss for different decision-maker preferences over time (Figures SI 15–SI 16) and across space (Figures SI 18–SI 24), where the ensemble did not always outperform the baseline model for risk-intolerant decision-makers (e.g., Figure SI 15 and SI 18). However, for binary event thresholds, the ensemble was the only model to provide a generally greater forecast value over the baseline model across different forecast horizons and most C/L ratios, aligning with the findings for WIS across the predictive distribution. Compared to risk-intolerant decision-makers and longer horizons, we found greater relative value for more risk-tolerant decision-makers (higher C/L ratios) and for shorter forecast horizons.

Focusing on the axis of epidemic events (see Figure 2 B, right panel), we found that for classifying the upper 10% of each location's observed incident cases (location-specific event thresholds $\theta$), the CORP-based decomposition of the BS indicated similar results – the ensemble providing more discriminative and better calibrated forecasts for decision-makers (Figure 3B). This event-specific analysis focuses on state-specific extreme events for a uniform family of decision-makers, yet ignores any likely risk preferences of the decision-maker that could expressed with a range of plausible C/L ratios, especially for extreme events.

User-specific (Figure 3 C.1) and event-specific (Figure 3 C.2) Murphy diagrams allow simultaneous assessment of forecast value for different epidemic events (or decision-makers) and a specific decision-maker (or a specific epidemic event). The Murphy diagrams for specific decision-makers (Figure 3 C.1, holding $\alpha$ fixed) trace a trend of the Karlen-pypm and ensemble model's forecasts often providing the most value for risk-intolerant decision-makers with low C/L ratios (for whom action is inexpensive, relative to the loss). We found less clear trends for rarer, larger event thresholds ($\theta$) across forecast horizons. For more risk-tolerant decision-makers (i.e., higher C/L ratios), the ensemble model provided clearer value for longer forecast horizons (Figure SI 26). These user-specific Murphy diagrams, while crowded and likely benefitting from further focus on event thresholds $\theta$ for specific policy questions, capture the value of the models' forecasts for informing a specific decision-maker's binary decision of whether to act based on the forecasted probability of the future (one or four weeks ahead) weekly incidence exceeding different event thresholds ($\theta$). Focusing on location-specific extrema (location-specific event thresholds $\theta$), the event-specific Murphy diagrams for BS (Figure 3 C.2) display that the ensemble model provided the most value for classifying the upper 10% of each location's incident cases for most decision-makers across both forecast horizons (up to C/L ratios of 0.75). There is benefit in using both types of Murphy diagram, as the user- and event- specific visualisations can capture forecast value when there is uncertainty in the defined event (e.g., evolving dynamics) and/or decision-maker's C/L ratio (e.g., likely uncertainty about cost of intervention or epidemic event loss). This can inform decision-making under different uncertainties.

The REV does not capture forecast performance in terms of elementary scores, but provides an alternative, interpretable measure of relative value for different forecasting models over other simple, operational models. The choice of reference model is crucial, and we used i) a probabilistic classifier that always predicts the binary event to occur with a fixed probability (similar to the climatology commonly used in weather forecasting [10, 34]), and ii) the COVID-19 Forecast Hub baseline model [39].

Using the REV for the upper 10% of each location's incident cases (location-specific event thresholds $\theta$), relative to a constant probabilistic classifier (forecasting occurrence of a binary event with probability 0.1, Figure 3 C.3 top row), we found that i) almost all models provided relative value for a range of decision-maker risk preferences, ii) REV was generally greater for shorter horizons, and iii) the ensemble model (alongside the Karlen-pypm model for one-week-ahead forecasts) provided the most clear relative value across the range of risk preferences. Although the relative value of models' forecasts was often lower when using the baseline model as reference, the ensemble model again provided the most clear relative value, including for risk-tolerant decision-makers (high C/L ratios). We repeated these REV analyses for different types of events, such as the upper 10% (15%) of all observed incident cases (Figures SI 11–SI 12) and for the upper 15% of each location's observed incident cases (Figures SI 9–SI 10 and Figures SI 13–SI 14), and found largely similar results (e.g., highest REV for these same two models and similar trends across C/L ratios and forecast horizons). In practice, the results from these REV diagrams could be filtered by a forecaster or evaluator to interpret them for a specific decision-maker's defined event, forecast horizon, and available action.

Relating forecast performance to predictability, higher predictability was moderately yet significantly (Table SI 4) correlated with higher $R^2$ for each model's forecasts across different horizons (Figures 4 and SI 27). Using relative measures such as rWIS (Figures SI 32, SI 29) or relative pinball loss across different C/L ratios (Figures 4 and SI 25), we estimated that several forecasting models generally had *greater* relative performance when epidemic dynamics were *less* predictable. However, greater relative performance for the given model during times of lower predictability, compared to times of high predictability, does not mean that the models necessarily provided improvements over a naive/baseline model at such times (Figure 4). It is necessary to understand their overall predictive performance during all regimes of predictability. We interpret the relative improvements alongside comparisons to the baseline (rWIS and relative pinball loss alongside predictability, Figures 4). We estimated generally consistent benefits, across space, time, and metrics, of using the ensemble model over the naive baseline model, for decision-making during less *and* more predictable times. The estimated relative improvements of the ensemble model, during high unpredictability, were often greater at longer forecast horizons (Figures SI 15 – SI 29).

These findings shed light on how predictability could have impacted the value of different models, yet we also considered whether such predictability itself can be anticipated and/or what drives different predictability regimes. Higher predictability within a window was strongly correlated with higher $R(t)$ estimates within the window and in past windows, while higher predictability was correlated with higher levels of incident cases in the recent past (Figures 4, SI 30). This indicates that predictability was greatest during the epidemic growth and peak phases (as classified by [16]), and lower during the declining and nadir phases. Overall, we found generally low predictability, and this varied across space and time (Figures SI 30 – SI 31). These insights, while needing further exploration beyond our scope of evaluation, may be useful for informing model development and selection and for anticipating the likely future reliability of outputs. For example, lower $R(t)$ preceding lower predictability, or past performance during growth or declining phases, may be used to guide changes to a model or to an ensemble's weights. In future works, the recent past and likely future predictability could be incorporated into the forecasting models or into the model selection procedures. For instance, we found that for the ensemble and several models, higher predictability was correlated with lower coverage of central 95% prediction intervals (PIs) (Figure 4, Table SI 5), particularly for longer forecast horizons (Figure SI 5). This reflects findings from [16] where coverage dropped during growth phases which we estimated to be generally more predictable. During such phases, central 95% PI coverage, even from the ensemble's forecasts, was often much lower than 95%, suggesting a need for updating of ensemble weights and/or models and their inputs (e.g., data collection).

#### H.2 Results for local, real-time decision-making

Above, we evaluated forecast quality and value *across* the 77 weeks. In practice, forecasters and evaluators will apply the framework for local decision-makers in specific locations at specific times. It is so far less clear how the forecasts could have informed decisions in real time for location-specific, important stages of the epidemic. Suppose now that a decision-maker in a given location is unknowingly at the onset of a large wave, which we define as the first date in 2021 when the median estimate of $R(t) > 1$. We focus on Arizona (a randomly chosen state).

Before the wave, predictability had been high during a previous wave and low at recent low case levels (Figure SI 32 B). High predictability had been moderately-to-strongly correlated with higher $R(t)$ ($r = 0.57$, 95% CI: 0.29, 0.76). Based on overall predictive performance (rWIS, Figure SI 32 B), the ensemble and Karlen-pypm forecasting models were likely the most suitable forecasting models to use going forward. Forecast value for specific events was generally higher for these models when measured using the BS, with superior discrimination and calibration properties (Figure SI 33 A), yet the "best" model depended on different decision-maker risk preferences (via C/L ratios), event thresholds, and forecast horizon (Figures SI 36 A, event-specific Murphy diagrams of Figure SI 34 A, or user-specific Murphy diagrams of Figure SI 35 A). The ensemble model generally provided the most value before the wave for one-week-ahead forecasts across all event thresholds for different decision-maker preferences (Figure SI 36 A), yet for four-weeks-ahead forecasts, the RW-ESG and Karlen-pypm model provided more clear value for risk-tolerant (high C/L ratio) and risk-intolerant (low C/L ratio) respectively (Figure SI 36 A). Compared to the baseline model, the ensemble provided the most consistent clear relative value for both risk-intolerant (Figure SI 37) and risk-tolerant (Figure SI 38) over time.

We found similar results for the pre-wave period, compared to the entire studied period (Section 6.2), with many models having higher $R^2$ (Figures SI 39, SI 41, Table SI 4) and lower 95% PI coverage (Figures SI 39, SI 43, Table SI 9) during times of greater predictability, which is what marks the onset of this wave, Figure SI 32 A). During such high predictability regimes, the ensemble offered value over the baseline model for risk-tolerant and risk-intolerant decision-makers (pinball loss, Figures SI 39, SI 37–SI 38), yet offered comparably less value (over the baseline) during times of lower predictability (rWIS and relative pinball loss, Figure SI 39). Before the wave, only the Karlen-pypm model had regularly outperformed the baseline in terms of overall predictive performance (rWIS) across both high and low predictability regimes, yet its relative value varied between risk-intolerant and risk-tolerant decision-makers.

Studying the large wave, rWIS results for one-week-ahead forecasts remain comparable to pre-wave results. The same two models (Ensemble and Karlen-pypm) provided the best overall predictive performance, yet no model consistently outperformed the baseline for four-weeks-ahead forecasts (Figure SI 32 B). Focusing on specific events, co-defining *a priori* the policy question and epidemic event to measure before the wave is non-trivial and infinite choices exist. First, a decision-maker may be interested in whether future incident cases at a specified horizon breach a fixed extreme threshold, say $\theta = 20,000$ (an extreme value *before* the wave, Figure SI 32 A). The decision problem for this event could correspond to taking action based on the costs and preventable losses of maintaining a public health measure (e.g., additional ICU capacity, see Section 3.2) if incident cases $y \geq 20,000$. Using the CORP-based decomposition of the mean BS for this event (Figure SI 32 C, left), both the ensemble and CU-select model provided the most value across discrimination ability and calibration properties. However, if we add a decision-maker's risk preferences (Figure SI 32 C, middle), the event-specific Murphy diagrams capture how the optimal model varied by risk preferences. The analogous REV plots (Figure SI 32 C, right) are also crowded, yet illustrate differences in selected models across risk preferences and the low value for longer forecast horizons. In practice, we could provide the operational forecast value of each model for a specific decision-maker using a plausible range of their C/L ratios for this event threshold ($y \geq 20,000$).

On the other hand, an event threshold may become less meaningful if dynamics are evolving quickly and/or incident cases are predictably above/below this threshold. Varying this epidemic event threshold ($\theta$), the user-specific Murphy diagram for pinball loss (Figure SI 32 D, right panel) captures how well the forecasts informed decisions using different event thresholds. These diagrams can also easily become crowded (e.g., due to a wide range of events on the x-axis, see Figure SI 32 D). Alternatives may be to focus on the CORP-based decompositions for BS or pinball loss (Figure SI 32 C–D) or to restrict x-axes of the Murphy diagram to event thresholds ($\theta$) of interest.

Suppose a decision-maker cannot pre-define a fixed event threshold and/or will not base decisions on a single event threshold, yet has a fixed risk profile (C/L ratio $\alpha$). Focusing on *all* observations of the wave for these fixed risk preferences, the pinball loss decompositions (Figures SI 32 D and SI 36) capture how the ensemble provided the clearest value for risk-intolerant decision-makers. However, the chosen model for decision-makers varied according to their relative cost of mitigation (C/L ratio) and the forecast horizon. The models with the most value for a given decision-maker during the wave (Figure SI 36 B) often differed to the models with the most value before (Figure SI 36 A) the wave. We found similar differences in models selected before versus during the wave when using the Brier score, REV, and relative pinball loss (Figure SI 37–SI 38). Collectively, these results suggest that for policy-focused model development and representative evaluation, forecasters and evaluators need to i) zoom in on user- and event-specific measures and ii) capture the spatiotemporal structures of epidemic data in evaluation and model selection. Otherwise, averaging over these heterogeneities may result in inaccurate measures of local, real-time forecast value and a failure to provide different decision-makers with the optimal model's forecasts.

Retrospectively, we can analyse how well the peak values were predicted, for example, using BS or REV for the upper 10% of incident cases during the wave. This assessment of wave-specific extreme events can be used by forecasters and/or evaluators to build better models going forward and/or to assign ensemble weights. Here, the best models across the C/L ratios for predicting these events again varied by different forecast horizons (Figure SI 32 E, right) and by different decision-makers' preferences (Figure SI 32 E, left), while the best models differed to those chosen for the upper 10% of events before the wave (Figure SI 33).

Compared to the pre-wave results, similar predictability trends persisted during the wave (Figures SI 32 A and SI 39–SI 40), as predictability rose and fell alongside the epidemic trajectory – $R(t)$ was strongly correlated with estimated predictability ($r = 0.80$, 95% CI: (0.62, 0.91)) during the wave. Low predictability was again correlated with lower pointwise accuracy for models, and during such low predictability regimes, some models had greater value relative to the baseline model for the uniform family of decision-maker preferences, but not necessarily individual decision-maker preferences (Figures SI 33 –SI 38 and Figure SI 40). Therefore, given that sharp predictability changes were also estimated towards the start and at the peak of the wave, it is perhaps unsurprising that the best models varied (Figures SI 33 – SI 38) i) for different decision-makers, ii) for different events, iii) over time within the wave, and iv) compared to the pre-wave period. Looking forward, these insights (e.g., predictability having consistent impacts on forecast performance), which benefit from our hindsight, suggest the need for model development and selection, ensemble weighting, and data collection to condition on predictability, in addition to time, space, and the decision-maker's preferences and priorities.

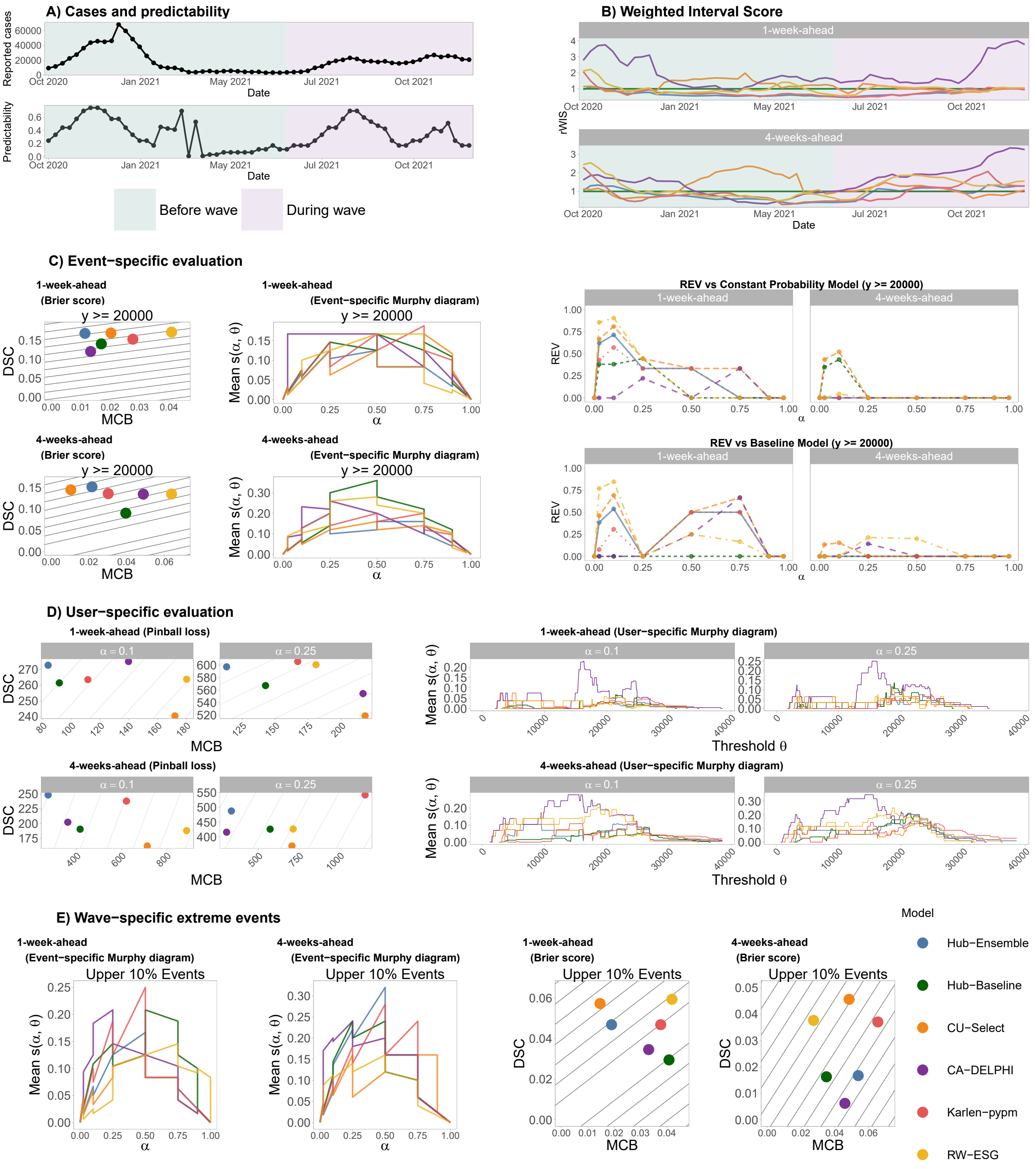


Figure SI 32: **Evaluation for real-time decision-making during a COVID-19 wave in Arizona:** **A)** Reported incident cases and estimated predictability before and during a COVID-19 wave that started in July 2021 (based on median $R(t) > 1$) and ended in January 2022 (based on the end of the dataset). **B)** 12-week trailing window relative Weighted Interval Score (rWIS) for all models across two forecast horizons before and during the wave. **C)** For two forecast horizons, Brier Score decomposition and event-specific Murphy diagrams, alongside a Relative Economic Value diagram, where the event is pre-defined as incident cases $y \geq 20{,}000$ (based on values before the wave). **D)** For two forecast horizons, Brier Score decomposition and event-specific Murphy diagrams for risk-intolerant decision-makers. We provide further visualisations for other risk-intolerant decision-maker preferences and risk-tolerant decision-makers in Figures SI 35 – SI 38. **E)** Event-specific Murphy diagrams and Brier Score decomposition and event-specific Murphy diagrams for forecasting the upper 10% of incident cases during this COVID-19 wave.

#### H.3 Brier score decomposition before and during wave

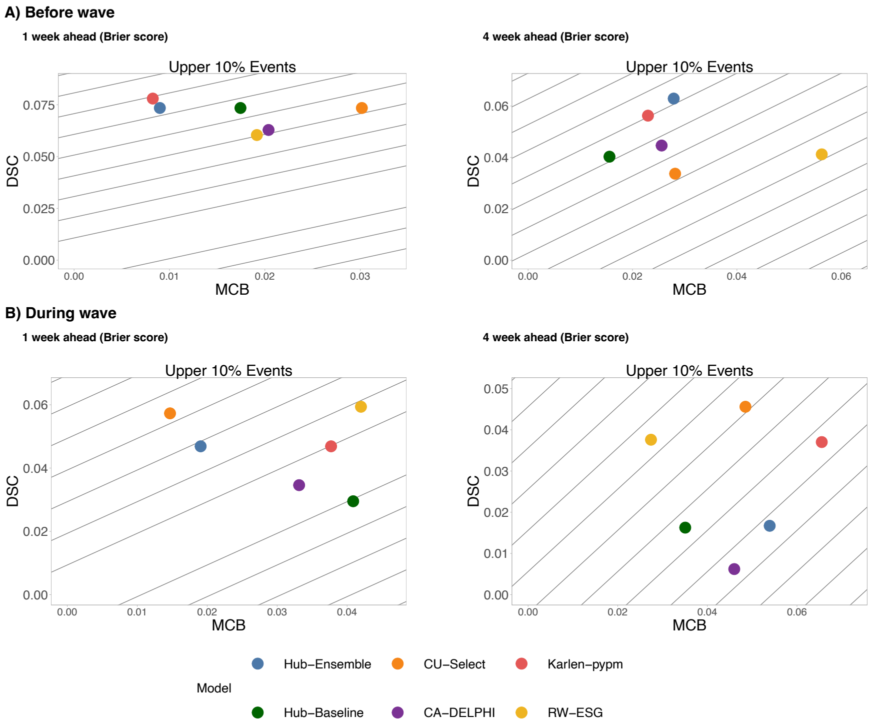


Figure SI 33: **Brier score decomposition for the upper 10% of cases before and during wave:** For the state of Arizona, we visualise the discrimination ability (DSC) component (y-axis) and miscalibration (MCB) component (x-axis) of each model's Brier score for the upper 10% of cases before ((**A**)) and during ((**B**)) the COVID-19 wave which started on 26 June 2021 (when the median $R(t)$ estimate was greater than 1). Higher DSC values and lower MCB values indicate greater forecast value for a the event (i.e., event probability/frequency $\pi(\theta) = 0.1$) across the set of observations.

#### H.4 Event-specific Murphy diagrams before and during wave

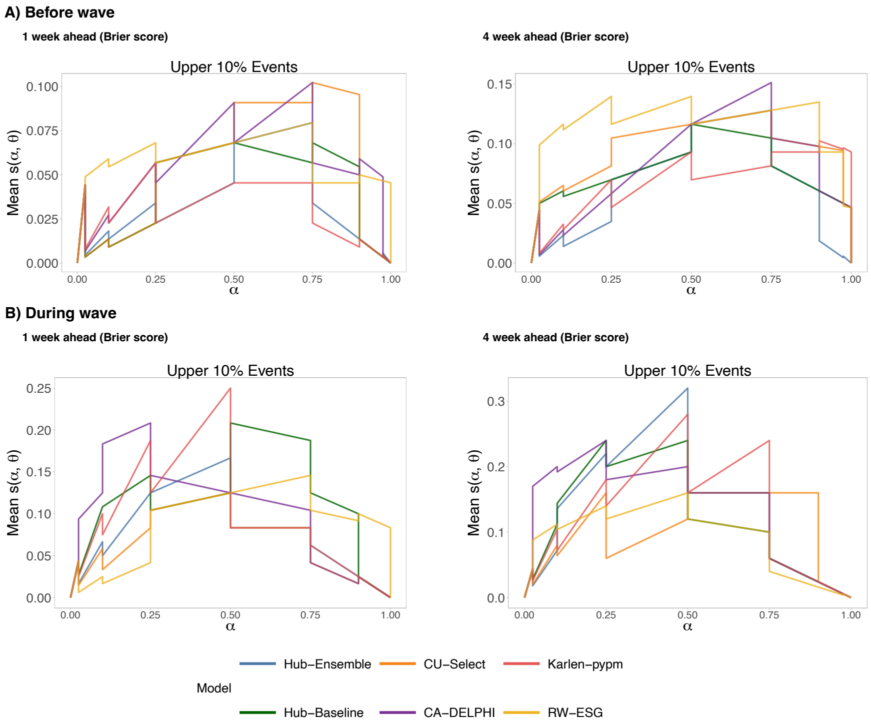


Figure SI 34: **Event-specific Murphy diagrams for the upper 10% of cases before and during wave:** For the state of Arizona, we visualise the Murphy diagrams relevant to specific decision-makers before the studied wave. Each Murphy curve captures mean elementary score $s(\alpha, \theta)$ for a fixed event threshold $(\theta)$ and varying C/L ratios $(\alpha)$. Lower values on Murphy curves indicate superior performance of a model's forecasts for the given user-event combination.

#### H.5 User-specific Murphy diagrams before and during wave

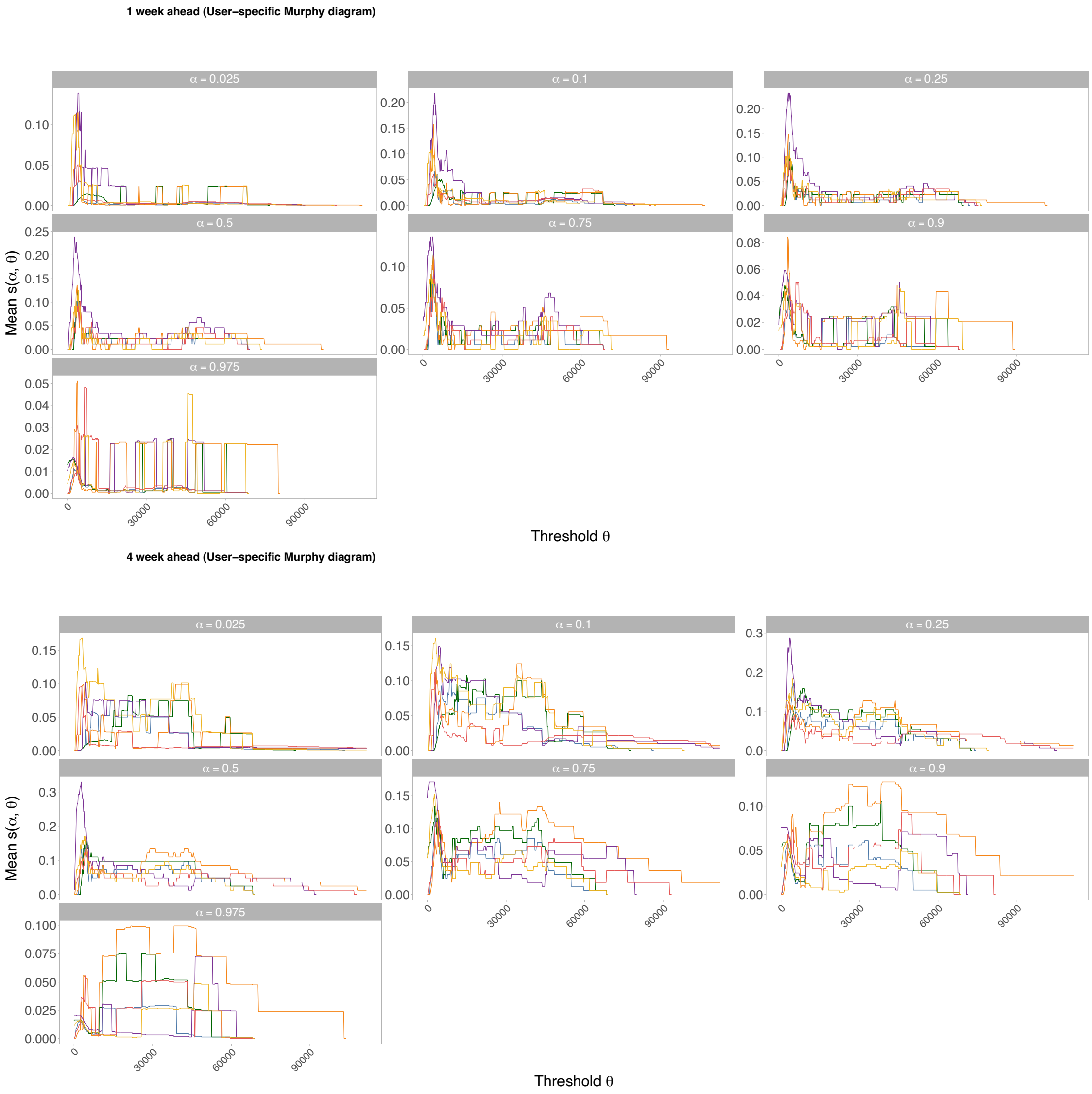


Figure SI 35: **User-specific Murphy diagrams before wave:** For the state of Arizona, we visualise the Murphy diagrams relevant to specific decision-makers before the studied COVID-19 wave which started on 26 June 2021 (when the median $R(t)$ estimate was greater than 1). Each Murphy curve captures mean elementary score $s(\alpha, \theta)$ for a fixed C/L ratio ($\alpha$) and varying event thresholds ($\theta$). Lower values on Murphy curves indicate superior performance of a model's forecasts for the given user-event combination.

#### H.6 Pinball loss decomposition before and during wave

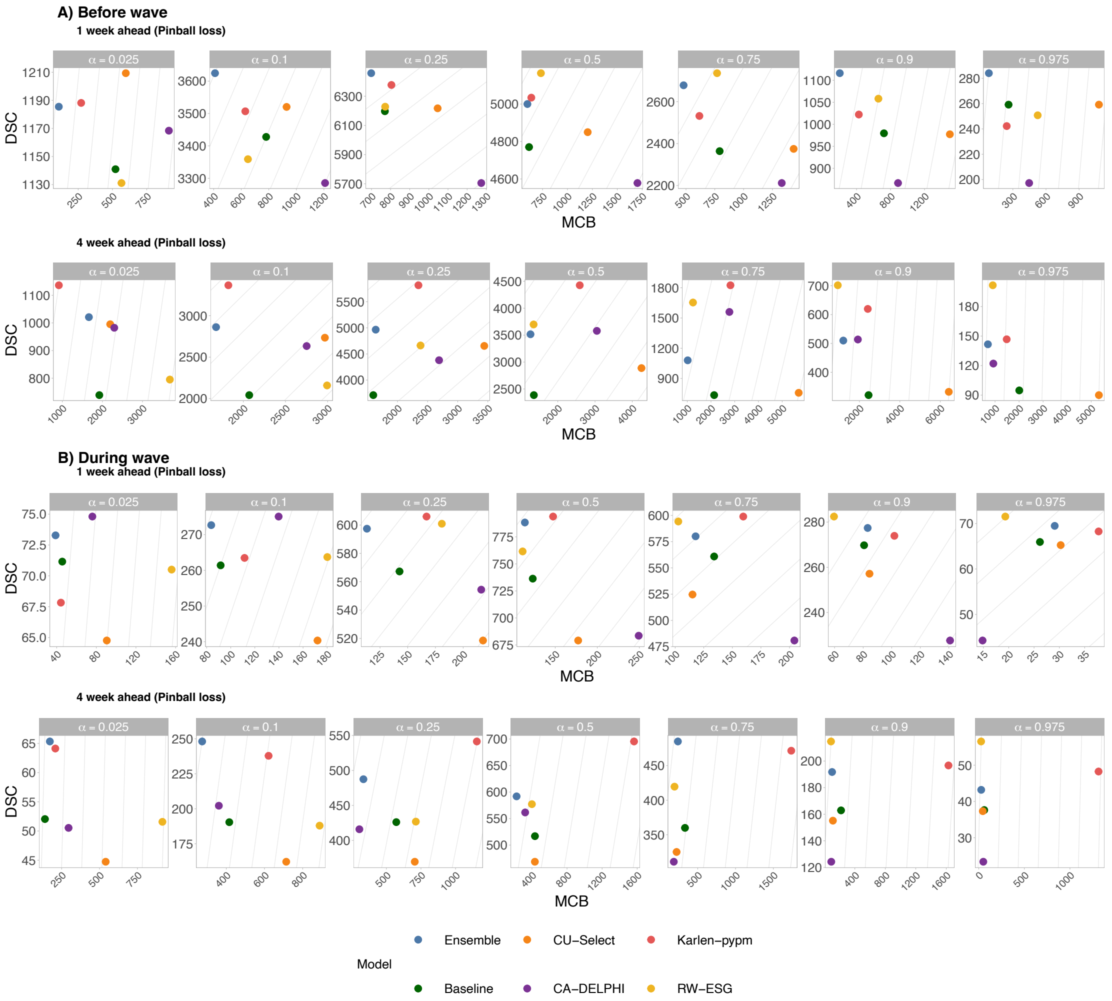


Figure SI 36: **Pinball loss decomposition before and during wave:** For the state of Arizona, we visualise the discrimination ability (DSC) component (y-axis) and miscalibration (MCB) component (x-axis) of each model's pinball loss before ((**A**)) and during ((**B**)) the COVID-19 wave which started on 26 June 2021 (when the median $R(t)$ estimate was greater than 1). Higher DSC values and lower MCB values indicate greater forecast value for a specific decision-maker's C/L ratio $\alpha$ across the set of observations.

#### H.7 Relative pinball loss before and during wave

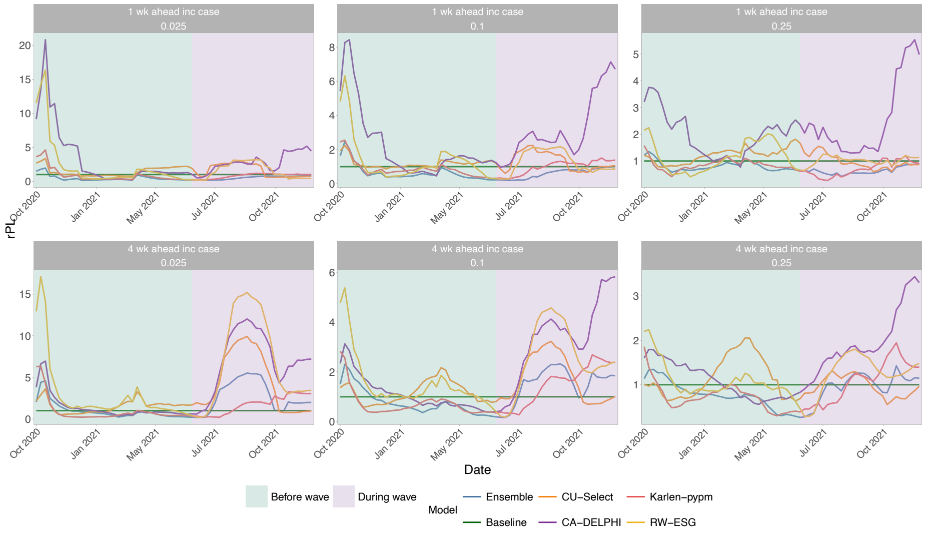


Figure SI 37: **Relative pinball loss over time for risk-intolerant decision-makers before and during wave:** For the state of Arizona, we visualise each model's relative pinball loss (rPL) (y-axis) before and during the COVID-19 wave which started on 26 June 2021 (when the median $R(t)$ estimate was greater than 1). rPL was calculated using the COVID-19 Forecast Hub baseline model in each 12-week trailing window, where the maximum date of each window is shown (x-axis). Values of rPL < 1 indicate greater relative forecast value of a given model, compared to the baseline model, for observations across that 12-week window and specific decision-maker's preferences (C/L ratio, $\alpha$). We focus on risk-tolerant decision-makers in Figure SI 38

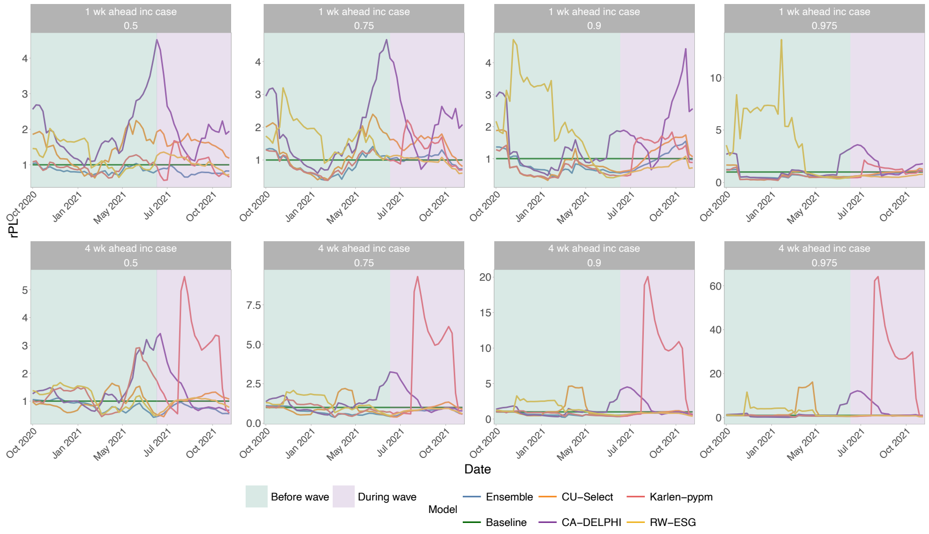


Figure SI 38: **Relative pinball loss over time for risk-tolerant decision-makers before and during wave:** For the state of Arizona, we visualise each model's relative pinball loss (rPL) (y-axis) before and during the COVID-19 wave which started on 26 June 2021 (when the median $R(t)$ estimate was greater than 1). rPL was calculated using the COVID-19 Forecast Hub baseline model in each 12-week trailing window, where the maximum date of each window is shown (x-axis). Values of rPL < 1 indicate greater relative forecast value of a given model, compared to the baseline model, for observations across that 12-week window and specific decision-maker's preferences (C/L ratio, $\alpha$). We focus on risk-intolerant (lower C/L ratio) decision-makers in Figure SI 37

#### H.8 Predictability impacts before and during wave

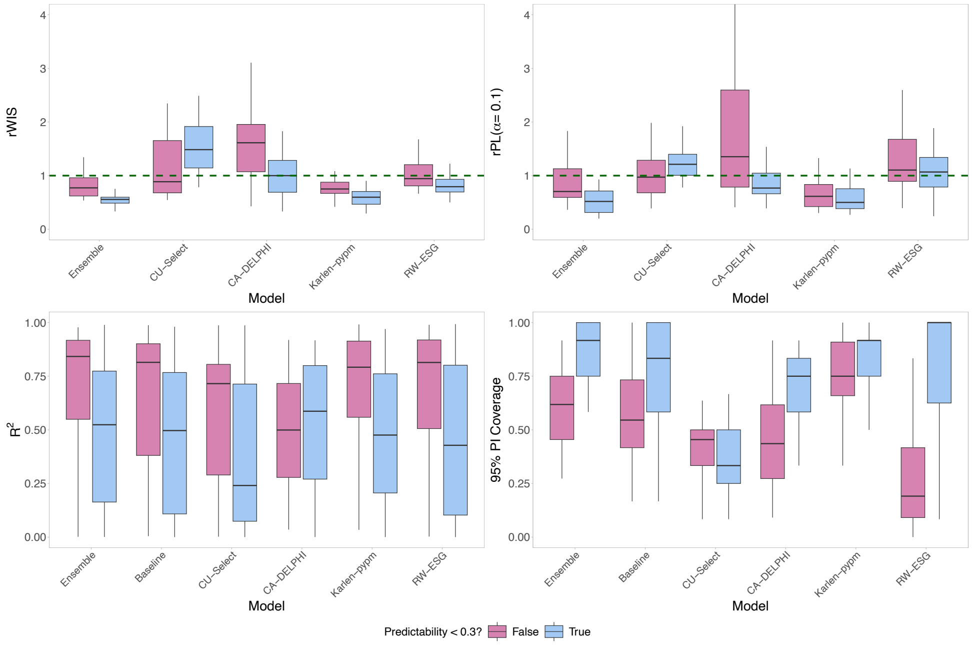


Figure SI 39: **Pre-wave predictability and forecast performance:** For the state of Arizona, we visualise the relationship between forecast performance metrics and predictability before the onset of the studied wave (starting July 2021).

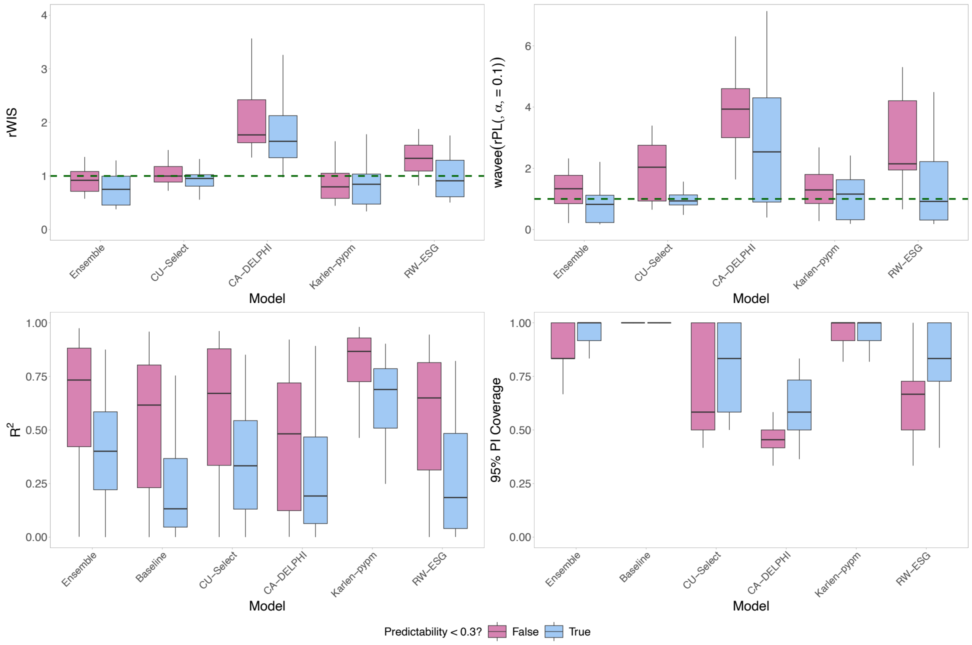


Figure SI 40: **During-wave predictability and forecast performance:** For the state of Arizona, we visualise the relationship between forecast performance metrics and predictability during the studied wave (starting July 2021).

#### H.9 Predictability versus forecast metrics

##### H.9.1 Predictability versus $R^2$

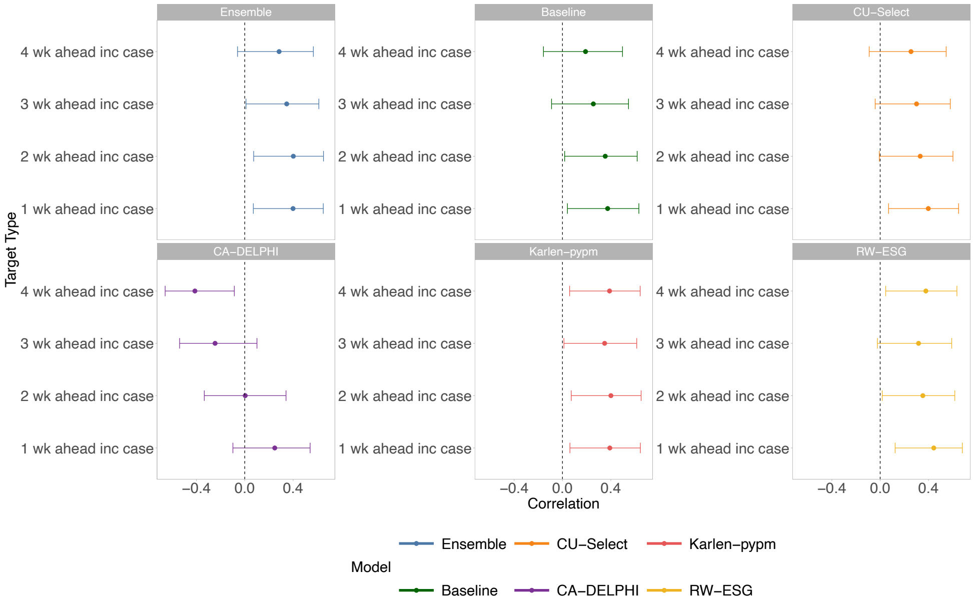


Figure SI 41: **Pre-wave correlations between predictability and $R^2$:** In Arizona before the onset of the studied wave (starting July 2021), we visualise the Pearson's correlations (Table SI 4) between predictability and $R^2$ for each model and forecast target type, alongside the 95% Confidence Intervals (CIs) calculated using Fisher z-transformations.

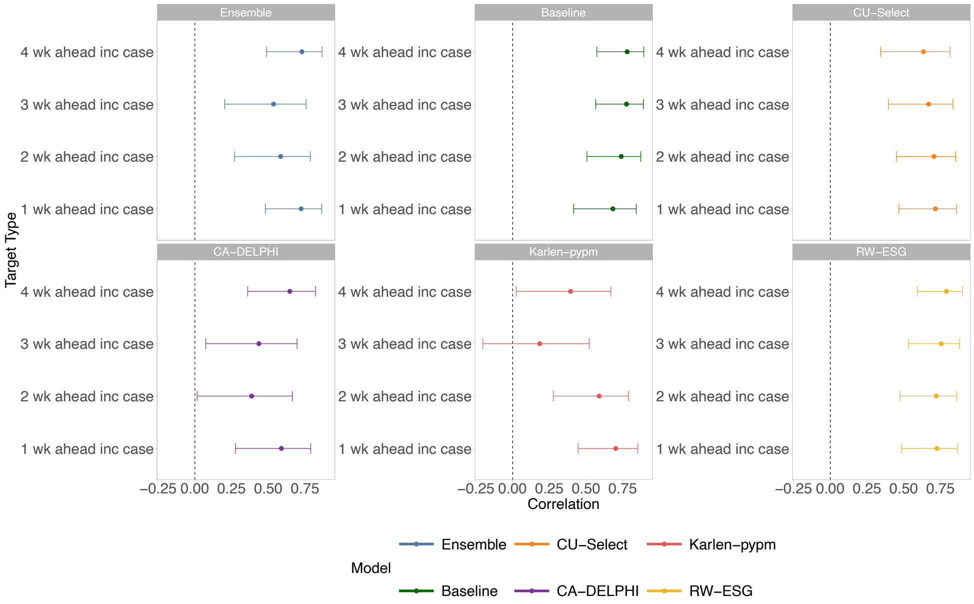


Figure SI 42: **During-wave correlations between predictability and $R^2$:** In Arizona during the studied wave (starting July 2021), we visualise the Pearson's correlations (Table SI 4) between predictability and $R^2$ for each model and forecast target type, alongside the 95% Confidence Intervals (CIs) calculated using Fisher z-transformations.

Table SI 7: **Pre-wave correlations between predictability and $R^2$:** In Arizona before the onset of the studied wave (starting July 2021), for each model and forecasted quantity, we report the Pearson's correlation $r$ between predictability and $R^2$, alongside the 95% Confidence Intervals (CIs) calculated using Fisher z-transformations and the Bonferroni-adjusted p-value for the two-sided hypothesis test with null hypothesis that $r = 0$.

| Model | Target Type | r | 95% CI | Bonferroni-adjusted $p$ |
|---|---|---|---|---|
| Ensemble | 1-week-ahead | 0.40 | [0.07, 0.65] | P = 0.461 |
| Ensemble | 2-weeks-ahead | 0.40 | [0.07, 0.65] | P = 0.442 |
| Ensemble | 3-weeks-ahead | 0.35 | [0.01, 0.61] | P = 1 |
| Ensemble | 4-weeks-ahead | 0.28 | [-0.06, 0.57] | P = 1 |
| COVIDhub-baseline | 1-week-ahead | 0.37 | [0.04, 0.63] | P = 0.703 |
| COVIDhub-baseline | 2-weeks-ahead | 0.35 | [0.02, 0.62] | P = 0.959 |
| COVIDhub-baseline | 3-weeks-ahead | 0.26 | [-0.09, 0.55] | P = 1 |
| COVIDhub-baseline | 4-weeks-ahead | 0.19 | [-0.16, 0.50] | P = 1 |
| CU-select | 1-week-ahead | 0.40 | [0.07, 0.65] | P = 0.473 |
| CU-select | 2-weeks-ahead | 0.33 | [-0.01, 0.60] | P = 1 |
| CU-select | 3-weeks-ahead | 0.30 | [-0.04, 0.58] | P = 1 |
| CU-select | 4-weeks-ahead | 0.26 | [-0.09, 0.55] | P = 1 |
| CovidAnalytics-DELPHI | 1-week-ahead | 0.25 | [-0.10, 0.54] | P = 1 |
| CovidAnalytics-DELPHI | 2-weeks-ahead | 0.00 | [-0.33, 0.34] | P = 1 |
| CovidAnalytics-DELPHI | 3-weeks-ahead | -0.25 | [-0.54, 0.10] | P = 1 |
| CovidAnalytics-DELPHI | 4-weeks-ahead | -0.41 | [-0.66, -0.09] | P = 0.369 |
| Karlen-pypm | 1-week-ahead | 0.39 | [0.06, 0.64] | P = 0.525 |
| Karlen-pypm | 2-weeks-ahead | 0.40 | [0.07, 0.65] | P = 0.451 |
| Karlen-pypm | 3-weeks-ahead | 0.35 | [0.01, 0.61] | P = 1 |
| Karlen-pypm | 4-weeks-ahead | 0.39 | [0.06, 0.64] | P = 0.546 |
| RobertWalraven-ESG | 1-week-ahead | 0.44 | [0.12, 0.68] | P = 0.207 |
| RobertWalraven-ESG | 2-weeks-ahead | 0.35 | [0.02, 0.62] | P = 0.974 |
| RobertWalraven-ESG | 3-weeks-ahead | 0.32 | [-0.02, 0.59] | P = 1 |
| RobertWalraven-ESG | 4-weeks-ahead | 0.38 | [0.05, 0.63] | P = 0.66 |

Table SI 8: **During-wave correlations between predictability and $R^2$:** In Arizona before the onset of the studied wave (starting July 2021), for each model and forecasted quantity, we report the Pearson's correlation $r$ between predictability and $R^2$, alongside the 95% Confidence Intervals (CIs) calculated using Fisher z-transformations and the Bonferroni-adjusted p-value for the two-sided hypothesis test with null hypothesis that $r = 0$.

| Model | Target Type | r | 95% CI | Bonferroni-adjusted $p$ |
|---|---|---|---|---|
| Ensemble | 1-week-ahead | 0.72 | [0.48, 0.86] | P < 0.001 |
| Ensemble | 2-weeks-ahead | 0.58 | [0.27, 0.79] | P = 0.0265 |
| Ensemble | 3-weeks-ahead | 0.54 | [0.20, 0.76] | P = 0.0794 |
| Ensemble | 4-weeks-ahead | 0.73 | [0.49, 0.87] | P < 0.001 |
| COVIDhub-baseline | 1-week-ahead | 0.68 | [0.42, 0.84] | P = 0.00151 |
| COVIDhub-baseline | 2-weeks-ahead | 0.74 | [0.51, 0.87] | P < 0.001 |
| COVIDhub-baseline | 3-weeks-ahead | 0.78 | [0.57, 0.89] | P < 0.001 |
| COVIDhub-baseline | 4-weeks-ahead | 0.78 | [0.57, 0.89] | P < 0.001 |
| CU-select | 1-week-ahead | 0.72 | [0.47, 0.86] | P < 0.001 |
| CU-select | 2-weeks-ahead | 0.71 | [0.45, 0.85] | P < 0.001 |
| CU-select | 3-weeks-ahead | 0.67 | [0.39, 0.83] | P = 0.00235 |
| CU-select | 4-weeks-ahead | 0.63 | [0.34, 0.81] | P = 0.00691 |
| CovidAnalytics-DELPHI | 1-week-ahead | 0.59 | [0.28, 0.79] | P = 0.0235 |
| CovidAnalytics-DELPHI | 2-weeks-ahead | 0.39 | [0.02, 0.66] | P = 1 |
| CovidAnalytics-DELPHI | 3-weeks-ahead | 0.44 | [0.07, 0.70] | P = 0.492 |
| CovidAnalytics-DELPHI | 4-weeks-ahead | 0.65 | [0.36, 0.82] | P = 0.00489 |
| Karlen-pypm | 1-week-ahead | 0.70 | [0.45, 0.85] | P < 0.001 |
| Karlen-pypm | 2-weeks-ahead | 0.59 | [0.28, 0.79] | P = 0.0232 |
| Karlen-pypm | 3-weeks-ahead | 0.18 | [-0.20, 0.52] | P = 1 |
| Karlen-pypm | 4-weeks-ahead | 0.39 | [0.03, 0.67] | P = 0.907 |
| RobertWalraven-ESG | 1-week-ahead | 0.73 | [0.48, 0.87] | P < 0.001 |
| RobertWalraven-ESG | 2-weeks-ahead | 0.72 | [0.47, 0.86] | P < 0.001 |
| RobertWalraven-ESG | 3-weeks-ahead | 0.75 | [0.53, 0.88] | P < 0.001 |
| RobertWalraven-ESG | 4-weeks-ahead | 0.79 | [0.59, 0.90] | P < 0.001 |

##### H.9.2 Predictability versus 95% Prediction Interval (PI) Coverage

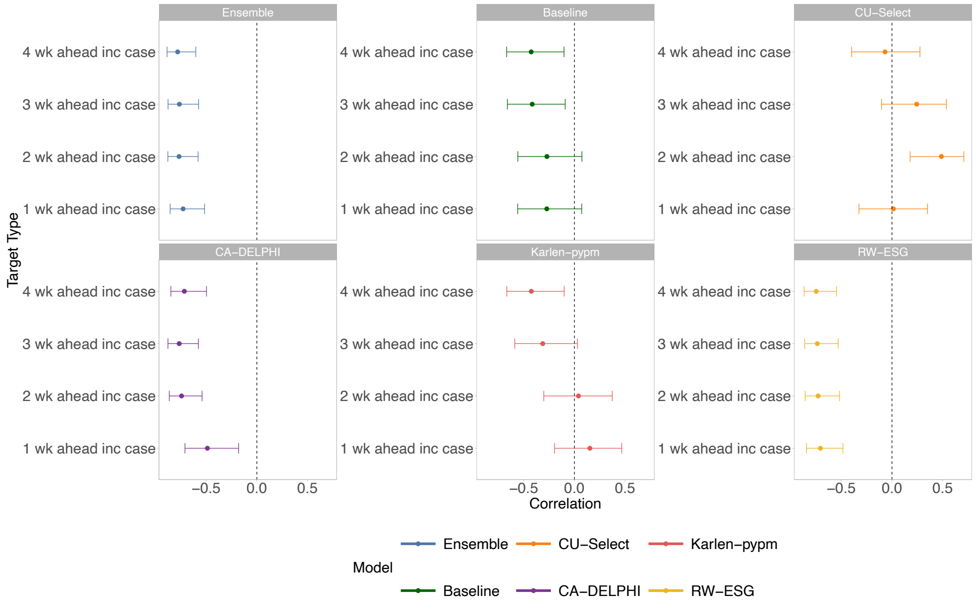


Figure SI 43: **Pre-wave correlations between predictability and central 95% prediction interval (PI) coverage:** In Arizona before the onset of the studied wave (starting July 2021), we visualise the Pearson's correlations (Table SI 5) between predictability and 95% PI coverage for each model and forecast target type, alongside the 95% Confidence Intervals (CIs) calculated using Fisher z-transformations.

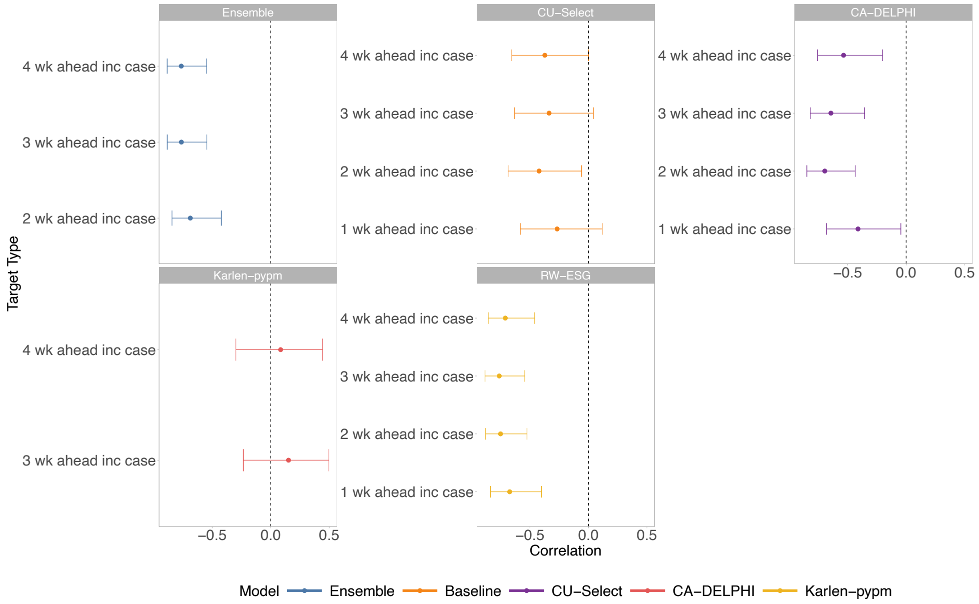


Figure SI 44: **During-wave correlations between predictability and central 95% prediction interval (PI) coverage:** In Arizona during the studied wave (starting July 2021), we visualise the Pearson's correlations (Table SI 5) between predictability and 95% PI coverage for each model and forecast target type, alongside the 95% Confidence Intervals (CIs) calculated using Fisher z-transformations. Note that no correlations are visualised for the one-week-ahead forecasts of the ensemble model, and for the two-weeks-ahead forecasts of the Karlen-pypm model, as these models had only one value within each trailing window for 95% PI coverage; 100%.

Table SI 9: **Pre-wave correlations between predictability and central 95% Prediction Interval (PI) coverage:** In Arizona before the onset of the studied wave (starting July 2021), for each model and forecasted quantity, we report the Pearson's correlation $r$ between predictability and 95% PI, alongside the 95% Confidence Intervals (CIs) calculated using Fisher z-transformations and the Bonferroni-adjusted p-value for the two-sided hypothesis test with null hypothesis that $r = 0$.

| Model | Target Type | r | 95% CI | Bonferroni-adjusted $p$ |
|---|---|---|---|---|
| Ensemble | 1-week-ahead | -0.73 | [-0.86, -0.52] | P < 0.001 |
| Ensemble | 2-weeks-ahead | -0.77 | [-0.88, -0.58] | P < 0.001 |
| Ensemble | 3-weeks-ahead | -0.76 | [-0.88, -0.57] | P < 0.001 |
| Ensemble | 4-weeks-ahead | -0.78 | [-0.89, -0.60] | P < 0.001 |
| COVIDhub-baseline | 1-week-ahead | -0.27 | [-0.56, 0.07] | P = 1 |
| COVIDhub-baseline | 2-weeks-ahead | -0.27 | [-0.56, 0.07] | P = 1 |
| COVIDhub-baseline | 3-weeks-ahead | -0.42 | [-0.66, -0.09] | P = 0.344 |
| COVIDhub-baseline | 4-weeks-ahead | -0.43 | [-0.67, -0.10] | P = 0.288 |
| CU-select | 1-week-ahead | 0.01 | [-0.33, 0.35] | P = 1 |
| CU-select | 2-weeks-ahead | 0.49 | [0.18, 0.71] | P = 0.0824 |
| CU-select | 3-weeks-ahead | 0.24 | [-0.10, 0.54] | P = 1 |
| CU-select | 4-weeks-ahead | -0.07 | [-0.40, 0.28] | P = 1 |
| CovidAnalytics-DELPHI | 1-week-ahead | -0.49 | [-0.71, -0.18] | P = 0.0821 |
| CovidAnalytics-DELPHI | 2-weeks-ahead | -0.74 | [-0.86, -0.54] | P < 0.001 |
| CovidAnalytics-DELPHI | 3-weeks-ahead | -0.77 | [-0.88, -0.58] | P < 0.001 |
| CovidAnalytics-DELPHI | 4-weeks-ahead | -0.71 | [-0.85, -0.50] | P < 0.001 |
| Karlen-pypm | 1-week-ahead | 0.15 | [-0.20, 0.47] | P = 1 |
| Karlen-pypm | 2-weeks-ahead | 0.04 | [-0.30, 0.37] | P = 1 |
| Karlen-pypm | 3-weeks-ahead | -0.31 | [-0.59, 0.03] | P = 1 |
| Karlen-pypm | 4-weeks-ahead | -0.42 | [-0.67, -0.10] | P = 0.296 |
| RobertWalraven-ESG | 1-week-ahead | -0.71 | [-0.84, -0.48] | P < 0.001 |
| RobertWalraven-ESG | 2-weeks-ahead | -0.73 | [-0.86, -0.52] | P < 0.001 |
| RobertWalraven-ESG | 3-weeks-ahead | -0.74 | [-0.86, -0.53] | P < 0.001 |
| RobertWalraven-ESG | 4-weeks-ahead | -0.75 | [-0.87, -0.55] | P < 0.001 |

Table SI 10: **During-wave correlations between predictability and central 95% Prediction Interval (PI) coverage:** In Arizona during the studied wave (starting July 2021), for each model and forecasted quantity, we report the Pearson's correlation $r$ between predictability and 95% PI, alongside the 95% Confidence Intervals (CIs) calculated using Fisher z-transformations and the Bonferroni-adjusted p-value for the two-sided hypothesis test with null hypothesis that $r = 0$.

| Model | Target Type | r | 95% CI | Bonferroni-adjusted $p$ |
|---|---|---|---|---|
| Ensemble | 1-week-ahead | NA | NA | NA |
| Ensemble | 2-weeks-ahead | -0.69 | [-0.84, -0.42] | P < 0.001 |
| Ensemble | 3-weeks-ahead | -0.76 | [-0.88, -0.55] | P < 0.001 |
| Ensemble | 4-weeks-ahead | -0.76 | [-0.89, -0.55] | P < 0.001 |
| COVIDhub-baseline | 1-week-ahead | NA | NA | NA |
| COVIDhub-baseline | 2-weeks-ahead | NA | NA | NA |
| COVIDhub-baseline | 3-weeks-ahead | NA | NA | NA |
| COVIDhub-baseline | 4-weeks-ahead | NA | NA | NA |
| CU-select | 1-week-ahead | -0.27 | [-0.58, 0.12] | P = 1 |
| CU-select | 2-weeks-ahead | -0.42 | [-0.69, -0.06] | P = 0.432 |
| CU-select | 3-weeks-ahead | -0.34 | [-0.63, 0.04] | P = 1 |
| CU-select | 4-weeks-ahead | -0.37 | [-0.65, 0.00] | P = 0.865 |
| CovidAnalytics-DELPHI | 1-week-ahead | -0.41 | [-0.68, -0.05] | P = 0.504 |
| CovidAnalytics-DELPHI | 2-weeks-ahead | -0.70 | [-0.85, -0.44] | P < 0.001 |
| CovidAnalytics-DELPHI | 3-weeks-ahead | -0.64 | [-0.82, -0.36] | P = 0.00381 |
| CovidAnalytics-DELPHI | 4-weeks-ahead | -0.53 | [-0.76, -0.20] | P = 0.0572 |
| Karlen-pypm | 1-week-ahead | NA | NA | NA |
| Karlen-pypm | 2-weeks-ahead | NA | NA | NA |
| Karlen-pypm | 3-weeks-ahead | 0.15 | [-0.23, 0.50] | P = 1 |
| Karlen-pypm | 4-weeks-ahead | 0.08 | [-0.30, 0.44] | P = 1 |
| RobertWalraven-ESG | 1-week-ahead | -0.67 | [-0.84, -0.40] | P = 0.00147 |
| RobertWalraven-ESG | 2-weeks-ahead | -0.75 | [-0.88, -0.53] | P < 0.001 |
| RobertWalraven-ESG | 3-weeks-ahead | -0.76 | [-0.88, -0.54] | P < 0.001 |
| RobertWalraven-ESG | 4-weeks-ahead | -0.71 | [-0.86, -0.46] | P < 0.001 |

##### H.9.3 Predictability versus relative Weighted Interval Score (rWIS)

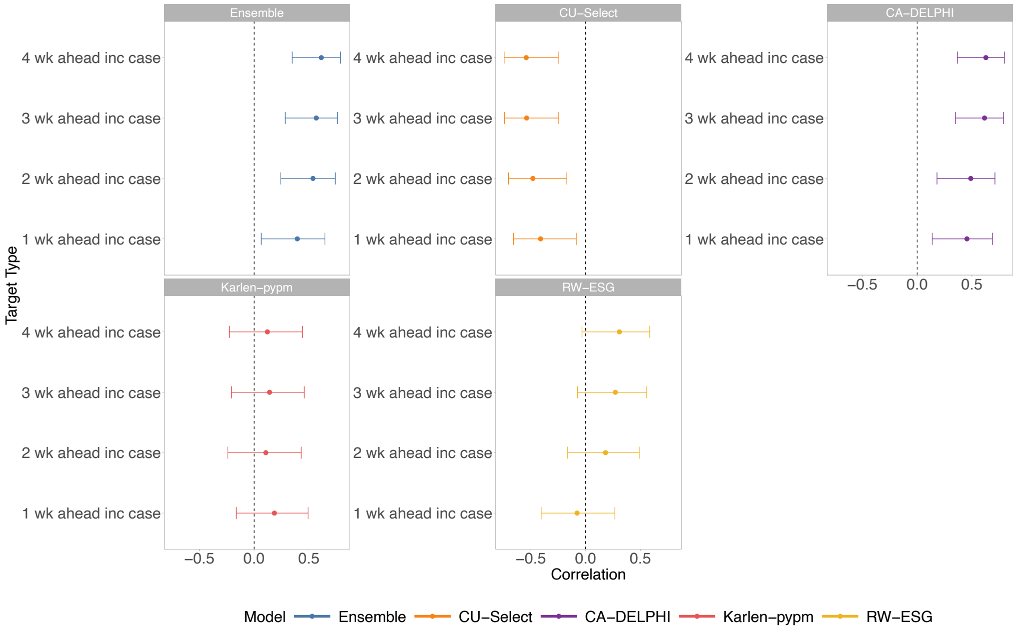


Figure SI 45: **Pre-wave correlations between predictability and relative Weighted Interval Score (rWIS)** In Arizona before the onset of the studied wave (starting July 2021), we visualise the Pearson's correlations (Table SI 6) between predictability and rWIS for each model and forecast target type, alongside the 95% Confidence Intervals (CIs) calculated using Fisher z-transformations.

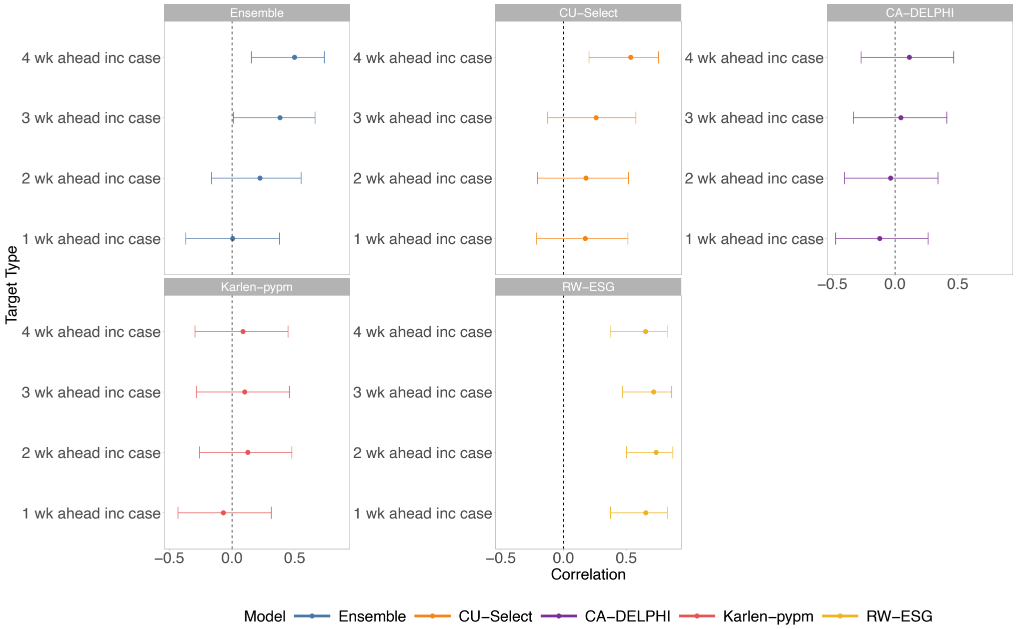


Figure SI 46: **During-wave correlations between predictability and relative Weighted Interval Score (rWIS)** In Arizona before the onset of the studied wave (starting July 2021), we visualise the Pearson's correlations (Table SI 6) between predictability and rWIS for each model and forecast target type, alongside the 95% Confidence Intervals (CIs) calculated using Fisher z-transformations.

Table SI 11: **Pre-wave correlations between predictability and relative Weighted Interval Score (rWIS)** In Arizona before the onset of the studied wave (starting July 2021), for each model and forecasted quantity, we report the Pearson's correlation $r$ between predictability and rWIS, alongside the 95% Confidence Intervals (CIs) calculated using Fisher z-transformations and the Bonferroni-adjusted p-value for the two-sided hypothesis test with null hypothesis that $r = 0$.

| Model | Target Type | r | 95% CI | Bonferroni-adjusted $p$ |
|---|---|---|---|---|
| Ensemble | 1-week-ahead | 0.39 | [0.06, 0.65] | P = 0.421 |
| Ensemble | 2-weeks-ahead | 0.54 | [0.24, 0.74] | P = 0.021 |
| Ensemble | 3-weeks-ahead | 0.57 | [0.28, 0.76] | P = 0.009 |
| Ensemble | 4-weeks-ahead | 0.61 | [0.35, 0.79] | P = 0.002 |
| COVIDhub-baseline | 1-week-ahead | NA | NA | NA |
| COVIDhub-baseline | 2-weeks-ahead | NA | NA | NA |
| COVIDhub-baseline | 3-weeks-ahead | NA | NA | NA |
| COVIDhub-baseline | 4-weeks-ahead | NA | NA | NA |
| CU-select | 1-week-ahead | -0.41 | [-0.66, -0.09] | P = 0.312 |
| CU-select | 2-weeks-ahead | -0.48 | [-0.71, -0.17] | P = 0.078 |
| CU-select | 3-weeks-ahead | -0.54 | [-0.74, -0.25] | P = 0.0201 |
| CU-select | 4-weeks-ahead | -0.54 | [-0.74, -0.25] | P = 0.0184 |
| CovidAnalytics-DELPHI | 1-week-ahead | 0.45 | [0.14, 0.69] | P = 0.139 |
| CovidAnalytics-DELPHI | 2-weeks-ahead | 0.49 | [0.18, 0.71] | P = 0.0662 |
| CovidAnalytics-DELPHI | 3-weeks-ahead | 0.61 | [0.35, 0.79] | P = 0.002 |
| CovidAnalytics-DELPHI | 4-weeks-ahead | 0.63 | [0.37, 0.80] | P = 0.00143 |
| Karlen-pypm | 1-week-ahead | 0.19 | [-0.16, 0.49] | P = 1 |
| Karlen-pypm | 2-weeks-ahead | 0.11 | [-0.24, 0.43] | P = 1 |
| Karlen-pypm | 3-weeks-ahead | 0.14 | [-0.21, 0.46] | P = 1 |
| Karlen-pypm | 4-weeks-ahead | 0.12 | [-0.23, 0.44] | P = 1 |
| RobertWalraven-ESG | 1-week-ahead | -0.08 | [-0.41, 0.27] | P = 1 |
| RobertWalraven-ESG | 2-weeks-ahead | 0.18 | [-0.17, 0.49] | P = 1 |
| RobertWalraven-ESG | 3-weeks-ahead | 0.27 | [-0.07, 0.56] | P = 1 |
| RobertWalraven-ESG | 4-weeks-ahead | 0.31 | [-0.03, 0.59] | P = 1 |

Table SI 12: **During-wave correlations between predictability and relative Weighted Interval Score (rWIS)** In Arizona during the studied wave (starting July 2021), for each model and forecasted quantity, we report the Pearson's correlation $r$ between predictability and rWIS, alongside the 95% Confidence Intervals (CIs) calculated using Fisher z-transformations and the Bonferroni-adjusted p-value for the two-sided hypothesis test with null hypothesis that $r = 0$.

| Model | Target Type | r | 95% CI | Bonferroni-adjusted $p$ |
|---|---|---|---|---|
| Ensemble | 1-week-ahead | 0.01 | [-0.37, 0.38] | P = 1 |
| Ensemble | 2-weeks-ahead | 0.22 | [-0.16, 0.55] | P = 1 |
| Ensemble | 3-weeks-ahead | 0.38 | [0.01, 0.66] | P = 0.897 |
| Ensemble | 4-weeks-ahead | 0.50 | [0.15, 0.73] | P = 0.139 |
| COVIDhub-baseline | 1-week-ahead | NA | NA | NA |
| COVIDhub-baseline | 2-weeks-ahead | NA | NA | NA |
| COVIDhub-baseline | 3-weeks-ahead | NA | NA | NA |
| COVIDhub-baseline | 4-weeks-ahead | NA | NA | NA |
| CU-select | 1-week-ahead | 0.17 | [-0.21, 0.51] | P = 1 |
| CU-select | 2-weeks-ahead | 0.18 | [-0.21, 0.52] | P = 1 |
| CU-select | 3-weeks-ahead | 0.26 | [-0.13, 0.58] | P = 1 |
| CU-select | 4-weeks-ahead | 0.54 | [0.20, 0.76] | P = 0.0665 |
| CovidAnalytics-DELPHI | 1-week-ahead | -0.12 | [-0.47, 0.26] | P = 1 |
| CovidAnalytics-DELPHI | 2-weeks-ahead | -0.04 | [-0.40, 0.34] | P = 1 |
| CovidAnalytics-DELPHI | 3-weeks-ahead | 0.05 | [-0.33, 0.41] | P = 1 |
| CovidAnalytics-DELPHI | 4-weeks-ahead | 0.11 | [-0.27, 0.47] | P = 1 |
| Karlen-pypm | 1-week-ahead | -0.07 | [-0.43, 0.31] | P = 1 |
| Karlen-pypm | 2-weeks-ahead | 0.13 | [-0.26, 0.48] | P = 1 |
| Karlen-pypm | 3-weeks-ahead | 0.10 | [-0.28, 0.46] | P = 1 |
| Karlen-pypm | 4-weeks-ahead | 0.09 | [-0.30, 0.45] | P = 1 |
| RobertWalraven-ESG | 1-week-ahead | 0.66 | [0.37, 0.83] | P = 0.003 |
| RobertWalraven-ESG | 2-weeks-ahead | 0.74 | [0.50, 0.87] | P < 0.001 |
| RobertWalraven-ESG | 3-weeks-ahead | 0.72 | [0.47, 0.86] | P < 0.001 |
| RobertWalraven-ESG | 4-weeks-ahead | 0.65 | [0.37, 0.83] | P = 0.003 |

### I Distributionally robust optimization for risk-averse decision-making

*Expected tail loss* (ETL) (also known as the *expected shortfall* or *conditional value at risk*) is a quantity central to the distributionally robust optimisation (DRO) literature [69, 70]. ETL measures the risk associated to extreme events occurring at the upper tail of the loss distribution. Specifically, with $Y$ denoting the outcome of interest (e.g., deaths, hospitalisations, or cases), ETL at level $\alpha$ is defined as[1]

$$\mathrm{ETL}_\alpha = \frac{1}{\alpha}\mathbb{E}\left[Y \cdot 1(Y \geq y_\alpha)\right],$$

where $y_\alpha$ is the $(1-\alpha)$-quantile of the distribution of $Y$ (also known as the *value at risk* (VaR)). In words, ETL is the average loss incurred among the worst $\alpha$% of outcomes. It is known that ETL and some other risk measures $T$ of interest (such as the variance and mode of $Y$) are not *elicitable* from any proper scoring rule $S$, in the sense that there is no $S$ for which predictions issued on the basis of $T$ uniquely minimize the score [71, 72].[2] In other words, evaluating forecasts based on $S$ would yield conclusions inconsistent with the choice of $T$ as a risk measure.

Arising within operations research and statistical learning, DRO is an alternative framework for decision-making under uncertainty. Above, we hinted at possible uncertainty in users' C/L ratios and in model rankings based on users' choice of evaluation trailing windows and spatial scale. Meanwhile, observed data during an epidemic are imperfect, impacted by time- and space-varying issues such as reporting errors, delayed reporting, and underrepresentation. All these issues can induce data distribution shifts and lead to metric-implied optimal decisions, from results on past data, no longer being optimal when making the decision in real-time on newly available data. Nevertheless, when seeking to make timely decisions (e.g., expanding ICU capacity) about important epidemic events (e.g., size of epidemic peak), a decision-maker would like the model-implied decision to be robust to uncertainty, distributional shifts, and worst-case scenarios.

We include DRO as a future tool for our framework to address these uncertainties in decision-making. Rather than assuming (unlikely) perfect knowledge of the outcome distribution $Y$ or, more generally, of some relevant random quantity $\xi$ indexing the loss function $h(x, \xi)$ associated to an action $x$, DRO attempts to minimise the worst-case cost of a decision across a range of plausible distributions of $\xi$. Mathematically, the DRO problem is

$$\inf_{x\in\mathcal{X}} \sup_{Q\in\mathcal{P}} \mathbb{E}^{Q}\left[h(x, \xi)\right], \tag{I.1}$$

where $x$ is a decision variable in feasibility set $\mathcal{X}$ and $\mathcal{P}$ is an *ambiguity set* of probability distributions $\xi \sim Q$. A common choice of ambiguity set is an $\epsilon$-neighborhood (with respect to some metric, discrepancy, or divergence, e.g., the Wasserstein metric or Kullback-Leibler divergence) around the empirical

[1] We assume for simplicity that $Y$ has a continuous distribution function, in which case ETL takes a more intuitive form.

[2] However, it is known that the pair (VaR, ETL) is jointly elicitable in a certain sense [72].

distribution $Q_N$ (or a density estimate) of the observations $(\xi_1, \ldots, \xi_N)$. Such a choice imparts robustness to uncertainty and shifts in the distribution of $\xi$. However, other ambiguity set constructions have been studied, yielding optimisation problems with different properties. In many cases, the resulting DRO problem (I.1) admits a computationally tractable formulation or relaxation as a convex program—often a linear program (LP), second-order cone program (SOCP), or semidefinite program (SDP) [70]. For example, ETL minimisation can be formulated as a DRO problem and as an LP amenable to efficient numerical optimisation via standard convex solvers [69].
